# How inherited variation shapes human disease trajectories

**DOI:** 10.64898/2026.11.11.26367654

**Authors:** Sarah M. Urbut, Buu Truong, Shaan Khurshid, Whitney Hornsby, Heidi L. Rehm, Patrick T. Ellinor, Pradeep Natarajan, Giovanni Parmigiani, Alexander Gusev

## Abstract

Although inherited variation is fixed at birth, its effects on health change with age and differ between common and rare variants. Here, we introduce Mendelnoulli, a longitudinal model that combines the age-dependent effects of polygenic and monogenic variation on health records, separating the overall disease accumulation of each individual from the diseases to which they are prone. Mendelnoulli further allows polygenic and monogenic components to exert different influences on latent disease trajectories. We apply the model to 348 diseases, 36 polygenic scores, and 34 monogenic conditions across three biobanks and provide an atlas of the age-dependent effects of inherited variation on common human diseases. We show that polygenic effects change significantly with age. Mendelnoulli improved held-out one-year discrimination for all 28 diseases tested compared with a model whose polygenic effects are constant across age (mean AUC gain 0.023). For individuals with no prior diagnoses, Mendelnoulli identified 8.6 more cases of every 100 new cases than that model, when each flagged the 5% at highest risk. For genes with an established disease, Mendelnoulli selected that disease for 27 of 30 genes and ranked it first for 17 (7 expected by chance), and recovered known differences in timing, such as earlier breast cancer effects for BRCA1 than BRCA2 (peak 48 vs 66 years). Mendelnoulli models common and rare genetic influences on disease trajectories jointly throughout the life course.

---

Many common human diseases are partly determined by inherited genetic factors [1, 2], yet a genetic susceptibility present from birth need not have a constant effect across the life course. The per-standard-deviation effect of a polygenic score is often largest early in life and declines with age [3, 4, 5]. Prevention therefore requires more than identifying who is at inherited risk. It requires understanding *when* that risk becomes clinically important, *what* phenotype it produces, and how it is modified by other events over the life course.

Two observations motivate our work. First, when polygenic effects attenuate with age (for example, due to the accumulation of environmental factors), they violate the proportional-hazards assumption used by many risk models. A clinically important consequence is that a polygenic score may carry substantial predictive information for rapidly progressing individuals before related diagnoses accumulate [3, 4]; we test this below. Second, pathogenic variants typically generate syndromic phenotypes, distinctive sets of diseases [6, 7], that are not reproduced by simply moving farther into the tail of a polygenic score for any one of the constituent diseases.

Rare variants have been studied across many diseases [8, 9] and within disease areas [10], and common and low-frequency variants in genome-wide studies [11]. Multiple lines of evidence suggest that combining them can improve risk prediction. Penetrance estimates from unselected biobanks are lower than the estimates from clinically ascertained families [12, 13], reflecting ascertainment; the same rare variant may confer different risk in different genetic backgrounds [14, 15]. Within carriers of tier-1 genomic conditions [16], polygenic background can shift the risk of the syndrome’s index disease across a wide range [17]. Likewise, a sufficiently extreme polygenic score can confer risk of that same disease comparable to a pathogenic variant [18]. Models that combine the two sources of risk exist for single diseases [19, 20], but to our knowledge, none has combined them across many diseases and organ systems at once.

Several approaches model many diseases jointly. Our previous model, Aladynoulli [21], represents longitudinal disease histories as mixtures of latent signatures of related diseases, with individual loadings that change with age. Age-dependent topic models of comorbidity in the UK Biobank identified disease subtypes with different genetic associations [22], and generative transformers trained on health records forecast the timing of more than a thousand diagnoses [23], with polygenic scores improving these forecasts [24]. In these models, genetic information is one input among many; they predict well but do not estimate how much disease an inherited factor adds, which diseases it affects, or at what ages. Life-course analyses of polygenic scores have mostly considered one trait at a time, such as weight from birth to adulthood [25] or coronary artery disease by age [26], and have used common variants only. Modeling many diseases jointly has two advantages for genetic questions. A genetic effect on one disease is estimated together with all the other diseases an individual develops, accounting for comorbidities; and the syndromic effects of a pathogenic variant, which span several diseases, can be estimated directly rather than one disease at a time.

Here we develop a model for improved disease prediction and for characterizing how inherited risk is expressed temporally across many conditions. We show that when germline information is modeled across the life course and across the spectrum of human disease, it improves prediction. We provide both an atlas of polygenic and monogenic effects on disease and methods for clinically relevant stratification. Compared with the multi-disease models above, ours adds three elements. First, polygenic effects on disease signatures change with age and are estimated jointly across 348 diseases. Second, the effects of pathogenic variants are sparse [27] and age-varying, so the model selects which diseases each variant affects. Third, a health component that follows an age curve, together with a per-person rate, separates how much disease an individual accrues from which diseases. We validate our conclusions against model-free comparisons of carriers with high polygenic score groups, held-out prediction, penetrance in the linked national cancer registry, and replication in two independent cohorts.

## Results

### The model: two genetic mechanisms

Our approach begins from the Aladynoulli model [21], which uses clinical data to learn latent patterns of disease, which we call disease signatures, and their contributions to an individual’s disease course over time. Each signature represents a group of diseases with a characteristic pattern of onset. An individual’s loading on a signature varies with age.

Starting from this framework, we now model two distinct ways for inherited variation to act. First, polygenic variation can change an individual’s *loading* on a signature, making every disease in the signature either more or less likely together; we allow this genetic effect to increase or decrease with age. Second, a pathogenic variant can change the *composition* of a signature for its carriers, making one disease more likely relative to the other diseases in the same signature. For brevity, we use *rescaling* and *reshaping* for these two mechanisms. Two individuals with the same loading on the same signature at the same age can be prone to different diseases, depending on carrier status. In Aladynoulli [21], by contrast, polygenic scores entered only as a constant effect on the signature loading and therefore could not influence how an individual’s loading changed with age or which diseases within the signature they developed.

The model describes each individual’s record as a set of first diagnoses by age. The outcome, for each individual, disease, and year of age, is whether that disease is first recorded at that age; follow-up for a disease ends at its first record or at censoring. The probability of a first diagnosis is a mixture over signatures. Each signature assigns every disease an age-specific probability, and the individual’s loadings weight these probabilities. One further component represents health. In it, every disease is less probable than in the population as a whole, and its weight is highest at young ages and declines along the population’s disease-free curve. A per-person rate, or frailty, scales how much disease an individual accrues overall, without changing which diseases they are prone to.

Formally, let *Y*_*idt*_ = 1 denote the event where disease *d* is first recorded for individual *i* at age *t*, and 0 otherwise. Let *E*_*id*_ represent the last age at which *i* is at risk for *d*. For signature *k*,

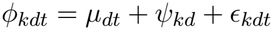

is the log-odds of a first diagnosis of *d* at age *t* for a non-carrier with all weight on that signature, where *µ*_*dt*_ is the population log-odds, *ψ*_*kd*_ is the signature’s enrichment for disease *d*, and *ϵ*_*kdt*_ is a smooth age-varying deviation with a Gaussian-process prior. Individual *i*’s loadings are

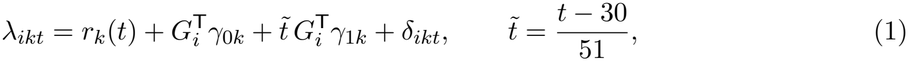

with weights *θ*_*i·t*_ = softmax(*λ*_*i·t*_), where *r*_*k*_(*t*) is a population-level age trajectory, *G*_*i*_ the individual’s polygenic scores and sex, *γ*_0*k*_ their effect on the loading, *γ*_1*k*_ the change in that effect with age, and *δ*_*ikt*_ a smooth per-person deviation learned from the individual’s own record. With *g*_*im*_ = 1 if individual *i* carries a pathogenic variant in gene *m*, the probability of a first diagnosis is

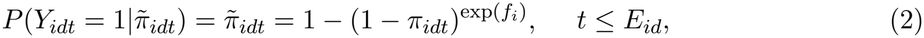

with

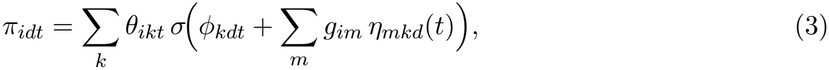

where *σ* is the logistic function, *η*_*mkd*_(*t*) is the effect of carrying a variant in gene *m* on the log-odds of disease *d* in signature *k* at age *t*, and *f*_*i*_ is the individual’s frailty.

We impose sparsity on the carrier effect with a ‘spike-and-slab’ prior, *η*_*mkd*_(*t*) = scope_*mk*_ *z*_*mkd*_ *v*_*mkd*_(*t*) The indicator scope_*mk*_ assigns each gene to a signature, usually the one in which the gene’s primary disease is most enriched (Methods). Within that signature, a binary selection indicator *z*_*mkd*_ records whether disease *d* has a non-zero carrier effect; a penalty on the number of selected diseases keeps the selected set small. The slab *v*_*mkd*_(*t*) is the size of the effect, with a Gaussian-process prior over age, to flexibly estimate the age at which a carrier effect is more pronounced.

The two genetic terms enter the model in different equations, changing disease risk in different ways (Fig. 1a). A polygenic score alters an individual’s loadings, which in turn multiplies the probability of every disease in a signature by the same factor, so the ratio of the probabilities of any two diseases within the signature remains unchanged. We refer to this as rescaling. A polygenic score, however extreme, moves weight between signatures but leaves each signature’s composition fixed (Fig. 3a,b). In contrast, the carrier effect changes the probability of one disease within a signature, and therefore its probability relative to the other diseases in that signature. We refer to this as reshaping. Only the carrier term can reshape a signature.

**Figure 1:**
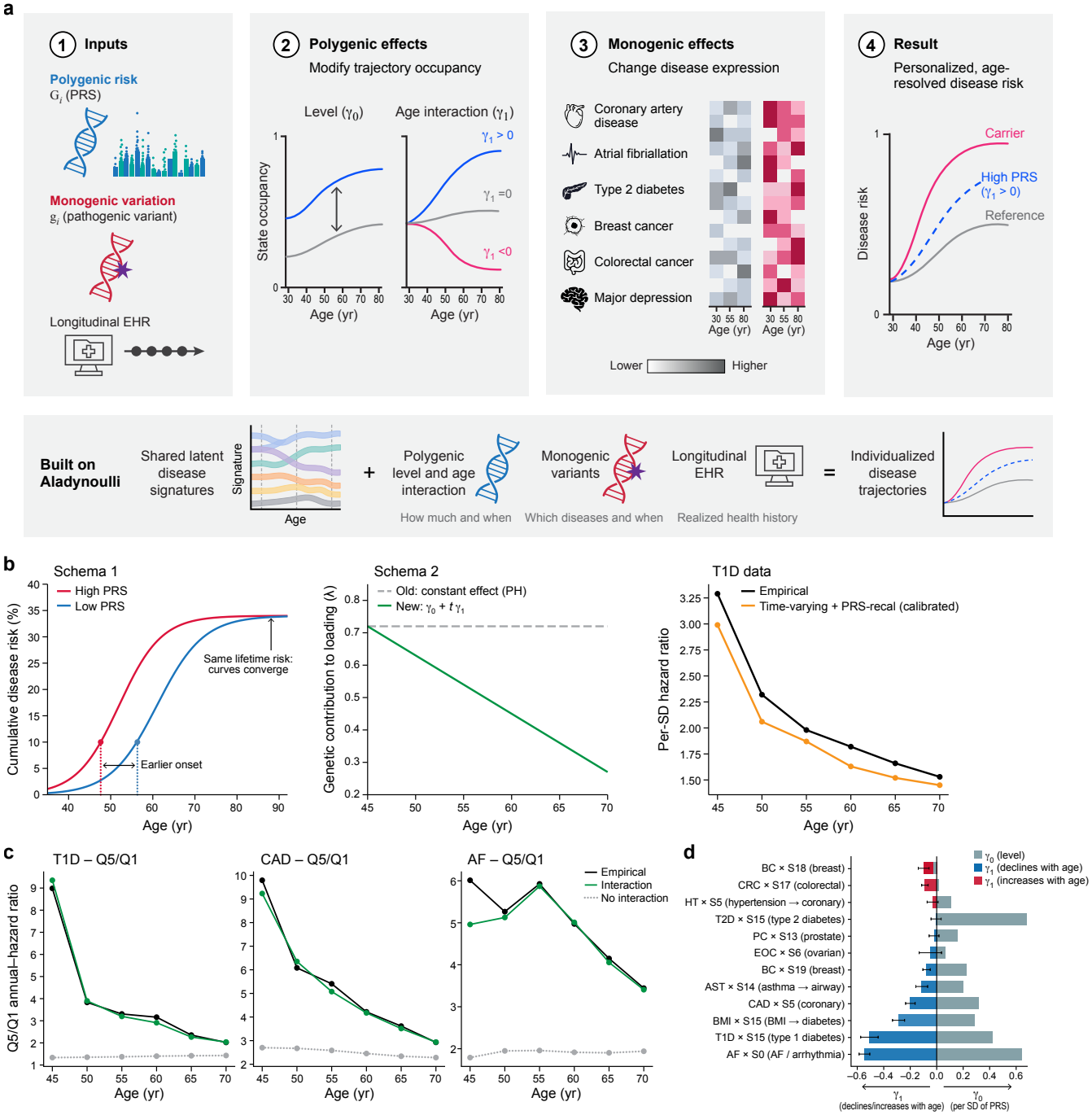
The model and the age dependence of polygenic effects. *(a)* Diseases are grouped into signatures, and each person’s loading on each signature changes with age. Polygenic scores change the loading (a level effect, *γ*_0_) and how it changes with age (an age effect, *γ*_1_); a pathogenic variant raises specific diseases within a signature (the carrier effect, *η*(*t*)). *(b)* With an age effect, a polygenic score need not have the same relative effect at every age. The hazard ratio per standard deviation of the type 1 diabetes score in five-year age windows, observed and from the model. *(c)* Ratio of event rates between the top and bottom score quintiles by age for type 1 diabetes, coronary artery disease and atrial fibrillation: observed, the full model, and the model without the age effect. Model curves use a per-age recalibration of the score effect (Methods). *(d)* Level and age effects for 12 scores, each paired with the signature containing its disease (mean across 40 training batches; *±* 1 standard error for the age effect). The level effect is positive for 11 of the 12 pairs.

We evaluate three ablations of the model as comparators, each obtained by removing one term from the *full model* above: the *no-interaction* model (*γ*_1_ ≡ 0), in which each polygenic score has a constant effect on the loadings over age; the model without the carrier term (*η* ≡ 0); and the *age-constant* model, in which the carrier effect does not vary with age.

We fit the model to the electronic health records of 400,000 UK Biobank participants [28]: *D* = 348 diseases defined by phecodes, first diagnoses in single years of age from 30 to 81 (*T* = 52), *K* = 20 signatures, and 36 polygenic scores [29] plus sex, with scores adjusted for the first ten genetic principal components. Carrier analyses use ClinVar [30] pathogenic and likely pathogenic variants in the 200,000 exome-sequenced participants [31]. Every term, with its dimension and whether it is data, fixed before fitting or learned, is listed in Table S1; estimation is described in Methods.

### Polygenic susceptibilities change with age

Polygenic scores are often modeled as having a constant effect at every age. We asked whether this assumption holds and whether an age interaction in the model captures departures from it. For ten illustrative disease–score pairs, we compared the model with two empirical measures: the hazard ratio per standard deviation of the score, estimated in five-year age windows, and the ratio of event rates between the top and bottom quintiles.

As expected [3, 4], the empirical hazard ratio declined with age for most pairs, and the full model reproduced the direction of the empirical trend in all ten pairs (binomial *p* = 0.002; Fig. 1b,c). For type 1 diabetes, for example, the ratio of event rates between the top and bottom score quintiles fell from 9.0 at age 45 to 2.0 at age 70, and the full model captured this decline (9.1 to 2.0). Each score acts through signatures that jointly model 348 diseases, so the model’s age effect is adjusted for comorbidities. The decline supports the observation that a polygenic score conveys most information early in life. In the fitted model, this decline arises from an age effect that reduces a positive overall level effect, most clearly for scores such as atrial fibrillation and type 1 diabetes (Fig. 1d).

### Age-varying polygenic effects improve prediction, most for individuals with little recorded history

We first asked whether modeling the age interaction improves the prediction of new diagnoses. We measured discrimination as the area under the receiver operating characteristic curve (AUC) for a first diagnosis of each of 28 diseases in the year after enrollment, in held-out participants, with both models fit so that they differ only in the age interaction.

The interaction model had a higher AUC for all 28 diseases (binomial *p <* 10^−8^), by 0.023 on average (95% CI across diseases 0.018–0.028). The improvement held regardless of prior history: when participants were grouped by the number of diseases recorded before enrollment (0, 1–2, 3–5, 6 or more), the AUC increased in 32 of 36 disease–group combinations (binomial *p <* 10^−5^), with no decline in the gain as the number of prior diagnoses increased (Fig. 2c).

**Figure 2:**
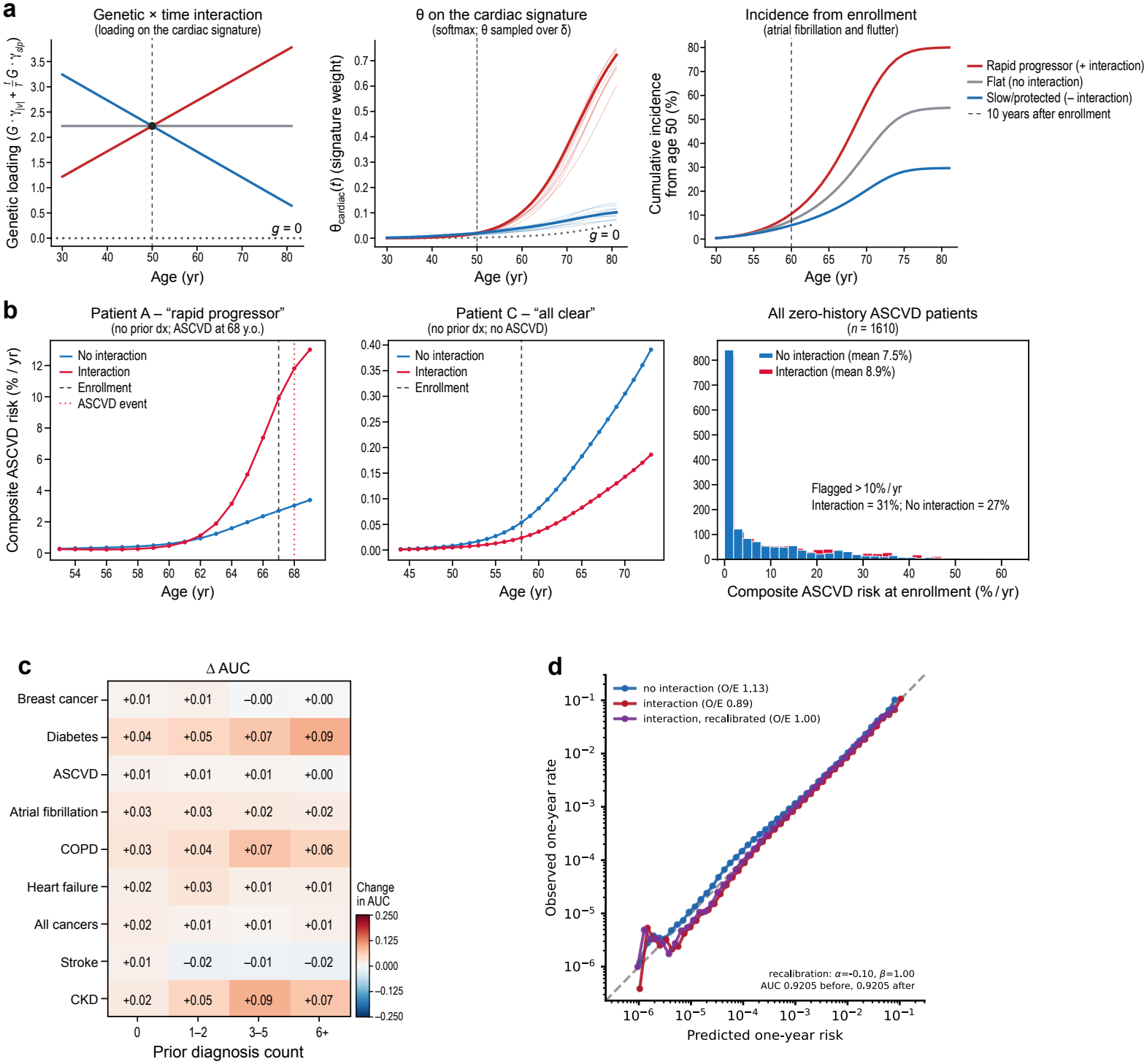
The genetic age interaction changes individual predictions before the first event. *(a) Mechanism (constructed from the fitted model)*. Three hypothetical genetic profiles built from the fitted cardiac-signature coefficients, with the same loading *θ* on that signature at age 50 and differing only in their genetic age term *G*_*i*_*γ*_1*k*_, set to the 10th percentile, zero and the 90th percentile of its distribution in the cohort (*left* ); genetic effects on other signatures are set to zero. A positive age term increases the loading with age and a negative one lets it decline (*middle*; trajectories sampled over *δ*), so ten-year atrial fibrillation risk reaches 10% against 6% from the same starting value (*right* ). The profiles are constructed, not selected participants; in real records the level effect *γ*_0*k*_ and the age interaction *γ*_1*k*_ act together. *(b) Participants with no prior diagnoses*. Among the 1,610 participants with no diagnoses before enrollment who developed ASCVD within two years, the mean predicted risk at enrollment was 8.9% per year with the age interaction and 7.5% without it (Wilcoxon signed-rank *p <* 10^−80^), and 31% against 27% exceeded 10% per year (McNemar *p <* 10^−16^; *right* ). Patient A (*left* ; no prior diagnoses, ASCVD at 68) has predicted risk 2.7% per year without the interaction and 9.9% with it. Patient C (*middle*) has the same empty record and no ASCVD during follow-up; both models predict low risk. Patients A and C are modified from participants’ trajectories. *(c) Gain by number of prior diagnoses*. Gain in one-year AUC in held-out participants, with against without the age interaction (both models refit identically, so they differ only in that term), in 36 disease *×* prior-diagnosis groups (0, 1–2, 3–5 and 6 or more distinct diagnoses before enrollment). The gain is positive in 32 of 36 groups (binomial *p <* 10^−5^) and does not decline with more prior diagnoses; the exceptions are three stroke groups with few events and one breast cancer group with no change. *(d) Calibration*. Observed against predicted one-year rate in held-out participants (log scale; bins of at least 10,000 person-years), for the model without the age interaction (observed/expected 1.13), the full model (0.89), and the full model after a monotone recalibration estimated on half of the participants and shown on the other half (1.00; *α* = −0.10, *β* = 1.00; AUC unchanged). The largest relative excess of predicted over observed events is at the lowest predicted risks. 24

**Figure 3:**
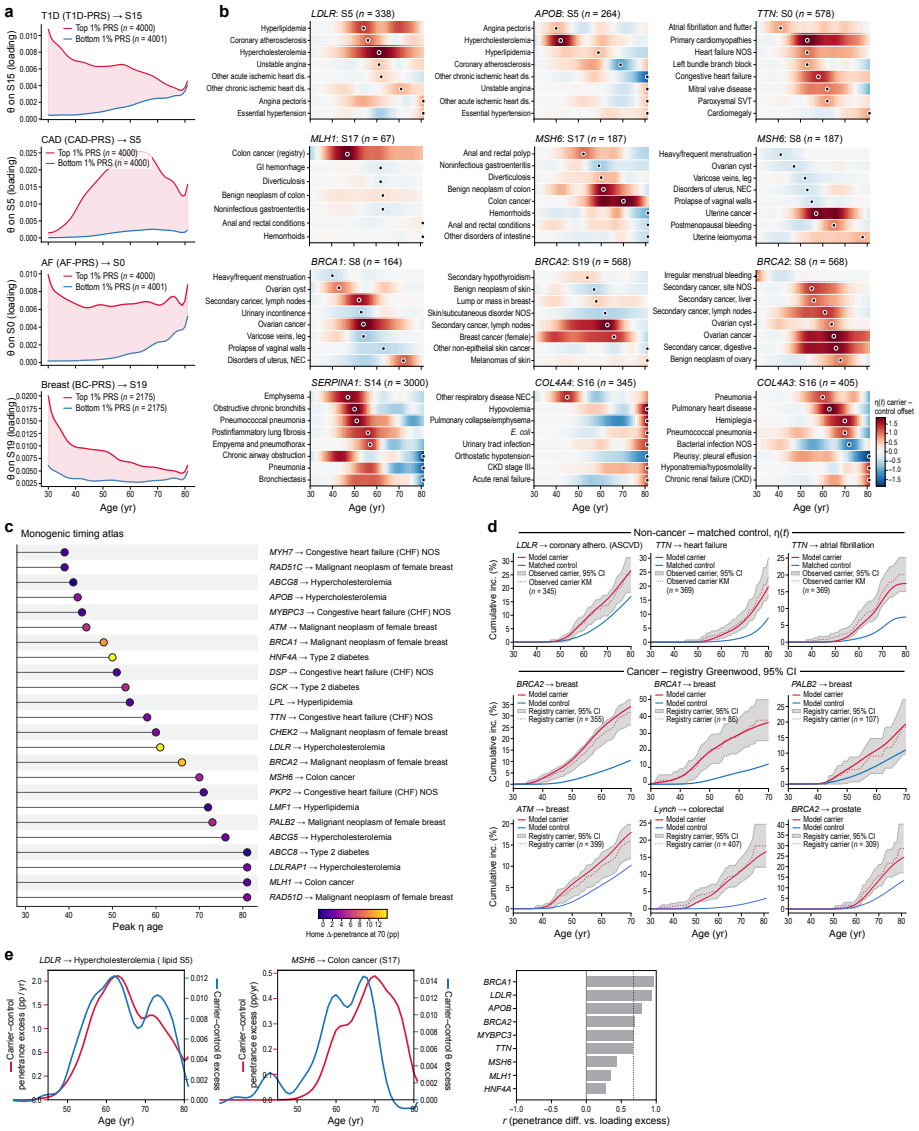
Polygenic variation rescales a signature, whereas pathogenic variants change its composition. *(a) Polygenic score and signature loading*. Mean loading over all individuals in the top and bottom 1% of each disease’s PRS (breast restricted to women; nothing refitted), for T1D (S15), CAD (S5), AF (S0) and breast cancer (S19). Raising the PRS increases the loading on a signature and leaves the relative risks of its diseases unchanged (Methods). In all four the polygenic contribution weakens with age, even where the loading itself rises; for T1D the top-to-bottom ratio of the share of non-healthy loading falls from 6.3 at age 40 to 0.9 by age 80. *(b) Pathogenic variants and signature composition*. The age-varying carrier effect *η*(*t*) raises specific diseases within the gene’s assigned signature, which changes the signature’s composition. *BRCA2* and *MSH6* are also shown fitted on S8, and *BRCA1* on S8 only; on S8, ovarian (*BRCA1, BRCA2* ) and uterine (*MSH6* ) cancer are selected. Color shows *η*(*t*). *(c) Timing*. Age at which the carrier effect on each gene’s main disease peaks (one fit per gene); color shows the carrier − control difference in cumulative incidence at age 70 (Δpen, percentage points). Peaks at 81 are still rising at the end of follow-up. *(d) Comparison with registries*. For the cancers, the carrier effect is estimated from diagnoses in the national cancer registry linked to UK Biobank. Model cumulative incidence for carriers (ClinVar P/LP; *TTN* truncating variants in highly expressed exons) and for age- and sex-matched non-carriers (band, carrier 95% CI), against the registry Kaplan–Meier curve with Greenwood 95% CI. All model carrier curves for the cancers lie within the registry CI at age 70. *(e) Agreement between penetrance and loading* (*LDLR* → hypercholesterolemia and *MSH6* → colon cancer shown). Over age, the carrier − control difference in penetrance is correlated with the carrier − control difference in signature loading (median *r* = 0.68 across genes), most strongly for *LDLR* (*r* = 0.94) and *BRCA1* (*r* = 0.97) and least for *HNF4A* (0.28) and the Lynch genes (*MLH1* 0.35, *MSH6* 0.43). The model-free comparison is Supp. Fig. S2 and Table 2; the 32-gene atlas is Supp. Table S7 and the FH signature 5 trajectory is Supp. Fig. S7.

We next asked whether the age interaction can explain part of the difference in future trajectories between individuals who look the same at enrollment. Using the fitted coefficients for the cardiac signature, which includes atrial fibrillation, we constructed three hypothetical genetic profiles with the same loading on that signature at age 50, differing only in their genetic age term (the 10^th^ percentile, zero, or the 90th percentile of its distribution in the cohort). Ten-year risk of atrial fibrillation reached 10% for the 90th-percentile profile and 6% for the 10th (Fig. 2a).

For individuals with no recorded history at enrollment, the interaction model stratifies risk better using polygenic scores. Among the 1,610 participants with no prior diagnoses who developed atherosclerotic cardiovascular disease (ASCVD) within two years, the mean predicted risk at enrollment was 8.9% per year with the interaction and 7.5% without it (Wilcoxon signed-rank *p <* 10^−80^), whereas in participants with equally empty records who remained free of ASCVD, the mean prediction changed by only 0.03 percentage points (Mann–Whitney *p <* 10^−100^ for the difference; Fig. 2b).

We then asked whether the gain differs among the people each model would flag for screening. For each disease, we let each model select the 5% of participants it considered at highest risk, and counted how many of the people diagnosed in the following year had been within that set. Among those diagnosed who had no prior diagnoses, the interaction model identified 8.6 more per 100 cases than the no-interaction model (1,746 found only by the interaction model, against 408 found only by the other; McNemar *p <* 10^−16^). Among those with six or more prior diagnoses, it found only 1.9 more per 100.

Finally, we asked whether the predicted risks are accurate in absolute terms. On held-out data, the model predicted about 12% more events than were observed (observed-to-expected ratio 0.89), with the largest relative excess at the lowest predicted risks (Fig. 2d). A single monotone recalibration, estimated on half of the participants and evaluated on the other half, brought the ratio to 1.00 without changing the AUC.

### Effects of pathogenic variants

First, without the model, we compared carriers of each gene with people at the top of the polygenic score for the gene’s primary disease, choosing the top fraction whose excess incidence of that disease, relative to everyone else, was closest to the carriers’ (Table 2). *BRCA1, BRCA2* and *PALB2* carriers had a 21.6 percentage-point excess of breast cancer by age 70, similar to the top 0.5% of the breast cancer polygenic score (19.7 points); yet carriers had a 6.5-point excess of ovarian cancer (95% CI 4.2–8.6) and the high-score group had none (0.0; −0.4–0.5). *TTN* carriers and the top 10% of the atrial fibrillation score had similar excesses of atrial fibrillation (5.5 and 6.1 points), but cardiomyopathy excesses of 3.8 and 0.3 points. No polygenic group reached the colon cancer excess of *MLH1* and *MSH6* carriers, so we compared secondary excess per point of primary excess:

0.70 (0.38–1.26) for uterine cancer in carriers, against at most 0.04 for any high-score group. At matched risk of the primary disease, carriers and high-score individuals differ in the other diseases they develop.

In the model, a polygenic score scales all diseases of a signature in proportion, whereas the carrier term uses the carriers’ records to select which diseases of the gene’s signature change (Fig. 3b).

Next, we asked whether the carrier effects recover each gene’s known disease, as a positive control. We fitted the model to carriers of ClinVar pathogenic and likely pathogenic variants among 200,000 exome-sequenced participants, specifying only the signature each gene acts on and allowing the model to select which diseases change. Before fitting, we assigned each gene a target disease family from its established clinical phenotype (for example, hypercholesterolemia for *LDLR*, colon cancer for *MLH1*, and cardiomyopathy or heart failure for *TTN* ; Methods). The target family was among the selected diseases for 27 of 30 genes (Table S7). Because many syndromes include several diseases, such as breast and ovarian cancer for *BRCA1* and *BRCA2*, the strongest selected effect need not fall on the target. It did for 17 of 30 genes, against 7.2 expected if the selected diseases were ordered at random (Poisson-binomial *p* = 10^−5^).

Allowing carrier effects to vary with age also estimates when each effect peaks. *BRCA1* and *BRCA2* illustrate this. Both genes increase the risk of breast cancer, and when allowed to act on the signature that encompasses most gynecological diagnoses, both also increase the risk of ovarian cancer. For both cancers, the effect peaked earlier in *BRCA1* carriers, at age 48 against 66 for breast cancer and 54 against 65 for ovarian cancer (Fig. 3b,c). The recorded diagnoses showed the same order: among UK Biobank women, breast cancer rates at ages 40–49 were 3.6 times higher in *BRCA1* than in *BRCA2* carriers (exact binomial *p* = 0.003), and excess ovarian cancer was highest at ages 50–59 in *BRCA1* carriers and at 60–79 in *BRCA2* carriers, although the *BRCA1* estimate rests on nine cases (rate ratio at ages 40–59, 2.4; *p* = 0.08; Supp. Fig. S1). The model recovers the established difference in timing between the two genes [32, 6].

We then asked whether the model provides a good fit to absolute carrier penetrance. For the cancers, we estimated the carrier effect from diagnoses in the national cancer registry linked to the UK Biobank and compared the model’s penetrance with the registry’s Kaplan–Meier estimate. By age 70, the model estimated that *BRCA2* carriers have a 34.1% cumulative incidence of breast cancer, compared with 31.5% in the registry, and every model carrier curve fell within the registry’s 95% confidence interval at age 70 (Fig. 3d).

Finally, the model represents a carrier’s risk in two places: the carrier effect on specific diseases and the carrier’s loading on the signature. We asked whether these two agree. Across genes, the carrier–control difference in penetrance was correlated over age with the carrier–control difference in signature loading (median Pearson *r* = 0.68; Fig. 3e).

### Selected diseases are specific to genes, stable, and match observed rates

Genes for monogenic conditions can affect diseases in different signatures, and two genes can also act on different diseases within the same signature. We next asked whether the model uses this flexibility. Five cardiac genes share the same assigned signature (S0, fixed before fitting; Methods), so any differences among them come from the diseases the model selects. The selections differed and matched each gene’s known phenotype: cardiomyopathy for *DSP* (carrier–control difference 11.1 percentage points), ventricular tachycardia for *MYBPC3* and *PKP2*, heart failure for *TTN*, and atrial fibrillation for *TTN, PKP2* and *MYH7* (Fig. 4a). When effects were allowed to vary with age, the model still selected diseases for every gene (*TTN* 15, *MYBPC3* 11, *PKP2* 9, *DSP* 7, *MYH7* 5), and all five selected heart failure (Fig. 4b).

**Figure 4:**
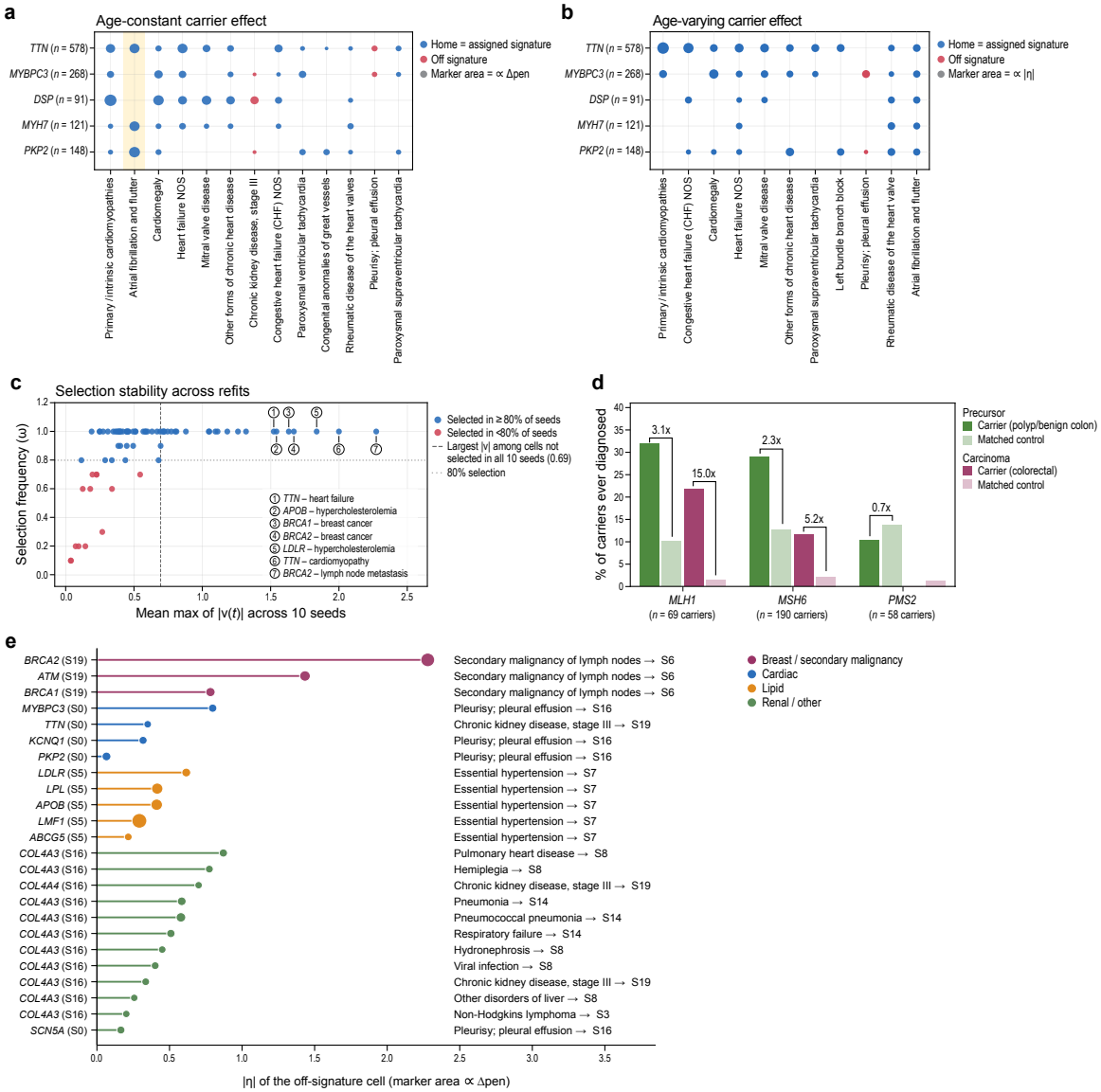
Five cardiac genes assigned to one signature select different diseases, and selection is stable. *(a) Age-constant carrier effects*. Diseases selected for the five cardiac genes, all assigned to signature S0, so differences between rows reflect only which diseases each gene selects: *TTN*, heart failure (Δpen, carrier − control difference in cumulative incidence at age 70 in percentage points, +5.4) and atrial fibrillation (+5.4); *DSP*, cardiomyopathy (+11.1) and cardiomegaly (+6.0); *MYBPC3*, cardiomegaly (+3.7) and ventricular tachycardia (+2.7); *PKP2*, atrial fibrillation (+6.5) and ventricular tachycardia (+1.8); *MYH7*, atrial fibrillation (+5.6). Atrial fibrillation (shaded) is selected for *TTN, PKP2* and *MYH7*. Columns are ordered by Δpen and marker area is proportional to it. Blue, the disease belongs mainly to the assigned signature; red, it belongs mainly to another (panel e). *(b) Age-varying carrier effects*. All five genes keep selected diseases (*TTN* 15, *MYBPC3* 11, *PKP2* 9, *DSP* 7, *MYH7* 5, including diseases not shown in the panel). *(c) Stability*. Selection frequency over 10 fits that differ only in random seed, against the mean peak carrier-effect magnitude max_*t*_ |*v*|, for six genes. Every gene–disease pair with mean |*v*| *>* 0.69 was selected in all 10 fits; every pair selected in fewer than 8 had |*v*| ≤ 0.54. We report pairs selected in at least 80% of fits. *(d) Observed rates without the model*. Diagnosis rates in ClinVar P/LP Lynch carriers against non-carriers matched 1:4 on sex and 5-year age group. Carriers have more colon polyps (*MLH1* 3.1 *×, MSH6* 2.3*×* ) and more colon cancer (*MLH1* 15 *×, MSH6* 5.2 *×*); *PMS2* carriers show neither (0.7*×* for polyps, no excess of cancer), which matches the known order of severity *MLH1 > MSH6 > PMS2. (e) Diseases outside the assigned signature*. Of the 307 gene–disease pairs selected across 34 genes (one fit per gene), 36 involve a disease that belongs mainly to another signature; the 24 of these that raise penetrance (Δpen *>* 0) are shown as gene (assigned signature) → disease → the disease’s main signature, ordered by |*η*| . Five of the 24 show a significant excess in carriers’ observed diagnoses over matched non-carriers (FDR *q <* 0.05 across the 307 pairs): secondary malignancy of the lymph nodes for *ATM, BRCA1* and *BRCA2*, and essential hypertension for *APOB* and *LMF1. COL4A3* and *COL4A4* contribute 11 of the 24, none with a significant excess.

We next asked whether these selections are stable when refitting the model with ten different random seeds. Every gene–disease pair with a large effect (mean magnitude above 0.69) was selected in all ten fits, and its effect size varied by at most 4% (Fig. 4c). Selection varied only for pairs with smaller effects (|*v*| ≤ 0.54). Finally, we compared these selections with empirical rates. In carriers of Lynch syndrome genes, matched to non-carriers on sex and age, observed rates of colorectal cancer followed the established order of severity, *MLH1* (21.7%) above *MSH6* (11.6%) above *PMS2* (0%) [7] (Fig. 4d).

Some carriers develop diseases that are primarily enriched in other signatures. We asked whether the model can capture these. Of the 307 gene–disease pairs selected across 34 genes, 36 involve such diseases, and 24 of these raise penetrance (Fig. 4e). Selected pairs are candidates, so we compared each with carriers’ observed diagnoses. Of the 24, 5 showed a significant excess over matched non-carriers (FDR *q <* 0.05): lymph-node metastasis for *ATM, BRCA1* and *BRCA2*, and hypertension for *APOB* and *LMF1* ; across all 307 pairs, 35 did. *COL4A3* and *COL4A4* contributed 74 of the 307 pairs and 11 of the 24, none with a significant excess.

### Carrier status, polygenic scores and the health record provide complementary information

Because our model combines carrier status and polygenic score multiplicatively on the underlying scale, we asked whether their empirical joint effect supports this and how much complementary information each provides.

First, for polygenic scores to add information in carriers, the carriers must not already be concentrated at high scores. We compared the distribution of all 36 scores between carriers and non-carriers. For 35 of the 36 scores, the standardized mean difference was within *±*0.1 (Supp. Fig. S9), as expected for unlinked variants. Polygenic scores therefore do not duplicate carrier status.

For genes of moderate effect, the polygenic gradient was similar among carriers and non-carriers (interaction odds ratio for *TTN*, 1.08; *p* = 0.59), whereas for high-penetrance cancer genes, the gradient was flatter in carriers (*BRCA2*, 0.72, 95% CI 0.57–0.92; Lynch genes, 0.69, 0.49–0.98; not adjusted for multiple testing; Fig. 5a,b). One explanation is that when a variant already confers high risk, each unit of polygenic score raises relative risk less; however, it may still raise absolute risk more because carriers start from a higher baseline. For *TTN* the data are consistent with the model’s multiplicative form; for high-penetrance cancer genes the polygenic score adds less on the relative scale than this form implies.

**Figure 5:**
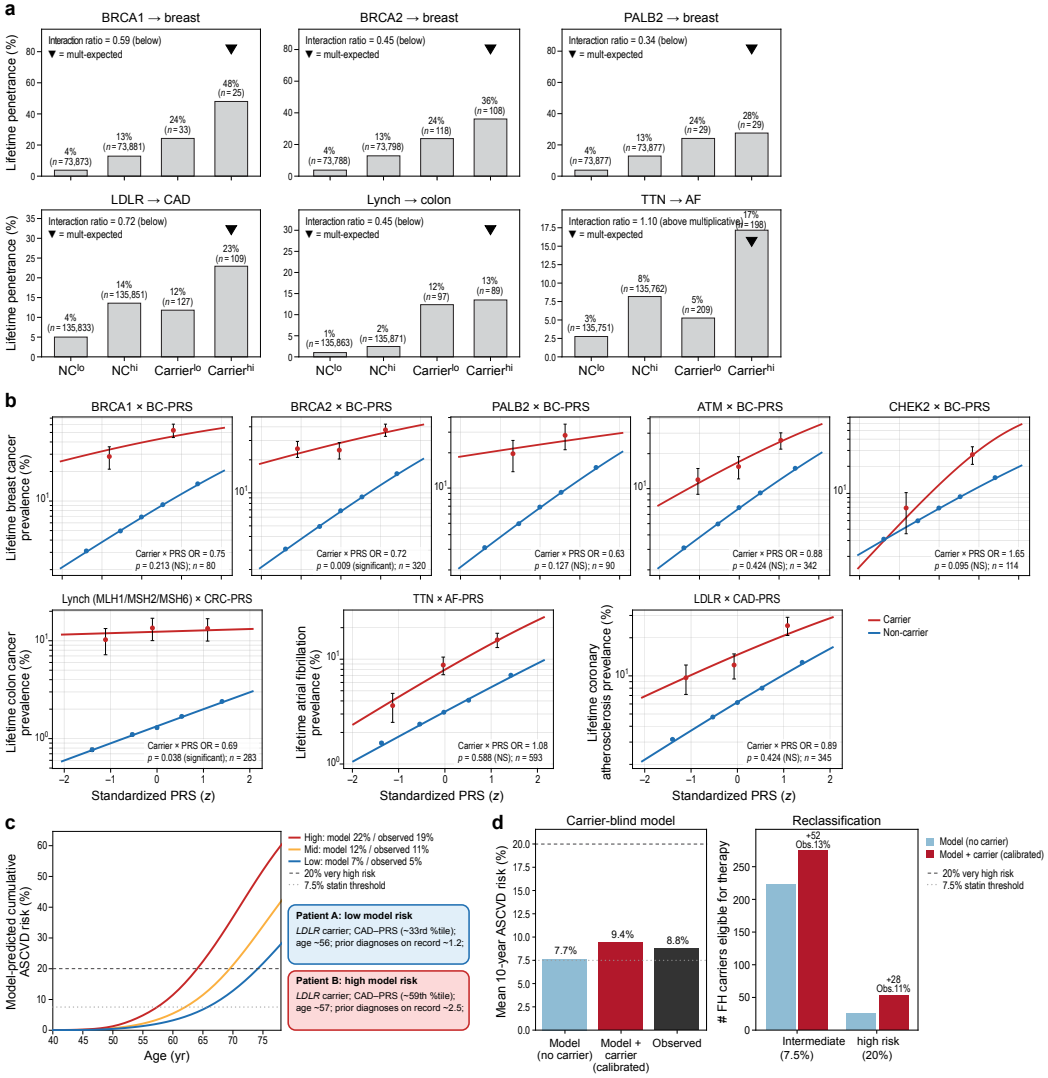
Carrier status, polygenic scores and the health record combined. *(a)* Observed lifetime risk in carriers and non-carriers by polygenic score, against the risk expected if the two acted multiplicatively. *(b)* Interaction odds ratio per standard deviation of score (logistic regression adjusted for age, sex and ten principal components) for eight gene–disease pairs. For moderate-effect genes the score has the same effect in carriers and non-carriers (*TTN*, odds ratio 1.08, *p* = 0.59); for high-penetrance cancer genes it has a smaller relative effect in carriers (*BRCA2*, 0.72, 95% CI 0.57–0.92; Lynch genes, 0.69, 0.49–0.98; not adjusted for multiple testing), although it may still add more absolute risk in carriers. *(c) LDLR* carriers ranked by predicted risk within five-year age groups and divided into thirds: observed ten-year ASCVD risk 5%, 11% and 19% (Kaplan–Meier; log-rank test for trend *p* = 0.002) against predicted 7%, 12% and 22%. The thirds differ in polygenic score as well as in recorded history. *(d)* Ten-year ASCVD risk in familial hypercholesterolemia carriers predicted without carrier status (7.7%, after recalibration to the observed population rate), that prediction scaled by the carriers’ observed rate ratio (9.4%), and observed (8.8%); 52 additional carriers cross the 7.5% statin threshold, with observed ten-year risk 13%.

We then asked whether the model’s predictions, from polygenic score and health record, separate carriers of the same gene by risk. We ranked *LDLR* carriers by the model’s predicted risk within five-year age bands and divided them into thirds. The observed ten-year ASCVD risk was 5%, 11% and 19% in the three groups (log-rank test for trend, *p* = 0.002), close to the model’s predictions of 7%, 12% and 22% (Fig. 5c).

Finally, because polygenic scores do not stand in for carrier status, a model without the carrier term should underestimate carriers’ risk. Across the 34 genes of the atlas, the model without the carrier term under-predicted carriers’ risk of the gene’s main disease (median observed/expected 2.2; Supp. Table S9). Without the carrier term, the model predicted a ten-year ASCVD risk of 7.7% for familial hypercholesterolemia carriers, against an observed 8.8%; scaling this prediction by the carriers’ observed rate ratio (1.23) gave 9.4% (Fig. 5d). This scaling moved 52 additional carriers above the 7.5% threshold for statin therapy, and their observed ten-year ASCVD risk was 13%.

### The disease selection replicates in two independent cohorts

Replication requires that carriers can be identified comparably across cohorts. We applied the same carrier definition to All of Us [33, 34] (170,825 participants with genome sequences) and the Mass General Brigham Biobank (MGB; 53,111 with exome sequences; Table 1). Carrier rates in UK Biobank and MGB agreed within two-fold for 39 of 61 genes (median ratio 1.19). Across all three cohorts, the largest discrepancies were in genes with few UK Biobank carriers, such as *PKD1*, which has six pseudogenes (17.6-fold higher in All of Us than in UK Biobank), and *INS* (14.3-fold). A sequence-quality filter in the UK Biobank exome release removes recurrent pathogenic alleles in *PMS2, CHEK2* and *BRCA1* (Fig. S12a).

**Table 1:** The three cohorts. Carriers can only be ascertained in the sequenced subset, so the carrier denominator is *N sequenced*, not the model cohort: UK Biobank’s model cohort holds 407,878 participants but the ClinVar extraction ran on the 200K exome release, and All of Us’s data set holds 243,303 people of whom 170,825 are genotyped with recorded sex. Variant counts are defined differently across cohorts: UK Biobank and MGB report panel positions *queried*, All of Us reports distinct panel variants *observed*, so the two are not compared directly. **End of follow-up** is the maximum age at which a participant is still at risk (max_*d*_ *E*_*id*_), computed from the diagnosis data, and it limits which diagnoses can be observed. Because it spans ≈ 17 years across the three cohorts, absolute ages are not comparable between cohorts, so ages are compared within each cohort (Fig. 6). MGB is a consented cohort, and its analogue of an assessment-center enrollment age is age at Biobank consent: median 55.8 (IQR 40–67), available for all 53,111 participants. Its interquartile range spans 27 years against UK Biobank’s 13, so the cohorts differ in the *heterogeneity* of entry as much as in its central value. *N (model cohort)* is the set the model was fitted on; for All of Us that is the analysis set (170,825), not the full data set. *Clean* genes are the 48 of the panel that are not driven by a single high-frequency allele; the common-allele genes are excluded because their carriers are not interpretable as monogenic (per-gene strata in Supp. Table S4).

|  | UK Biobank | MGB | All of Us |
|---|---|---|---|
| *N* (model cohort) | 407,878 | 53,111 | 170,825 |
| *N* sequenced | 200,343 | 53,111 | 170,825 |
| Assay | WES (rLCR_SEGDUP-filtered) | WES | WGS |
| ClinVar P/LP variants | 58,197 queried | 58,197 queried | 5,693 observed |
| Carriers (48 clean genes) | 8,264 | 2,795 | 8,914 |
| Carrier rate (%) | 4.12 | 5.26 | 5.22 |
| Age at entry, median (IQR) | 59 (51–64) | 55.8 (40–67) | 54 (38–65) |
| End of follow-up, median (IQR) | 73 (66–78) | 63 (47–74) | 56 (40–68) |

**Table 2:** Carriers and people at matched polygenic risk develop different diseases (model-free). For each gene, carriers are compared with the group at the top of the polygenic score for the gene’s primary disease whose excess of that disease is closest to the carriers’. Excess is the difference in the proportion diagnosed by age 70 between the group and everyone else in the same sex-restricted population (percentage points; 95% bootstrap CI, 300 resamples). No polygenic group reached the colon cancer excess of Lynch carriers; the last column gives the secondary excess per percentage point of primary excess.

| Genes | Primary disease | Secondary disease | Group | Primary excess (pp) | Secondary excess (pp) | Secondary per primary |
|---|---|---|---|---|---|---|
| *BRCA1*, *BRCA2*, *PALB2* | breast cancer | ovarian cancer | carriers (*n* = 490) | 21.6 (17.8, 25.5) | 6.5 (4.2, 8.6) | 0.30 |
|  |  |  | breast cancer score, top 0.5% (*n* = 1,107) | 19.7 (17.5, 22.1) | 0.0 (−0.4, 0.5) | 0.00 |
| *TTN* | atrial fibrillation | cardiomyopathy | carriers (*n* = 593) | 5.5 (3.2, 8.2) | 3.8 (2.3, 5.5) | 0.68 |
|  |  |  | atrial fibrillation score, top 10% (*n* = 40,729) | 6.1 (5.8, 6.4) | 0.3 (0.2, 0.4) | 0.04 |
| *MLH1*, *MSH6* | colon cancer | uterine cancer | carriers (*n* = 259) | 9.7 (6.6, 13.8) | 6.9 (4.0, 10.5) | 0.70 |
|  |  |  | colorectal cancer score, top 0.5% (*n* = 2,039) | 2.6 (1.9, 3.5) | −0.2 (−0.4, 0.0) | −0.09 |

We then asked whether the diseases selected for each gene in UK Biobank are also selected when the model is fitted separately in All of Us. All of Us recovered a median of 31% of each gene’s UK Biobank selections and two of its three strongest (67%), against 13% expected for a random selection of the same size; for 20 of 26 genes the overlap exceeded chance (Fig. S12c). In MGB, the diseases selected in both cohorts distinguished carriers from matched non-carriers in the observed data (Fig. S13). The diseases linked to each gene, therefore, replicate in independent health systems.

Finally, we asked whether carriers develop these diseases earlier than non-carriers, using only observed diagnoses. For each selected disease, we compared the median age at first diagnosis in carriers with that in matched non-carriers. Carriers were diagnosed a median of 3.0 years earlier across 34 gene–disease pairs (earlier for 26, later for 5; sign test *p* = 2 *×* 10^−4^); for example, *BRCA1* carriers were diagnosed with breast cancer 8.5 years earlier (Fig. 6b). The order in which carriers of each gene are diagnosed with these diseases is shown in Fig. 6a. Because follow-up differs between cohorts, we compare ages within each cohort only. Some later diagnoses, such as osteoporosis after endocrine therapy, likely reflect treatment, so we report the carrier–non-carrier difference rather than absolute ages.

**Figure 6:**
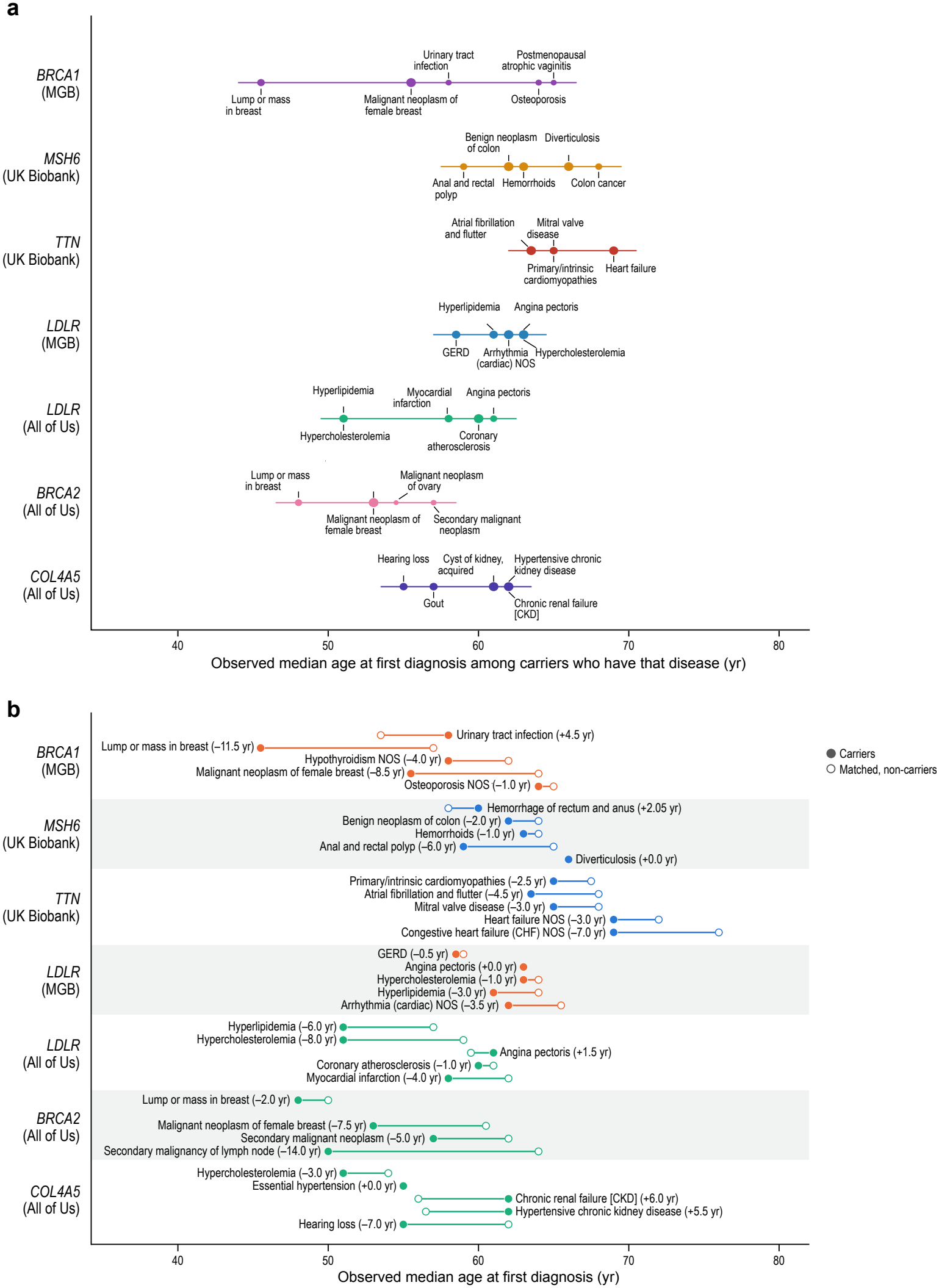
Carrier life courses in three cohorts. *(a) Order of diagnoses*. For each gene and cohort, each marker is a disease selected for that gene. Its position is the median age at which carriers in that cohort are first diagnosed with it, and its size is the fitted carrier effect. Some diagnoses reflect treatment (for example osteoporosis after endocrine therapy in *BRCA1* carriers). We compare the order of diagnoses within each gene and cohort, not ages across cohorts, because ages depend on each cohort’s follow-up. *LDLR* carriers in All of Us reach myocardial infarction and coronary atherosclerosis, which are not selected in MGB, possibly because treatment differs between cohorts. *(b) Comparison with matched non-carriers*. For each disease, a segment runs from the median age at first diagnosis in non-carriers matched 2:1 on sex and 5-year age group (open circle) to the carrier median (filled circle); the number in parentheses is the carrier minus non-carrier difference in years (negative, earlier in carriers). Because shorter follow-up censors carriers and controls alike, this difference is less sensitive to follow-up than the absolute ages. Diseases are shown when the matched non-carriers have enough diagnoses to estimate a median; 7 of 163 are lost this way (0*/*22 in MGB, 2*/*22 in UK Biobank and 5*/*119 in All of Us).

## Discussion

Common and rare variants are often summarized by their relative risk for a single disease. Our work shows that they shape disease over the life course and across diseases in different ways. A polygenic score shifts an individual’s loading on a signature, scaling the probabilities of all its diseases together, and this effect changes with age. A pathogenic variant instead changes the probability of specific diseases within a signature, and when they occur. Both effects are learned from the health record across 348 diseases, without prespecifying which diseases a gene affects or how a score’s effect changes with age.

The clearest evidence comes from the recorded diagnoses, without the model. Polygenic scores can identify people whose risk of a disease equals that of monogenic carriers [18], but equal risk of the primary disease does not mean the same disease course. At matched breast cancer risk, carriers of pathogenic variants developed ovarian cancer, while high-score individuals did not. The model represents this difference with two terms: one that scales a signature and one that modifies its composition. These describe patterns of disease rather than molecular mechanisms.

Estimating carrier effects by disease and age reproduces established biology without being told which diseases each gene affects. For most genes, the largest inferred effect aligned with the gene’s known disease. Cardiac genes sharing the same signature selected distinct cardiac conditions, and these selected diseases were consistent when the model was fit independently in All of Us and MGB. Letting effects change with age also captures when they occur. The effect of *BRCA1* on breast cancer peaks almost two decades earlier than that of *BRCA2*, matching the observed diagnosis patterns, and carriers received diagnoses of their selected diseases a median of three years earlier than matched non-carriers.

Disease trajectories have been described from population registries and health records [35, 22, 23], and health record phenotypes can reveal unrecognized Mendelian disease [36]; here, the trajectory model itself separates the effects of common and rare variation and estimates when each acts. Evidence that polygenic effects weaken with age has been reported for several common diseases [3, 4, 5]. Here, the age dependence is estimated jointly across diseases through the signatures that link them, rather than being assumed. These effects declined with age for most scores but increased for some, and modeling this age dependence improved held-out discrimination for all 28 diseases tested and in 32 of 36 disease–prior-history groups, with the largest gain in the identification of future cases among people with no prior diagnoses. Separating how much disease a person accrues from which diseases, through a health component, per-person frailty, and ancestry adjustment of the scores improved the calibration of absolute risk.

These results bear on how genetic information is used in prevention. Because a polygenic score carries most of its information early, it is most useful before diagnoses accumulate. Because carrier effects have gene-specific timing, surveillance ages could follow the age at which each variant’s effect rises, rather than a single lifetime risk. Genotype alone does not determine who will develop the disease: among *LDLR* carriers of the same age, the model combining the health record with polygenic scores separated carriers with observed ten-year ASCVD risks of 5% and 19%, while a model without the carrier term underestimated their risk and would have left 52 carriers below the statin threshold. Carrier status and polygenic score combined roughly multiplicatively for moderate-effect genes. For high-penetrance cancer genes, each unit of polygenic score raised the relative risk less in carriers, as reported for breast cancer in *BRCA* carriers [37], although it may still raise their absolute risk more [17].

This study has limitations. Electronic health records capture diagnoses when they are recorded, not when the disease begins. Some diseases in carrier trajectories reflect treatment rather than the variant itself. Absolute penetrance is calibrated to population registries; calibration to family-based estimates would require explicit correction for ascertainment. Penetrance estimated in population cohorts is generally lower and more variable than in clinically ascertained families [12, 14, 13], and depends on polygenic background [17]. Future work on joint modeling of laboratory measurements could link these trajectories to intermediate phenotypes and mechanisms.

A genetic result is more informative when read alongside age and the clinical record. A polygenic score is most informative early in life; a pathogenic variant indicates which diseases to expect and when; and the record shows which individuals are already progressing toward them.

## Methods

### Model

#### Relation to Aladynoulli

We develop a model that builds on the Aladynoulli model [21]. An individual’s recorded history is the binary array *Y*_*idt*_, which is 1 at the age disease *d* is first recorded for individual *i* and 0 otherwise; *E*_*id*_ is the last age at which *i* is at risk for *d*, the age at that first record or at censoring, whichever comes first. The model gives the probability of *Y* as a *mixture of probabilities* over *K* = 20 latent signatures of related illnesses together with a *healthy reference*: one further component, in which the probability of every disease is systematically lower than the population baseline [21]; an individual whose record is sparse relative to their age loads mainly on it instead of on a disease signature. The index *k* runs over all *K*+1 components. For individual *i*, disease *d* and age *t* ≤ *E*_*id*_,

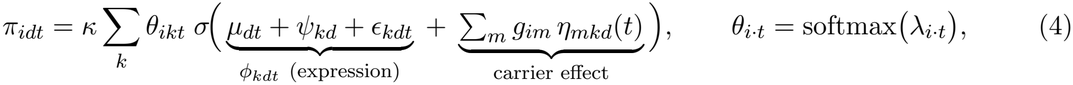

and the loading is defined by

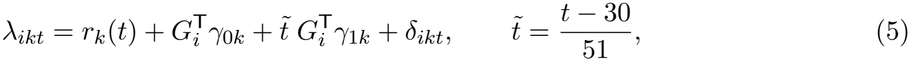

for every component. For the healthy reference the loading has one further term, a fixed per-person level *α*_*i*_ (*Health component*, below). Genetics enters *π*_*idt*_ at two points. Polygenic scores *G*_*i*_ act on the loadings *λ*_*ikt*_, and so on the mixture weights *θ*_*i·t*_ (eq. (5)), through a level effect *γ*_0*k*_ and an age interaction *γ*_1*k*_. A carrier indicator *g*_*im*_ acts on the signature-specific log-odds *ϕ*_*kdt*_ through the carrier effect *η*_*mkd*_(*t*), inside the sigmoid in eq. (4); *ϕ*_*kdt*_ itself is held fixed while the carrier effect is fit. Two terms are new here relative to ref. [21]: the age interaction *γ*_1*k*_ in eq. (5) and the carrier effect *η*_*mkd*_(*t*) in eq. (4). In simulation, the age interaction is recovered from data of this size (Supp. Fig. S10). Three further changes concern how the model represents overall health: the healthy reference follows an age curve (*Health component* ), ancestry principal components are excluded from *G*_*i*_ (*Polygenic scores and analysis cohort* ), and each individual has a frailty *f*_*i*_ (*Frailty* ). Table S1 lists every term, its size, and whether it is data, fixed before fitting, or learned.

#### Rescaling and reshaping

The two genetic terms change disease risk in different ways. *(i) The loading multiplies a signature’s disease probabilities by a common factor*. A polygenic effect shifts *λ*_*ik*_ and, through the softmax, the whole weight vector *θ*_*i·*_; within any one signature, *θ*_*ik*_ multiplies *σ*(*ϕ*_*kdt*_) for every disease *d* by the same factor, so that signature’s contribution 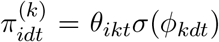 changes by that factor for all *d* and every within-signature ratio 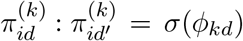 : 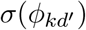 is unchanged. The diseases whose probabilities move are the few the signature expresses: *σ*(*ϕ*_*k·*_) is concentrated there and near zero elsewhere. Which diseases those are is set by *ϕ* and does not depend on *θ*_*ik*_. *(ii) The carrier effect adds to the expression of particular diseases*. It moves *ϕ*_*kd*_ → *ϕ*_*kd*_ + *η*_*mkd*_ for specific *d*, so those ratios change. Only *η* can therefore raise one disease’s probability relative to the other diseases in its signature (for example, ovarian cancer in *BRCA* carriers); a change in the loading moves weight between signatures instead. By construction, a polygenic score, which enters *λ*, can only change the level at which a signature is expressed, whereas a carrier indicator, which enters *η*, can change which diseases within that signature are elevated, including one the signature did not previously express. The Results compare the two directly in the recorded diagnoses, without the model (Table 2; Supp. Fig. S2).

### Parameterizing the carrier effect: Learned sparsity and off-signature specificity

Each gene’s carrier effect may raise only a few diseases, chosen by the data, within one assigned signature, and may change with age. We write *η*_*mkd*_(*t*) = scope_*mk*_ *z*_*mkd*_ *v*_*mkd*_(*t*) (Table S1; Supp. Fig. S5). The assigned signature is fixed before fitting; the selection indicator *z*_*mkd*_ and the magnitude *v*_*mkd*_ are learned, and together they give *η*_*mkd*_ a spike-and-slab prior: a point mass at zero, the *spike*, on gene–disease pairs that are not selected, and a diffuse Gaussian component, the *slab*, on gene–disease pairs that are selected.

- *Assigned signature. K*_*m*_ = *{k* : scope_*mk*_ = 1*}*, usually a single signature; the rule and its one refinement are given below under *Assigning each gene to a signature*. Because *π*_*idt*_ contains *θ*_*ikt*_ *σ*(*·*), a carrier effect on signature *k* moves *π*_*idt*_ in proportion to *θ*_*ikt*_: if *θ*_*ik*_ ≈ 0 the change is near zero however large *η* is. The bound this places on attainable penetrance is derived in Supplementary Note 1, *The ceiling set by the loading*.
- *Selection indicator z*_*mkd*_: *which diseases*. Within the assigned signature, *z*_*mkd*_ is the inclusion indicator, selected with probability *ω*_*mkd*_ = Pr(*z*_*mkd*_ *>* 0), which is estimated (*Selecting diseases*, below); its prior is described below.
- *Magnitude v*_*mkd*_: *how large*. On a selected gene–disease pair, the slab is *N*(0, *τ* ^2^). The prior variance applies to the carrier effect; the population log-odds *µ*_*dt*_ is fixed throughout. In the age-constant fits *v*_*mkd*_ is one number per gene–disease pair.
- *Magnitude as a function of age, v*_*mkd*_(*t*). Where the carrier effect varies with age, *v*_*mkd*_(*·*) ∼ *GP*(0, *τ* ^2^*K*_*η*_), the carrier analogue of *ϵ*_*kd*_(*·*). Since the expression *ϕ*_*kdt*_ is the log-odds of disease *d* at age *t* within signature *k*, and carriers of gene *m* enter eq. (4) as *ϕ*_*kdt*_ + *η*_*mkd*_(*t*), *η*_*mkd*_(*t*) is the difference in that expression between carriers and non-carriers at the same disease, signature and age. For the onset curve we take *K*_*η*_ to be the second-difference (cubic-smoothing-spline) kernel, whose precision matrix is banded and costs *O*(*T* ); *ϵ* uses the radial basis function (RBF) kernel (Supp. Fig. S8).

The prior has three parts: independent Bernoulli inclusion indicators, each with prior probability *q*_0_; a *N*(0, *τ* ^2^) slab on the magnitude; and, where the magnitude varies with age, the second difference kernel, whose density adds a term in 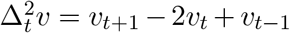, the local second difference, weighted by *ρ*. A Bernoulli(*q*_0_) prior adds log*{*(1 − *q*_0_)*/q*_0_*}* to the negative log prior for every selected gene–disease pair, so it is equivalent to a penalty *ν*_0_ per selected gene–disease pair with *q*_0_ = *σ*(−*ν*_0_); we set *ν*_0_ = 0.5, giving *q*_0_ = 0.38. The objective these enter is given below under *Objective for the carrier effects*. We use the second difference rather than the first because the two shrink *v* towards different things. A first-difference prior shrinks *v* towards a constant in age (that is, towards the age-constant carrier effect) and so would damp the age dependence we aim to estimate. The second-difference prior shrinks *v* towards a straight line: every straight line in age has 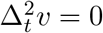 and contributes nothing, so a steady age trend of any slope is left alone and only local curvature is damped. The spike controls how many gene–disease pairs are selected and the slab the magnitude of each, so the model can select few carrier effects and still allow each to be large.

#### Health and overall disease burden

Two further terms separate how much disease a person has from which diseases. Without them, differences in overall disease burden are absorbed by the signature loadings and genetic effects.

#### Health component

The mixture weights *θ*_*i·t*_ = softmax(*λ*_*i·t*_) are invariant to a common perperson, per-time shift *λ*_*ikt*_ → *λ*_*ikt*_ + *c*_*it*_: it leaves every *θ* and every *π*_*idt*_ unchanged, and so is not identified by the data. Two such shifts matter here. One constant in age, 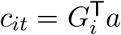, is *γ*_0*k*_ → *γ*_0*k*_ +*a* for every *k*; one proportional to age, 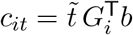, is *γ*_1*k*_ → *γ*_1*k*_ + *b* for every *k*. We hold the healthy reference’s genetic level fixed rather than re-estimating it in the population fit,

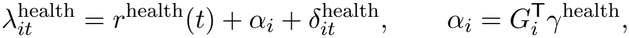

where *r*^health^(*t*) is the population disease-free log-odds curve, tempered towards the mean of the *K* disease references by a factor 0.6 and shifted to mean 1.5, so that the healthy reference is strongest when young, fades with age, and stays above the most-loaded disease signature through mid-life. The coefficient *γ*^health^ is estimated in the first fitting step (Table S2) and *α*_*i*_ is held fixed thereafter; *δ*^health^ carries the same Gaussian-process prior as the *K* signatures. The two shifts are then removed in different ways. An age-constant *a* cannot be applied to a component whose genetic level is held fixed, so the level effects *γ*_0*k*_ are determined in absolute terms against an estimated reference. The healthy reference carries no term in 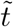, which sets *γ*_1,*K*_ ≡ 0 and forces *b* = 0, so the age interactions *γ*_1*k*_ are identified relative to a healthy reference assumed to carry no genetic age trend. The predictions do not depend on this choice; it fixes only the scale of *γ*_1*k*_.

A recovery simulation (Supp. Fig. S11) assesses the effect of the level anchor. It fixes *α*_*i*_ but leaves the healthy reference’s age interaction free, so there the age-proportional direction is restrained only by the Gaussian-process prior on *δ*. Without the anchor the estimates follow the *relative* slopes (*r* = 0.996; mean absolute error 0.0063 against those relative targets); with it they follow the *absolute* slopes (*r* = 0.974; mean absolute error 0.0041 against true slopes of 0.01– 0.05). Correlation is unchanged by affine rescaling of the estimates, so the error magnitudes, not *r*, distinguish absolute from relative recovery; by this measure the anchored fit recovers the absolute scale.

#### Frailty

We add a per-person frailty, which allows individuals to differ in their overall rate of disease without changing which diseases they have. Because *θ*_*i·t*_ is a composition, nothing in *λ*_*i·t*_ can express *how much* disease an individual has, only *which* diseases: two people with the same mix of diagnoses, one with twice as many, receive the same *θ*. Without a separate rate term, a difference in overall event rate can be represented only by moving weight onto whichever components happen to carry higher (or lower) disease probabilities, so that composition absorbs rate. We therefore give each individual a single frailty *f*_*i*_, applied to the mixture probability after the sum over components:

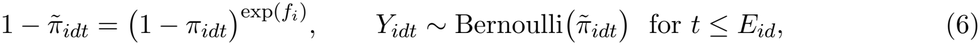

with *π*_*idt*_ as in eq. (4). Equivalently,

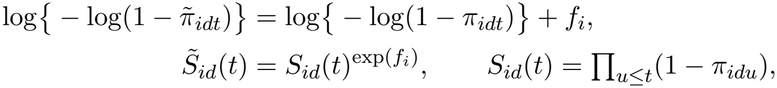

so *f*_*i*_ is an additive intercept on the complementary log–log scale and exp(*f*_*i*_) multiplies the individual’s cumulative hazard for every disease at every age; for small *π*_*idt*_, 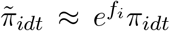. The complementary form keeps 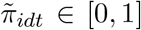 without truncation. Because *f*_*i*_ enters after the mixture, *∂θ*_*i·t*_*/∂f*_*i*_ = 0: it changes an individual’s overall rate but not their signature composition, and it carries no dependence on age or disease. *θ* describes the pattern of an individual’s disease and its evolution with age, and *f*_*i*_ its overall level.

A common shift *f*_*i*_ → *f*_*i*_ + *c* for all *i* is indistinguishable from raising the baseline *µ*_*dt*_ for every disease and age, so only differences between individuals are identified. We remove the common shift by centering, 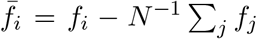, in the forward pass, and place a Gaussian prior on the centered values. The objective is the negative log-likelihood of eq. (6) plus

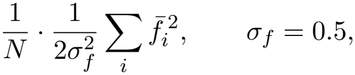

scaled by 1*/N*, as is the data term. The frailties are initialized at zero and are not warm-started from any earlier fit. In the model with the age interaction they are learned throughout the population fit, including the phase in which *δ* is held fixed, so that an individual’s overall rate is not absorbed by the genetic age interactions *γ*_1*k*_ estimated in that phase. Their learning rate is reduced tenfold once *δ* is released, so that *δ*, with *N ×*(*K*+1) *× T* parameters, cannot absorb the rate that *f*_*i*_ carries. The level-only model has no phase with *δ* held fixed and learns *f*_*i*_ at a single rate throughout. The frailties are then held fixed while the carrier effects are estimated. The fitted spread is 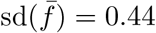 for the age-interaction model and 0.53 for the level-only model, against the prior *σ*_*f*_ = 0.5.

#### Assigning each gene to a signature

Each gene’s carrier effect is restricted to *K*_*m*_ (*Parameterizing the carrier effect*, above), typically a single signature. By default, the assigned signature is arg max_*k*_ *ψ*_*kd*_ at the gene’s canonical disease *d* (*Disease outcomes*, below), the signature that expresses it. Only the choice of signature depends on that one disease; several diseases may then be selected within it. The default suffices when the question is whether a carrier effect is present at all, because the estimated penetrance does not depend on which signature the carrier effect acts through (Supplementary Note 1). For the externally calibrated cancers, where absolute magnitude matters, the assigned signature is instead 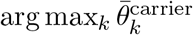, the signature on which carriers load most. All atlas genes use the default rule, including the eight breast-panel genes, which are assigned to the breast-expressing S19. We also fit *BRCA1, BRCA2* and *MSH6* with the carrier effect on S8, the signature that holds most gynecological diagnoses, keeping every other setting the same (Fig. 3b).

### Study cohorts and disease outcomes in the UK Biobank

#### Disease outcomes

Disease outcomes are ICD-derived phecodes [38, 39]; cancer outcomes for calibration use the national cancer registry (ICD-10 C50 breast cancer; C18–C20 colorectal cancer). Each gene is assigned one *canonical disease* (the disease the gene is established to cause) by a fixed gene-to-disease map applied before any fit and matched to the phecode vocabulary by name (*LDLR* → hypercholesterolemia, *TTN* → heart failure, *KCNQ1* → atrial fibrillation, *BRCA1/2* → malignant neoplasm of female breast); the full map is Table S7. Of the 64 panel genes, 61 have at least one P/LP carrier in the UK Biobank callset (*STK11, APOC3* and *PROS1* have none), 42 carry a canonical-disease assignment, and those with at least 30 carriers were fit.

##### Two atlases, two gene sets

The age-varying *η*(*t*) atlas covers 34 genes (Table S8) and the age-constant *η* atlas 32 (Table S7); 30 are common to both. *KCNQ1, LMF1, LPL* and *SCN5A* appear only in the age-varying atlas, while *PKD1* and *HFE* appear only in the age-constant one, fit with gene-specific carrier definitions: *PKD1* assigned to the renal/urologic signature S12 with functional-mask rather than strict-ClinVar carriers, and *HFE* restricted to the 1,202 C282Y homozygotes and assigned to several signatures covering the hemochromatosis manifestations, because the broad P/LP set (28,260 carriers) is dominated by heterozygotes. Where the two atlases are compared, the comparison is restricted to the 30 shared genes.

For *LDLR, BRCA1/2, MLH1, TTN* and *DSP* the canonical disease is selected. Two common-variant genes are included for contrast rather than as monogenic hypotheses: *SERPINA1* (*N* = 25,827 P/LP carriers) and *HFE* (as above). Genes not fit had too few strict-ClinVar carriers (*PCSK9*, 3 carriers; *UMOD*, 1).

#### Polygenic scores and analysis cohort

We used 36 polygenic scores from the UK Biobank Polygenic Risk Score Release [29]. *G*_*i*_ holds the 36 polygenic scores of ref. [21], each residualized on the first 10 genetic principal components by one cohort-wide least-squares fit, and sex (*P* = 37), each standardized to mean 0 and unit SD. The principal components themselves are excluded from *G*_*i*_ so that ancestry cannot shift the healthy reference. The scores, their construction, and the ancestry-stratified validation of the loadings are as described there; all scores are calculated using GWAS that exclude UKB and are externally validated [29]. Because the scores are residualized on the principal components, the fitted loadings are adjusted for ancestry, whereas the empirical per-SD hazard ratios below are not (*Polygenic effects by age*, below). *N* ≈ 400,000 for the polygenic analyses; the carrier analyses use the 200K exome subset.

#### Carriers

We used a list of 64 genes fixed before any fitting: monogenic disease genes, together with the common-variant negative controls named under *Disease outcomes* above. Carriers were identified in the UK Biobank [28] 200K whole-exome sequence release [31]. A variant counts only if ClinVar [30] records it as pathogenic or likely pathogenic (P/LP) at that exact coordinate, so a variant is never counted on its predicted consequence or its rarity alone, as it would be in a gene-level rare-variant burden test [40]. We took all ClinVar P/LP variants in the panel genes (57,765 positions for UK Biobank; MGB was queried against the 2026-08-04 release, 58,197 positions across the same 64 genes; Table 1) and matched exome variants to them by chromosome, position, reference and alternate allele (Supplementary Note 1). Anyone with at least one P/LP allele at a matched site is a carrier. This gives carrier lists for 61 of the 64 genes; the other three (*STK11, APOC3* and *PROS1* ) have no carrier at a matched site in the UK Biobank exome callset, which the cross-cohort comparison attributes to a sequence-quality filter rather than to absence in the population (Fig. S12a). For two genes the P/LP set is narrowed to the variant class or genotype established as pathogenic. For *TTN*, truncating variants are restricted to exons highly expressed in heart (hiPSI); this recovers the heart-failure and atrial-fibrillation signal. The carrier−control difference in cumulative incidence at age 70 rises from −0.4 (heart failure) and −0.6 (atrial fibrillation) percentage points under an allele-frequency mask (MAF *<* 10^−4^, *n* = 1,673), to +4.3 and +4.9 under ClinVar P/LP (*n* = 593), and to +5.8 and +7.0 with the hiPSI restriction (*n* = 369). For *HFE*, we used C282Y homozygotes, the penetrant genotype. Carrier counts agreed with an independent functional mask (*BRCA2* : 664 carriers called from the exome sequences against 649 under the mask).

### Model fitting

#### Overview

All learned parameters (Table S1) are point estimates, obtained by gradient-based optimization of the log likelihood plus the log prior; fits in 40 subsets of the data give confidence intervals for the parameter estimates. Fitting has two steps. First, the population model (signatures, polygenic effects, the health component and each individual’s frailty) is fitted to all 400,000 UK Biobank participants. Second, carrier effects are fitted to carriers and matched non-carriers with the population model held fixed, so that the small carrier samples cannot alter the signatures shared by the whole cohort. This is a modular, or ‘cut’, analysis [41, 42]: the carrier estimates do not carry the uncertainty of the population fit (Supplementary Note 1). Starting from a model without the age interaction, the full model is fitted in two phases. In the first, the per-person deviations *δ* are held at zero, so that age-dependent genetic signal is captured by the polygenic age effects *γ*_1_ rather than by individual deviations. In the second, *δ* is estimated as well, starting from zero, while the signature parameters are fine-tuned. Participants are fitted in 40 batches of 10,000. For prediction, population parameters are averaged over the other 39 batches and held fixed, and each held-out individual’s *δ* and frailty are refit from zero on that individual’s records up to the prediction age (200 optimization steps).

#### Objective for the carrier effects

The carrier effect is fit with the population parameters *{ψ, ϵ, γ*_0_, *γ*_1_*}* held fixed (Table S2), by minimizing

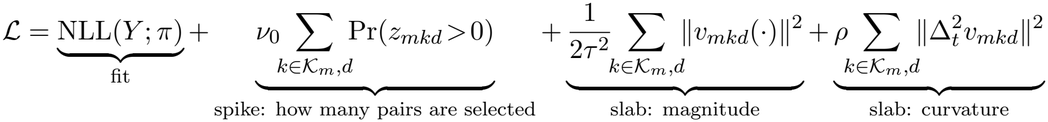

over *{l*_*mkd*_, *v*_*mkd*_(*·*), *δ}*, with all three sums running over *k* ∈ *K*_*m*_ and all *d*. The three terms beside the fit are the negative log priors set out under *Parameterizing the carrier effect* above.

#### Selecting diseases

Because the inclusion indicator *z*_*mkd*_ is binary, its contribution to the objective is not differentiable. We use the hard-concrete relaxation of Louizos et al. [43], which allows gradient-based fitting while still letting a gene–disease pair be switched off exactly; selection parameters start with almost every pair included (*ω*_*mkd*_ = 0.96), and the penalty *ν*_0_ removes those the data do not support (details in Supplementary Note 1).

#### Order of fitting for carrier effects

A carrier effect raises one disease within a signature, whereas a per-person deviation *δ* raises every disease in that signature; the two are distinguished by this contrast across diseases. For age-varying carrier effects we fit the carrier effects first, with *δ* held at zero, and then estimate both, because *δ* can change freely with age and could otherwise absorb part of an age-varying carrier effect. For age-constant carrier effects, which *δ* cannot mimic in this way, *δ* is estimated first. The same reasoning motivates holding *δ* at zero while the polygenic age effects are fitted. Optimization settings are given in Supplementary Note 1.

### Calibration of polygenic risk

#### Empirical PRS calibration

A recalibration *mapping* is a transformation applied to the predicted probability after fitting, leaving every model parameter unchanged. We use three; Table S11 lists them with the data each is fit on and its effect, and numbers them 1–3. Mapping 1 is described here. After this recalibration, observed and predicted rates agree within each score quintile (Supp. Fig. S4). To compare observed and predicted hazards within PRS strata, we write *π* = *σ*(*φ*) with *φ* the model logit. Events are rare, so log *π* ≈ *φ* and

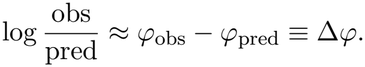

A discrepancy that is multiplicative on the probability scale is therefore additive on the logit. If Δ*φ* grows with PRS, the model’s PRS slope on the logit is too shallow (Supp. Fig. S4). Let *z* be the standardized PRS. The fit is carried out separately in each *window*, a 5-year age interval labeled by its starting age *a*; in each window we fit Δ*φ* as a quadratic in *z* and add it back on the logit:

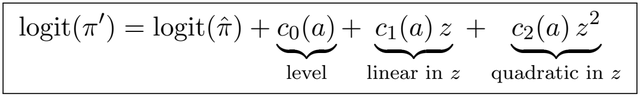

The *c*_*j*_ are fit to calibration alone: observed against predicted annual hazard across 20 PRS-percentile bins. The top-versus-bottom PRS-quintile event-rate ratio *Q*_5_*/Q*_1_ and the per-SD hazard ratio are held out of the fit. Because the two summaries average the data differently, *Q*_5_*/Q*_1_ over pooled person-years and the per-SD ratio over per-person at-risk hazards, the bin means are formed the same way as the summary they are compared with; fit on the mismatched basis, the T1D per-SD ratio falls to about half its empirical value at age 45 while *Q*_5_*/Q*_1_ is unaffected. The quadratic term is needed only for T1D, where Δ*φ* rises faster than linearly in *z*; for CAD and AF it is close to linear. Before that term is applied, *z* is clipped to the range spanned by the fitting data (any *z* outside that range is replaced by the nearer endpoint), so the correction is never extrapolated.

#### Per-window application

Each window has its own coefficient triple *c*_*j*_(*a*), *j* ∈ *{*0, 1, 2*}*. The correction used in Fig. 1b,c applies each window’s coefficients unsmoothed, which matches the within-stratum hazards more closely than the smoothed variant; a variant that smooths each co-efficient across age with a low-order polynomial in *a* (degree ≤ 2) and evaluates it at each year removes the step at window boundaries and serves as a continuity check. Fitting and application use the same per-year at-risk hazards. The correction is applied to the predictions after fitting, and no model parameter, including *δ*, is re-estimated.

#### Held-out refit of the calibration mapping

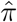is leave-one-out, so the predictions entering the recalibration are out-of-sample for each participant, and Fig. 1b,c show the full-cohort recalibration. The mapping itself is fit on the cohort to which it is applied, so we also refit the *c*_*j*_(*a*) on a random half and evaluate *Q*_5_*/Q*_1_ on the disjoint half (Supp. Fig. S3): with *N* ≈ 400,000 and low-degree per-window coefficients the two *Q*_5_*/Q*_1_ curves coincide at every age (type 1 diabetes: 9.3 to 2.2 in the held-out half and 9.8 to 2.1 in the fitting half, against 9.1 to 2.0 in the full cohort, Fig. 1c).

### Calibration of carrier penetrance to registries

#### Fitting to registry diagnoses

Hospital records capture cancer diagnoses incompletely, so for breast, colorectal and prostate cancer we estimated the carrier effect from diagnoses in the national cancer registry linked to UK Biobank (ICD-10 C50, C18–C20 and C61). For carriers and matched non-carriers, the cancer outcome was replaced by the registry diagnosis, and the age-varying carrier effect *η*_*mkd*_(*t*) was fitted to these events with the population parameters held fixed (*Order of fitting for carrier effects*). Carrier penetrance by age *a* is

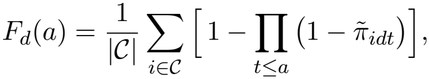

averaged over the carrier set *C*, and is compared with the registry Kaplan–Meier estimate for the same carriers, with Greenwood 95% confidence intervals (Fig. 3d). Because the carrier effect is estimated from these diagnoses, the comparison shows that the model can represent the registry penetrance curve and its shape with age. A carrier effect held constant in age cannot: when fit to match the registry at age 70, it over-predicts at age 60 (Supp. Fig. S6).

#### Interpreting penetrance

The carrier effect *η* is on the log-odds scale within the assigned signature, so the same *η* can mean very different changes in probability depending on the baseline: where a disease is rare in the assigned signature, a large *η* is a small change in risk. To compare genes we therefore report the carrier−control difference in cumulative incidence at age 70 (Δpen, percentage points), which is on the probability scale. How large an effect can be attained also depends on the assigned signature, because a carrier effect raises disease risk only in proportion to the carriers’ loading on that signature.

### Statistical analyses

#### Significance tests

Counts of concordant directions (disease–score pairs, diseases, disease–group combinations, gene–disease pairs) are tested against 0.5 with exact binomial (sign) tests. Paired per-person comparisons of predicted risk between the full and no-interaction models use the Wilcoxon signed-rank test, and the fraction above a risk threshold uses McNemar’s exact test; the comparison of risk changes between participants who did and did not develop disease uses the Mann–Whitney test. The confidence interval for the mean AUC gain treats the 28 diseases as units (*t* interval). Interaction odds ratios are reported with 95% confidence intervals, not adjusted for multiple testing.

#### Polygenic effects by age (Fig. 1)

For each disease and its polygenic score, standardized to mean 0 and unit standard deviation, we divided age into five-year windows. Within each window we kept participants at risk at the window start and recorded whether they had a first diagnosis during the window. The empirical per-SD hazard ratio in each window is from a regression of that outcome on the score (Supplementary Note 1). The model curves apply the same windows and at-risk sets to the held-out predicted probabilities, from the full model and from the model without the age interaction. As a check that does not depend on the regression, we also compared the event rate in the top and bottom score quintiles in each window (Fig. 1c; Supplementary Note 1).

#### Prediction and calibration (Fig. 2)

##### Participants with no prior diagnoses

Cases were participants with no diagnosis recorded before enrollment, no prior ASCVD, and a first ASCVD diagnosis within two years after enrollment (*n* = 1,610); non-cases had no prior diagnoses and never developed ASCVD. ASCVD risk was the probability of any of its five coronary component diseases in the enrollment year. We compared the two models’ predictions in cases with the Wilcoxon signed-rank test, the fraction above 10% per year with McNemar’s exact test, and the change in predicted risk between cases and non-cases with the Mann–Whitney test. *Cases identified at a fixed screening fraction*. For each disease, each model flagged the 5% of at-risk participants with the highest predicted one-year risk, so that both models flag the same number of people. Among participants diagnosed in the following year, we counted those flagged by only one of the two models, pooled across diseases with at least 20 cases and grouped by the number of diagnoses recorded before enrollment, and compared the two counts with McNemar’s exact test. The net gain is the difference between the two counts per 100 cases. *Calibration*. For held-out participants (the first 50,000), we pooled every person-year in which a participant was at risk for a disease and grouped them into 50 bins of predicted one-year risk, equally spaced on the log scale, keeping bins with at least 10,000 person-years, and compared mean predicted risk with the observed rate in each bin. The recalibration logit *π*^*′*^ = *α* + *β* logit *π* was estimated by logistic regression on a random half of the person-years and evaluated on the other half; because it is monotone, it does not change the AUC.

#### Carrier analyses (Figs. 3 and 4)

##### Positive control

Before fitting, each gene was assigned the disease used to choose its signature and a clinical keyword describing its known phenotype (for example, hypercholesterolemia for *LDLR*, colon cancer for *MLH1*, cardiomyopathy for *TTN* ). The target family for a gene was this disease, the diseases sharing its phecode to one decimal place, and any disease whose name contains the keyword. For the 30 ClinVar genes assigned to a single signature, we asked whether the model selected a target disease and whether a target was the strongest selected effect (largest carrier–control difference in cumulative incidence at age 70). Because each gene’s signature was chosen using its target disease, the main test is restricted to the diseases whose strongest signature is the gene’s own: we compared the number of genes whose strongest selected effect was a target disease with the number expected if the selected diseases were ordered at random (Poisson-binomial test).

###### Carriers and polygenic risk matched on the primary disease

For each gene, we compared carriers with groups at high polygenic risk for the gene’s primary disease (*BRCA1, BRCA2* and *PALB2* : breast cancer score; Lynch genes: colorectal cancer score; *TTN* : atrial fibrillation score), defined as the top 10%, 5%, 2%, 1% and 0.5% of the score. Each group was compared with everyone else in the same sex-restricted population, as carriers were with non-carriers, and the excess was the difference in the proportion diagnosed by age 70. We chose the polygenic group whose excess of the primary disease was closest to the carriers’ and compared the two groups’ excess of the gene’s secondary disease (ovarian cancer, uterine cancer and cardiomyopathy, respectively). Where no polygenic group matched, we compared the secondary excess per percentage point of primary excess. Confidence intervals are from 300 bootstrap resamples within each group.

###### Observed check of selected pairs

For each of the 307 selected gene–disease pairs, we compared the proportion of carriers ever diagnosed with the disease with that of non-carriers matched 1:4 on sex and five-year age group (two-sided Fisher exact test, Benjamini–Hochberg FDR across the 307 pairs). The check uses the same carriers as the selection, so an excess is a necessary condition not independent confirmation. With few carriers, a true effect may not reach significance.

### Clinical analyses in *LDLR* carriers (Fig. 5c,d)

#### Ten-year ASCVD risk

Predictions are the held-out predictions described under *Model fitting* : on each held-out batch the per-person deviation *δ* and the frailty *f*_*i*_ are estimated from the participant’s records up to enrollment, and risk is read forward from the enrollment age. ASCVD is any of seven phecodes: coronary and ischemic heart disease (unstable angina, myocardial infarction, angina, coronary atherosclerosis, other chronic and acute ischemic heart disease) and ischemic stroke (cerebral-artery occlusion with infarction). With the per-year ASCVD hazard *h*_*it*_ = 1 − ∏_*d*∈ASCVD_ (1 − *π*_*idt*_), the ten-year risk from enrollment age *e*_*i*_ is 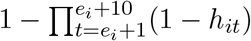.

#### Risk groups

Among the 309 *LDLR* carriers free of ASCVD at enrollment, we ranked predicted ten-year ASCVD risk within five-year groups of enrollment age and divided carriers into thirds, so that the three groups have the same age distribution. Observed ten-year risk was estimated by Kaplan–Meier, censoring at the end of follow-up, and compared across the thirds with the log-rank test for trend. The thirds differ in polygenic score as well as in recorded history (mean CAD score percentile 33, 49 and 59).

#### Reclassification

For familial hypercholesterolemia carriers (*LDLR, APOB, PCSK9* ), the risk without the carrier term is the held-out prediction from the model without the carrier term after recalibrating ten-year ASCVD risk to the observed population rate (one intercept, estimated in non-carriers). The risk with the carrier term is that prediction multiplied by the carriers’ observed rate ratio in UK Biobank (1.23, age- and sex-matched). Both are compared with the carriers’ observed ten-year risk, and with the 7.5% treatment threshold. Details in Supplementary Note 1.

### Replication in All of Us and MGB

#### Replication of selected diseases

Cross-cohort agreement is reported as recall, the fraction of the reference cohort’s selected gene–disease pairs the second cohort also selects, because All of Us selects more diseases per gene for reasons unrelated to agreement: partly greater power (3–5*×* the carriers), partly a spike component that is not *κ*-scaled. The Jaccard index would penalize the larger union even when every reference gene–disease pair was recovered. Chance is the recall a random selection of the same size would achieve, so selecting more diseases raises the baseline. We report the median enrichment, recall divided by chance, as the replication measure. Because recall also falls as the reference selection grows, we additionally report recall of each gene’s *k* strongest reference gene–disease pairs, ranked by the carrier−control penetrance difference |Δpen|, for *k* = 3 (the minimum reference selection size analysed) and *k* = 5; this fixes the reference size and so does not depend on how many gene–disease pairs either cohort selects. The carrier atlases were fit separately in each cohort.

#### Entry and censoring in the cross-cohort panels

Observed curves are right-censored only, because in these cohorts the only available entry age is the first recorded diagnosis, which would remove disease-free person-time and bias incidence upward. Carriers and matched controls enter and are censored identically, so the convention cancels in the carrier–control difference and in the ordering of diseases; the absolute level is not a lifetime penetrance estimate. Absolute penetrance is calibrated separately against the published external curves, which apply delayed entry (*Fitting to registry diagnoses*, above). MGB’s follow-up ends nearly a decade earlier than UK Biobank’s (median end-of-follow-up ≈ 56 All of Us, 63 MGB, 73 UK Biobank), so cross-cohort comparisons are made on the Kaplan–Meier scale within each gene and cohort, not across cohorts. For each gene and cohort, the selected diseases are ordered by the carriers’ median age at first diagnosis, so that the order of diagnoses, not their absolute ages, is compared.

## Ethics

This study analyzed coded or de-identified data from three biobanks under institutional review board (IRB) approvals at Mass General Brigham (MGB), in accordance with the relevant guidelines and regulations of each biobank. Analyses of Mass General Brigham Biobank data were performed under MGB IRB protocol 2018P001236; all participants provided written or electronic informed consent for research use of their biospecimens and health records. Analyses of UK Biobank data were performed under MGB IRB protocol 2021P002228, determined exempt; UK Biobank received ethical approval from the North West Multi-Centre Research Ethics Committee (REC reference 11/NW/0382), all participants provided informed consent at recruitment, and this work was conducted under UK Biobank application 7089. Analyses of All of Us data were performed under MGB IRB protocol 2020P001737, determined to be analysis of de-identified data not meeting the definition of human subjects research; the All of Us Research Program has ethical oversight through its central IRB, and all participants provided informed consent at enrollment.

## Data availability

UK Biobank individual-level data (genotypes, whole-exome sequences, and linked hospital/registry records) are available to approved researchers through the UK Biobank Access Management System (https://www.ukbiobank.ac.uk); this study was conducted under application 7089. All of Us data are available to registered researchers through the All of Us Researcher Workbench (https://www.researchallofus.org); Mass General Brigham Biobank data are available to investigators under an approved Mass General Brigham institutional review board protocol and cannot be shared publicly; ClinVar variant annotations are publicly available (https://www.ncbi.nlm.nih.gov/clinvar). Cancer registry diagnoses are part of the UK Biobank linked records. Aggregate results, fitted parameters, and the exact values underlying each figure are provided as Source Data.

## Code availability

All model, analysis, and figure-generating code, with notebooks that reproduce each main figure, is available at https://surbut.github.io/mendelnoulli_paper/. The model is implemented in PyTorch.

## Acknowledgements

This work was supported by the Burroughs Wellcome Fund Career Award for Medical Scientists, NIH/NHLBI K08 HL183784, and American Heart Association DA 25CDA1444806 and generous donations from the Wilcox Foundation. This research was conducted using the UK Biobank Resource under application 7089. We thank the participants of UK Biobank, All of Us and the Mass General Brigham Biobank. This research would not be possible without the partnership of the respective biobank participants.

## Author contributions

S.M.U. conceived and developed the model, implemented the pipeline, performed the analyses, and wrote the manuscript. P.N., A.G., and G.P. supervised the work and revised the manuscript. B.T. and H.R. contributed to ClinVar variant curation and discussion. S.K. and P.T.E. provided clinical guidance on the interpretation of results. W.H. assisted with data acquisition and edited the manuscript. All authors reviewed and approved the final manuscript.

## Supplementary Note 1: Fitting and implementation details

This note collects technical details of model fitting and interpretation that support the Methods.

### Implementation

All fits use Adam (PyTorch 2.5.1, Python 3.9.21) at a constant learning rate of 0.1. The age-constant carrier effect is fit *δ*-first: 80 epochs on *δ* alone, then 150 epochs on *{δ, v, l}* jointly. The age-varying carrier effect reverses the order, *η*-first: 150 epochs on *{v, l}* with *δ* held at zero, then 300 epochs with *δ* free; the fits to registry diagnoses (Fig. 3d) use 400 epochs in the first phase and 300 in the second. Carriers are compared with non-carriers matched on sex and 5-year age bin at 2:1 (Figs. 3, 6), except in the registry fits, which use 4:1; the model-free Lynch panel (Fig. 4d) uses 1:4. The population fit of eq. (5) runs 100 epochs with *δ* held at zero followed by 200 with *δ* free. Learning rates are 0.1 for *γ*_0_ throughout and for *δ* once it is freed; 0.1 for *γ*_1_ in the first phase and 0.01 in the second; and 0.01 (one tenth) for *ψ* and *ϵ* in both. The curvature weight is *ρ* = 30, fixed before fitting and not tuned. The stability refits (Fig. 4c) are ten runs identical to the main fit in every respect except the random seed.

### Modular (cut) estimation

In a *cut* (or modularized) analysis the parameters of one module are estimated from part of the data and conditioned on as if known, so that information cannot flow back from the second module into the first [41]. Here the population module (*ψ, ϵ, γ*_0_, *γ*_1_, the health component’s age curve and genetic effect, and each individual’s frailty *f*_*i*_) is estimated on the full cohort, and the carrier module (the selection indicator and the magnitude of *η*, with the per-person deviation *δ* re-estimated) is estimated with it fixed. The population module therefore reaches stage 3 only as a point value: nothing in the carrier sample returns to the population parameters or to *f*_*i*_, which is what makes this a cut, and nothing about them beyond the fitted values (their uncertainty, and their dependence on the cohort the folds were drawn from) reaches *η* (refs. [41, 42] discuss estimators of this kind). The stage-3 estimates are not the mode of a posterior for any single joint model. Equations (4) and (5) remain a complete generative specification, and every quantity we report (penetrance curves, carrier−control contrasts, predicted hazards) is obtained by evaluating them forward at the fitted values. The fit does not provide a posterior distribution for those values.

### Fitting the population model in two phases

Phase 1 holds *δ* at zero while *γ*_1_ is learned, so the age interaction is what fits the age-dependent genetic signal; Phase 2 then estimates *δ* as well, starting it at zero. We write *δ*_0_ for any fit in which *δ* starts at zero rather than from the no-interaction fit. Starting *δ* instead from the no-interaction fit (mean |*δ*| ≈ 0.175) lets *δ* fit the same age-varying signal that *γ*_1_ is there to estimate, and the identifiability simulation that validated the two-phase approach used a *δ* ≈ 0 start. The signature log-odds shifts (*ψ*), the Gaussian-process residual (*ϵ*) and the level effect (*γ*_0_) carry over from stage 1 (Table S2); they are fine-tuned rather than held fixed, so that the signature structure stays close to the stage-1 solution while *γ*_1_ is learned. The two-phase *γ*_1_ is more stable across folds and is the one used for interpretation. For prediction, the population parameters are averaged over the other 39 batches and held fixed, and *δ* and *f*_*i*_ are refit from zero for 200 Adam steps (learning rate 0.1), *f*_*i*_ under its prior. A leave-one-out prediction for an individual uses only that individual’s records up to the prediction age.

### Order of fitting for carrier effects

*δ* and *η* are distinguished only by how they act across diseases. Raising 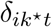 raises *π*_*idt*_ for every disease in signature *k*^⋆^, raising 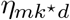 raises it for disease *d* alone, so at any one disease the two are interchangeable and what identifies *η* is the contrast between the selected disease and the other diseases in its signature. For an age-constant carrier effect we estimate *δ* first: Phase A estimates *δ* alone, with *η* and the selection indicator held at their initial values (*v* ≡ 0, and selection logits at 1.5, i.e. Pr(*z >* 0) = *σ*(1.5 + 1.599) ≈ 0.96; nearly every disease starts selected, so Phase A runs with the carrier effect switched on but zero-valued). For an age-varying carrier effect the order reverses: Phase A estimates *η*(*t*) and the selection indicator, Phase B adds *δ*. Every parameter is updated on every gradient step, so the phases decide only which are updated when, and the order matters because in Phase A *δ* is the only parameter that can reduce the carrier residual. It takes up as much of the excess at the selected disease as the rest of the likelihood allows (raising 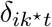 also raises the signature’s other diseases in the same carriers, where there is no excess to explain), and Phase B estimates *η* against what is left. Because the fit is stopped after a fixed number of epochs rather than run to convergence, how the excess divides depends on how far Phase A ran. For the age-constant carrier effect that is 80 epochs in Phase A and 150 in Phase B (*Implementation*, above); the corresponding counts for the age-varying carrier effect are 150 in Phase A, 300 in Phase B. At a joint optimum the across-disease contrast identifies *η* under either order, so this is a property of the optimization path and not of the model. We reverse the order for an age-varying carrier effect because *δ* mimics an age-varying *η*(*t*) more easily than a constant *η*. The deviation *δ* is estimated for carriers and controls alike in the age-constant Phase A too, but *δ*_*ikt*_ is free in age, so it can reproduce a smooth *η*(*t*) almost exactly, whereas a constant *η* has no age structure for *δ* to absorb and is pinned by the across-disease contrast alone. The asymmetry is in what *δ* can mimic, not in whom it is estimated for. Phase 1 of stage 2 holds *δ* at zero on the same argument: *δ* is held fixed whenever an age-varying genetic effect is being estimated, polygenic (*γ*_1_) or monogenic (*η*(*t*)).

### Gene–signature–disease combinations

Gene *m* is *assigned* to a set of signatures, those with scope_*mk*_ = 1, on which its carrier effect is allowed to act; within that set a per-disease *selection indicator z*_*mkd*_ decides which of those diseases the carrier effect applies to. Both are defined in full below. A triple (*m, k, d*) (one gene, one signature, one disease) with scope_*mk*_ = 1 is a *gene–disease pair*, the unit the selection indicator switches on or off, and the unit every count in the atlas refers to; triples outside a gene’s assigned signatures have *η*_*mkd*_ ≡ 0 and are not counted. A pair is *selected* when *z*_*mkd*_ *>* 0. Because a gene is usually assigned to one signature, its pairs share a *k* and differ in *d*; a gene fit under more than one assignment contributes pairs at each.

### Which signature a gene–disease pair is read on

Every selected gene–disease pair involves two signatures, and a model curve for the pair can be computed on either. The gene is assigned to *k*^⋆^, the signature its carriers load, and the carrier effect 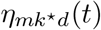 is indexed by *k*^⋆^. The disease *d* has its own *home signature*, home(*d*) = arg max_*k*_ *ψ*_*kd*_, the signature that expresses it. For an off-signature pair the two differ. We always compute on home(*d*), and because home(*d*) depends on the disease and not on the gene, the choice is made separately for each pair: one gene’s selection indicators can select diseases with different home signatures (*BRCA2* assigned to S8 selects diseases whose home signatures are S6, S16, S13, S3, S4 and S2), so the convention applies per pair. We keep home(*d*) even where the other signature agrees better with the observed curve. Where a disease is barely expressed in a signature (*ψ*_*kd*_ strongly negative, so the predicted baseline there is near zero, *σ*(*ϕ*) ≈ 0.001), almost any *η*(*t*) is consistent with the carrier data and the fitted curve can follow the observed hazard closely even though the data barely constrain it. In the All of Us fit (signature numbers are that cohort’s own), *LDLR*→hypercholesterolemia is the worked case: computed on S16 (*ψ* = −2.08) the fitted peak is 54 against an observed 49; computed on its home signature S14 (*ψ* = +0.28) it is 75. We report 75, which falls outside the bootstrap interval for the reason given below (*Multi-modal curves and the peak-age summary* ).

### Multi-modal curves and the peak-age summary

In the All of Us fit, *LDLR*→hypercholesterolem has its fitted onset peak at age 75 against an observed peak at 49, outside the bootstrap interval. Both curves are multi-modal, and they agree on where the modes are (model at ages 36, 54 and 75; data at 37, 49−56 and 71); they differ only in which mode is tallest, and a small change in relative amplitude moves the peak age by decades while the curve barely moves. Agreement between a fitted and an observed onset curve is therefore reported as *r*_*s*_, the Spearman rank correlation over ages 35–75, and not as agreement between peak ages; *r*_*s*_ is stable across 50 independent matched-control draws.

### The ceiling set by the loading

As 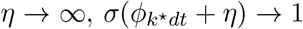, so the mixture probability rises to 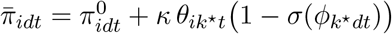, where 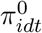 is the same individual’s mixture probability when *η*_*mkd*_ ≡ 0. The observed probability 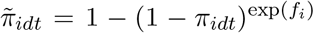 is increasing in *π*_*idt*_, so with the frailty *f*_*i*_ held at its fitted value,

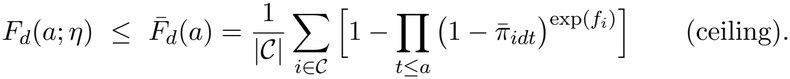

If 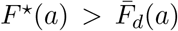, the external curve lies above the ceiling and no carrier effect reaches it, age-constant or age-varying. The limit is set by how much carriers load the assigned signature and is movable only through the loading: neither a larger *η* or *η*(*t*) nor a larger slab variance *τ* ^2^ raises it.

#### Derivation (chain rule)

Writing *p*(*z*) = E[*y* | *z*] for the conditional event probability, the per-SD effect on the log-odds scale follows from the chain *z* → *p* → logit *p*,

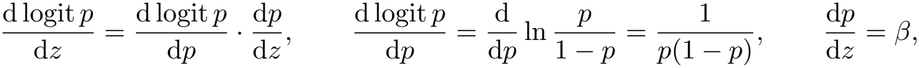

so that, evaluating the link derivative at the base rate 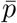,

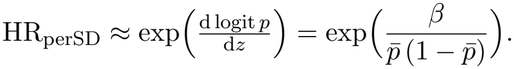

The two curves use the same computation and differ only in the regression target *y*_*i*_:

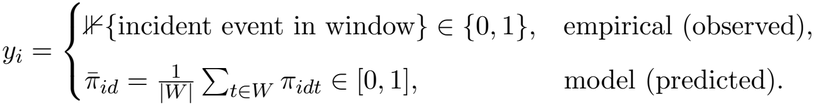

For the empirical curve *y* is binary and the fit is a linear-probability model, with 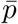 the observed event rate and 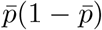 its Bernoulli variance; for the model curve 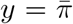 is already a probability, the fit is ordinary least squares of a continuous target, 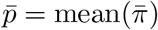, and 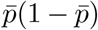 is the same logit link-derivative evaluated at the mean predicted probability rather than a binomial variance. Were the model to reproduce the genetic effect exactly, the two curves, and their decline with age, would coincide.

### Absolute vs. age-relative

The factor 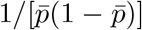grows without bound as 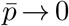, so at young ages, where events are rare, a small change in *β* moves HR_perSD_ a long way: the absolute empirical values there are noisy, while the model’s curve is smooth by construction. The model’s per-SD effect is also bounded by the *K*-signature representation it shares across diseases. We therefore compare the age-relative trajectory (each curve divided by its value at the youngest age, which removes any common factor), together with the direction of the age trend in the log hazard ratio, which the age-varying model matches in 10/10 disease–PRS pairs.

### Carrier extraction details

*Position matching*. Each ClinVar coordinate was matched by chromosome, position, reference and alternate allele against the exome variant list, and the matched variants were extracted with plink2 (v2.00a).

### Selecting diseases: the hard-concrete relaxation

The discreteness of the inclusion indicator obstructs gradient descent. Under the prior as stated, *z*_*mkd*_ is a Bernoulli draw taking the values 0 and 1, and so is a step function of the selection parameter *l*_*mkd*_: the likelihood is piecewise constant in *l*_*mkd*_, with derivative zero almost everywhere and none at the jump, and the gene–disease pair count ∑**1** *z*_*mkd*_ *>* 0 is piecewise constant for the same reason. The count is therefore replaced by its expectation *ν*_0_ ∑ Pr(*z*_*mkd*_ *>* 0), which is smooth in *l*_*mkd*_ (above), and the draw by the “hard concrete” distribution of Louizos et al. [43], their term for a binary concrete variable stretched to [*γ*_*g*_, *ζ*] and clamped to [0, 1]. This puts atoms at exactly 0 and 1, so gene–disease pairs can still be switched off outright, while *z*_*mkd*_ is a differentiable function of *l*_*mkd*_ for almost every draw. With *u* ∼ *U*(0, 1),

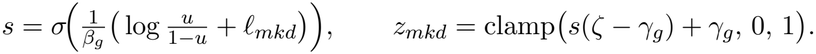

The selection probability is then *ω*_*mkd*_ = Pr(*z*_*mkd*_ *>* 0) = *σ*, so *l*_*mkd*_ is the logit of *ω*_*mkd*_ shifted by a constant fixed by the stretch, here *β*_*g*_ log(−*γ*_*g*_*/ζ*) = −1.60. Selection parameters are initialized at *l*_*mkd*_ = 1.5, giving *ω*_*mkd*_ = 0.96: almost every pair starts selected, and the spike removes those the data do not support.

### Frailty during carrier fits and prediction

The carrier effect *η*_*mkd*_(*t*) (eq. (4)) is estimated against 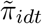, with each individual’s *f*_*i*_ held at its fitted value, so that the carrier effect is not asked to absorb rate that the frailty already carries. For prediction, *f*_*i*_ is refit together with *δ* from the individual’s records up to the prediction age, under the same prior (Methods, *Model fitting* ). Carrier-penetrance curves are reported both at *f*_*i*_ = 0, an individual at the population-average rate, and at the fitted *f*_*i*_.

### Observed ovarian comparison and the breast signature

For the observed ovarian comparison we used women in the model cohort (80 *BRCA1* carriers, 313 *BRCA2* carriers and 217,065 non-carriers) and computed Kaplan–Meier cumulative incidence and hazards in 10-year age bands. Risk-reducing surgery is not recorded, so carriers who had it remain in the risk set.

### Sex-limited diseases

A disease is *sex-limited* when only one sex can develop it. Prevalence 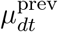 is computed once in preprocessing as the incidence among those still at risk (*E*_*id*_ ≥ *t*), smoothed on the logit scale, with both sexes in the denominator. For a sex-limited disease that denominator therefore includes people who cannot develop the disease, so 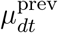 falls below the incidence among the sex at risk: women remain in the prostate cancer denominator. Training compensates through *ψ*_*kd*_ and *ϵ*_*kdt*_, and sex is a component of *G*_*i*_, so it enters *λ* and hence *θ*: women have *θ*_S13_ ≈ 0 on the male signature while men load it at 0.88. The trained base model is therefore calibrated population-wide: predicted prostate cancer cumulative incidence in men is 6.3% against an observed 6.2% at age 70, so neither *µ*_*dt*_ nor *ϕ*_*kdt*_ needs any correction. Carrier penetrance is then reproduced by *η*(*t*) acting on these fixed, calibrated population parameters with *δ* estimated: for prostate cancer the control cumulative incidence at age 70 is 5.6% (6.8% in the cancer registry) with the carrier effect set to zero, and an estimated *η*(*t*) reproduces both the 2.3-fold higher cumulative incidence observed in carriers and its earlier onset. Breast cancer is assigned the same way, to S19, the signature that expresses it, with *µ*_*d*_ computed among women. With the age-varying healthy reference, women’s baseline is not held in a female cancer signature, so carriers’ excess acts through S19 directly and the observed carrier penetrance is within reach of *η* (Fig. 3).

### Per-SD hazard ratios by age

In each five-year window we regressed the window outcome (a first diagnosis during the window) on the standardized score *z* by ordinary least squares, giving slope *β* and mean outcome 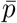, and converted the slope to a per-SD odds ratio, 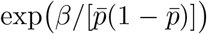, using the logistic derivative at 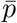; for rare outcomes this approximates the per-SD hazard ratio. For the model curves the outcome is replaced by the participant’s mean predicted yearly probability over the window, with the same windows, at-risk sets and score standardization. Restricting each window to participants still at risk removes most of the selection that comes from following people of different enrollment ages over different lengths of time; the mean score of the at-risk set changes by less than 0.1 standard deviation by age 75, so selection explains little of the decline.

### Quintile event rates

As a check that needs neither the regression nor its conversion, we divided participants into score quintiles and computed, in each window, the fraction of those at risk with a first diagnosis, for the observed data and for the model’s predicted probabilities. The ratio between the top and bottom quintiles falls with age in both (Fig. 1c).

### *LDLR* risk groups

Ranking within five-year groups of enrollment age gives thirds with the same mean age (56.3–56.7 years). For each third we report the mean predicted and observed ten-year risk and a summary of the group (mean age, polygenic score percentile, number of prior diagnoses, and the prior diagnoses that differ most between the highest and lowest thirds). The two example patients in Fig. 5c are these group averages, not individual records.

### *LDLR* reclassification

The ten-year ASCVD risk combines seven diseases as if they were independent, which overstates predicted risk, so we recalibrated it to the observed population rate with one intercept (*α* = −0.69; mapping 3 of Table S11), estimated in non-carriers and applied to carriers. The risk with the carrier term is this recalibrated risk multiplied by 1.23, the age- and sex-matched rate ratio between carriers and non-carriers in UK Biobank. Of the carriers who cross a treatment threshold once carrier status is used, those crossing 7.5% (*n* = 52) had observed ten-year ASCVD risk of 13%, and those crossing 20% (*n* = 28), 11%.

## Supplementary Figures

**Figure S1:**
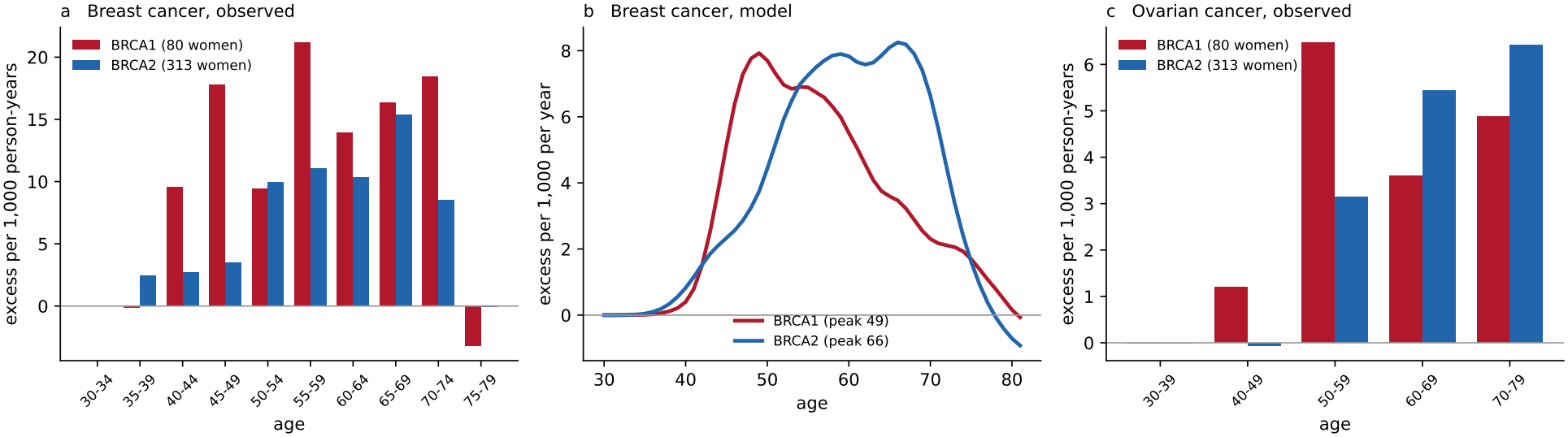
Carrier effects arise earlier for *BRCA1* than for *BRCA2*, in the recorded diagnoses and in the model. UK Biobank women (80 *BRCA1* and 313 *BRCA2* carriers). *(a)* Observed excess breast cancer rate (carriers minus non-carriers, per 1,000 person-years) by 5-year age group, with no model. At ages 40–49 the rate was 3.6 times higher in *BRCA1* than in *BRCA2* carriers (exact binomial *p* = 0.003); median age at diagnosis was 55 and 58 years (Mann–Whitney *p* = 0.04). *(b)* Model excess breast cancer incidence (carriers minus matched controls) from the age-varying carrier effect, which peaks at age 49 for *BRCA1* and 66 for *BRCA2*. (Fig. 3c reports the peak of the carrier effect itself, 48 and 66.) *(c)* Observed excess ovarian cancer rate by 10-year age group. The excess is highest at 50–59 in *BRCA1* carriers and at 60–79 in *BRCA2* carriers; the *BRCA1* estimate rests on nine cases (rate ratio at 40–59, 2.4; *p* = 0.08). Diagnoses before enrollment (ages 40–69) are recorded retrospectively, which affects both genes alike and so does not bias the comparison between them. Risk-reducing surgery is not censored; because it is recommended earlier for *BRCA1* carriers, it would lower their observed rates more, working against the earlier *BRCA1* excess.

**Figure S2:**
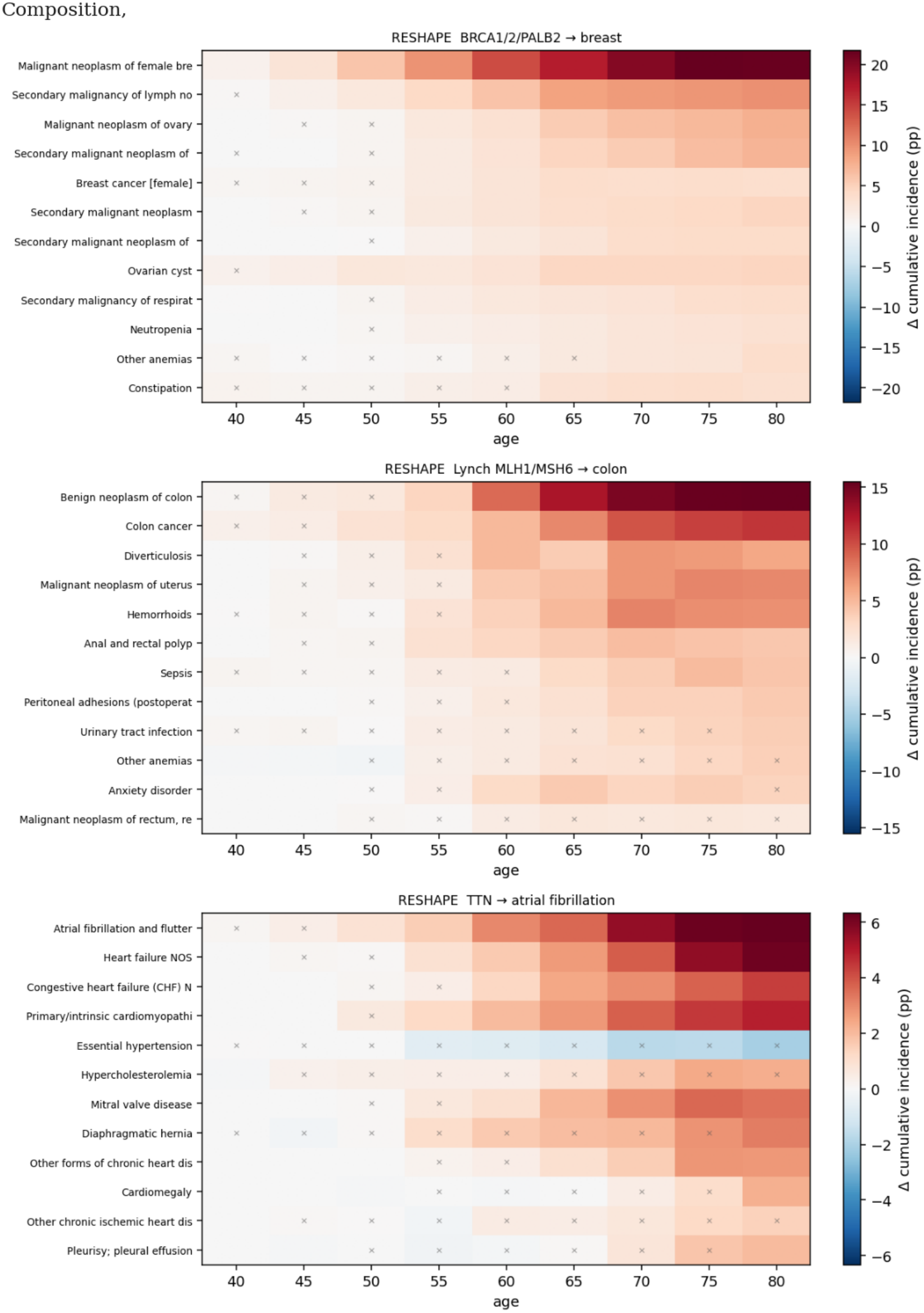
Diseases in excess among carriers, model-free. Difference in cumulative incidence (percentage points) between carriers and all non-carriers, by disease and age (40–80), with no conditioning on the primary diagnosis: *BRCA1* /*BRCA2* /*PALB2* carriers (*top*; primary disease breast cancer), Lynch (*MLH1* /*MSH6* ) carriers (*middle*; colon cancer) and *TTN* carriers (*bottom*; atrial fibrillation). No fitted quantity enters. In each group the excess extends beyond the primary disease to other diseases, for example ovarian cancer and secondary malignancies for *BRCA1* /*BRCA2* /*PALB2*, uterine cancer for Lynch, and heart failure and cardiomyopathy for *TTN*. The comparison with people at matched polygenic risk is in Table 2.

**Figure S3:**
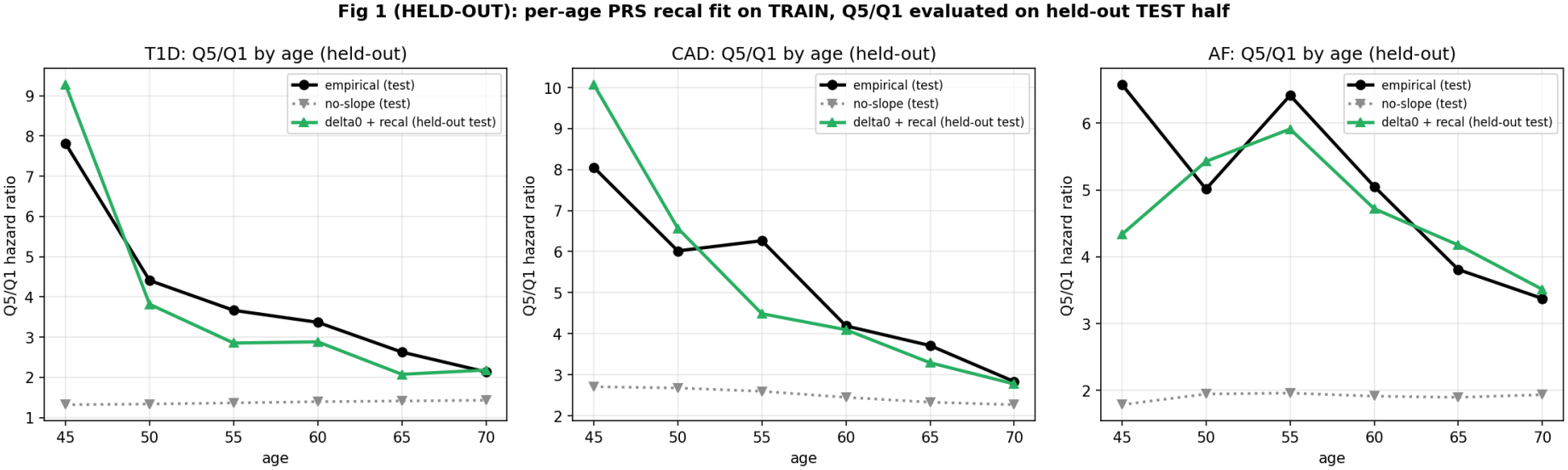
Held-out validation of the per-age PRS recalibration (cf. Fig. 1c). Recalibration coefficients *c*_*j*_(*a*) fit on a random *train* half; *Q*_5_*/Q*_1_ hazard ratios shown on the disjoint *held-out* test half (observed; model without the age interaction; full model with recalibration). Held-out and train curves coincide at every age (e.g. T1D 9.3 to 2.2 test vs. 9.8 to 2.1 train), so the per-window map does not overfit at *N* ≈ 400,000; Fig. 1c (full cohort) is essentially identical.

**Figure S4:**
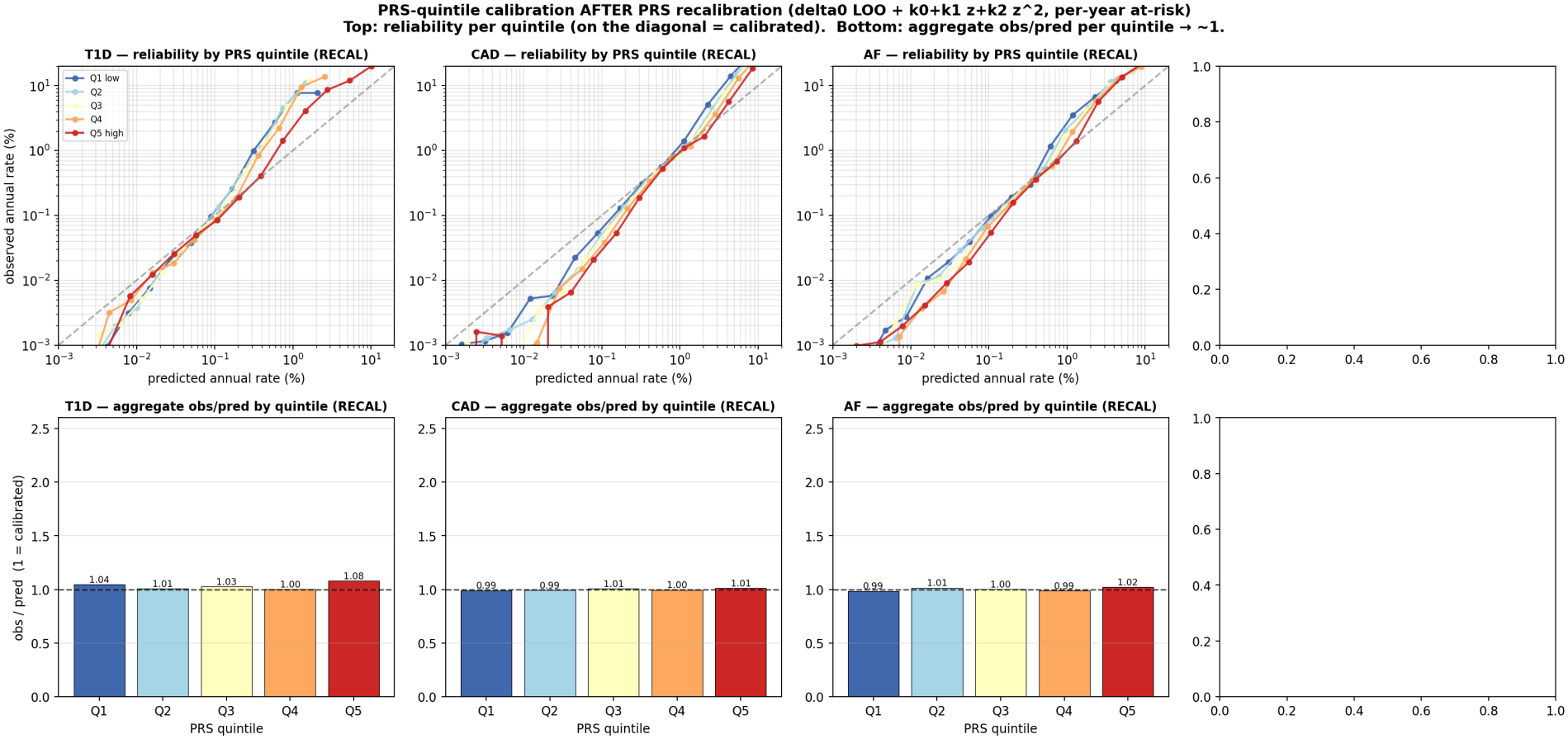
PRS-quintile calibration after the per-age recalibration (leave-one-out predictions with *c*_0_ + *c*_1_*z* + *c*_2_*z*^2^, out-of-sample). Computed on at-risk person-years as in the calibration of Fig. 2d, stratified by PRS quintile, for three exemplar diseases (T1D, CAD, AF). *Top:* reliability per quintile: observed vs. predicted annual rate across bins. *Bottom:* aggregate observed/predicted ratio per quintile (1 = calibrated). After recalibration the ratio is ≈ 1 in all quintiles (T1D 1.04*/*1.01*/*1.03*/*1.00*/*1.08; CAD and AF within 0.99–1.02). The per-age recalibration removes the under-prediction in the high-PRS tail, so the model is calibrated across the full PRS distribution as well as on average.

**Figure S5:**
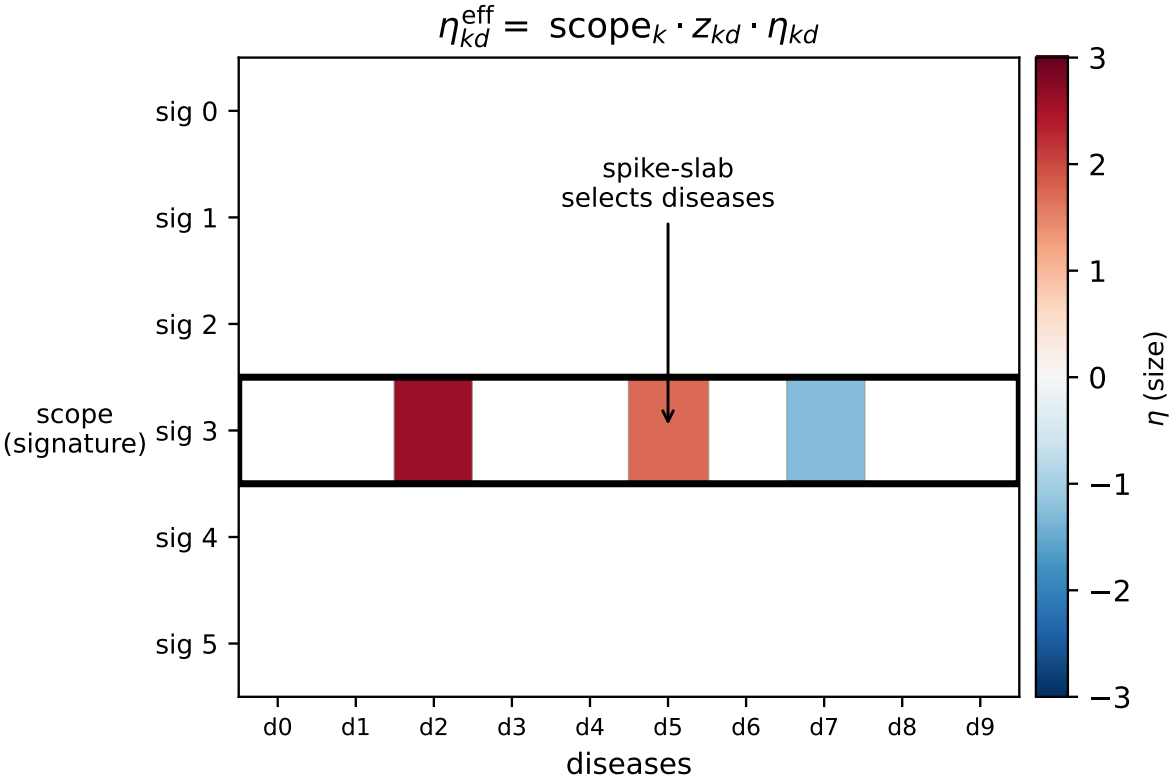
The disease-specific carrier effect: signature assignment and spike-and-slab prior. For gene *m*: the assignment scope_*mk*_ selects one signature row *k*^*^; the *spike* (the selection indicator *z*) selects a few gene–disease pairs (which diseases); the *slab* (*v*) sets how large the effect is. *η*_*mkd*_ = scope_*mk*_ *z*_*mkd*_ *v*_*mkd*_.

**Figure S6:**
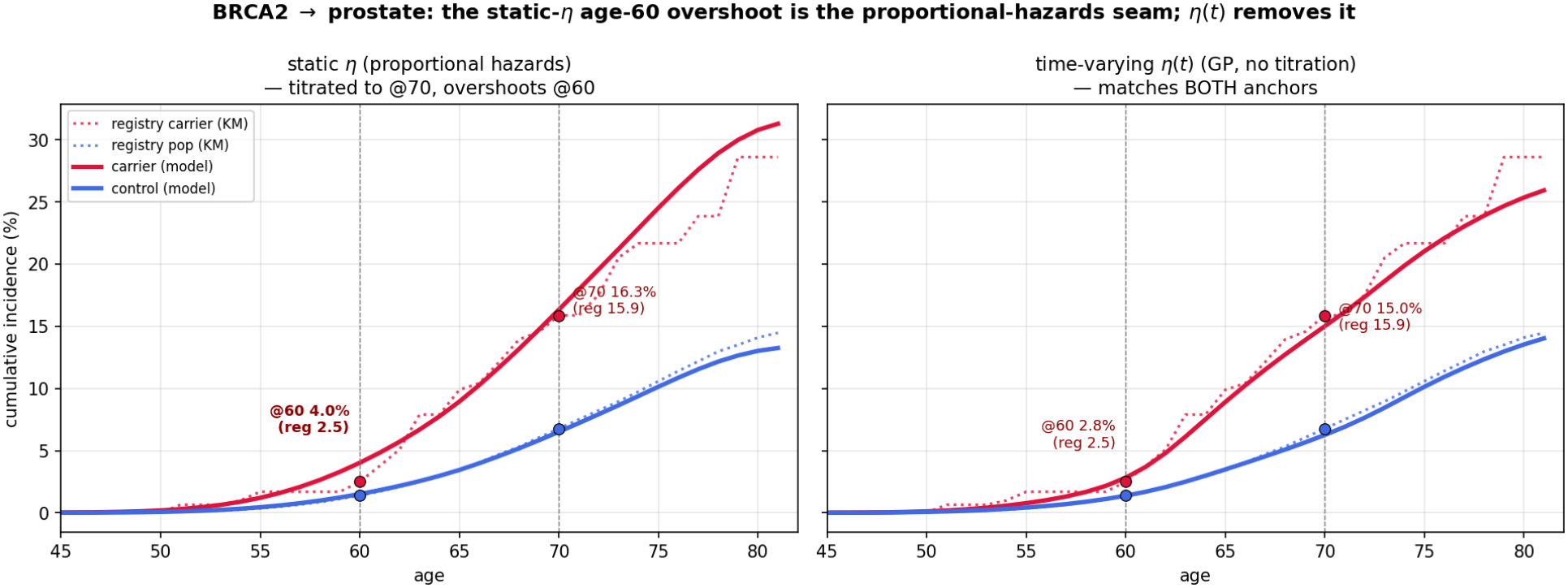
Constant and age-varying carrier effects for *BRCA2* and prostate cancer, against the cancer registry. *Left:* a carrier effect held constant in age, chosen to match the registry carrier rate at age 70, over-predicts at age 60 (model 4.0%, registry 2.5%). *Right:* an age-varying carrier effect, fitted to the registry diagnoses, reproduces both the age-60 and age-70 values. Dotted, registry Kaplan–Meier (carriers and population); solid, model; points, registry at ages 60 and 70. This illustration uses a single-gene fit; the main registry-calibrated fit gives 13.4% at age 70 (Supp. Table S6).

**Figure S7:**
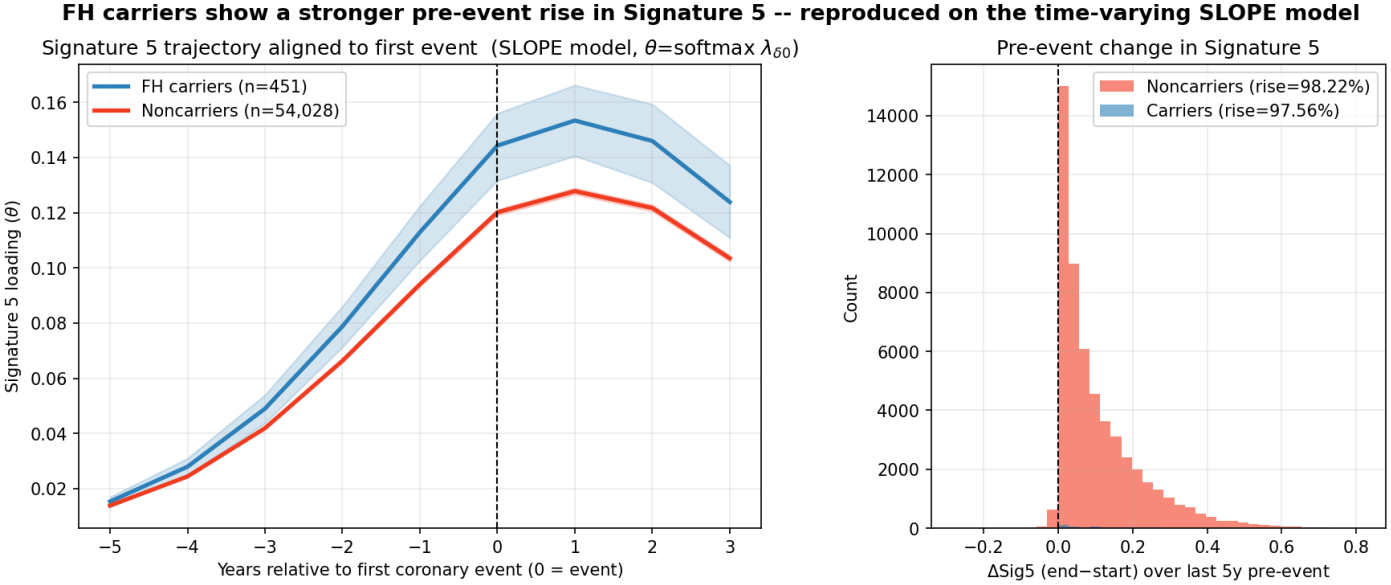
Familial-hypercholesterolemia carriers load higher on Signature 5 before their first coronary event. Signature loading *θ* from the full model with the age interaction. Left: S5 loading aligned to the first coronary event; FH carriers (*n*=451) sit above non-carriers (*n*=54,028) at every pre-event year, peaking ∼ 0.15 vs. ∼ 0.13 one year after the event. Right: change in S5 loading over the five years before the event; nearly everyone’s loading rises (carriers 97.6%, non-carriers 98.2%), so the carrier signal is the loading *magnitude*, not its direction. A clinical example of carrier status reflected in the signature loading.

**Figure S8:**
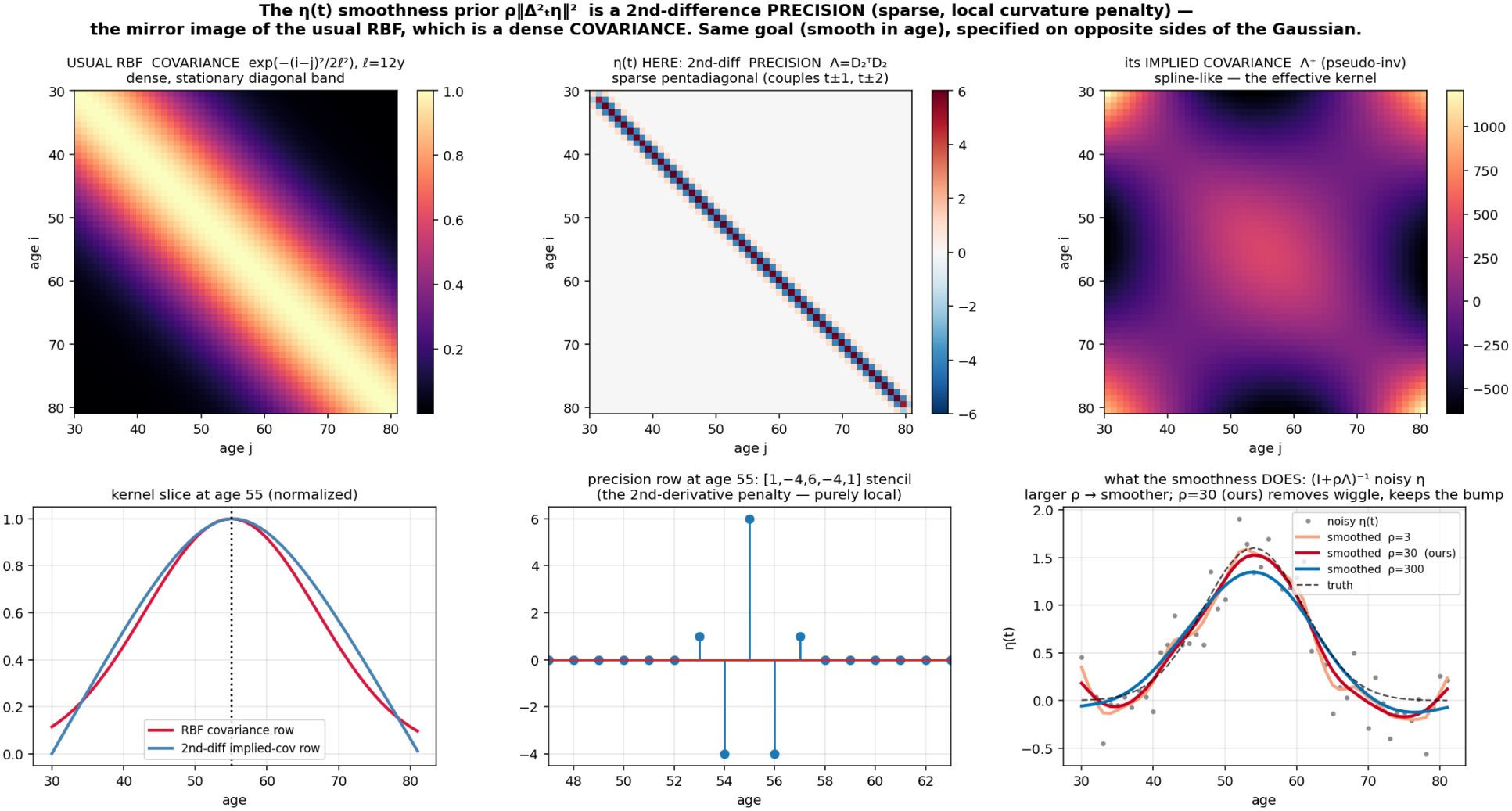
The *η*(*t*) smoothness prior is a second-difference *precision* (a local curvature penalty), the mirror image of an RBF covariance. The age-varying carrier effect is smoothed by 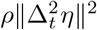with precision 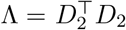. *Top:* the usual RBF is a dense, stationary *covariance* (left); our prior is instead a sparse pentadiagonal *precision* coupling only *t ±* 1, *t ±* 2 (middle), whose implied covariance is spline-like (right). *Bottom:* the precision row is the second-derivative stencil [1, − 4, 6, − 4, 1], purely local (middle); applying (*I* +*ρ*Λ)^−1^ to a noisy *η* removes high-frequency variation while preserving the age-of-onset bump (*ρ* = 30 used here, left/right). Penalizing *curvature* (not level or slope) lets *η*(*t*) bend smoothly without shrinking its magnitude or manufacturing age-dependence; this is the second-order random-walk (cubic smoothing spline) prior of the Methods.

**Figure S9:**
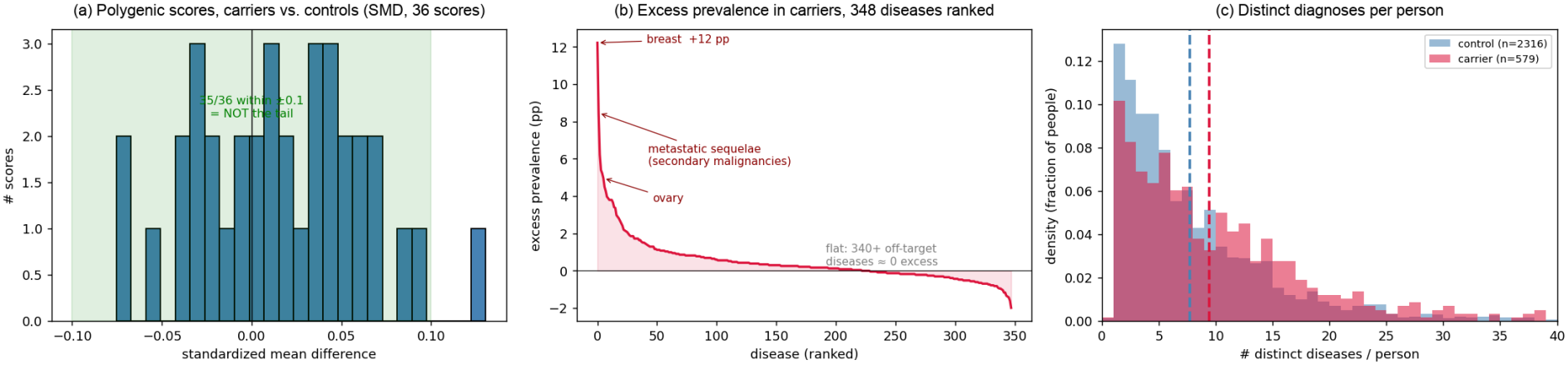
*BRCA2* carriers are not concentrated in the polygenic tail, and their excess is confined to a few diseases. *(a)* Standardized mean difference of the 36 polygenic scores between carriers and matched controls; 35 of 36 lie within *±* 0.1. *(b)* Excess lifetime prevalence in carriers (carriers minus controls, percentage points) for each of the 348 diseases, ranked: breast cancer (+12), secondary malignancies and ovarian cancer lead, and most other diseases show no excess. *(c)* Number of distinct diagnoses per person: 9.4 in carriers against 7.7 in controls, a difference made up mostly of cancers.

**Figure S10:**
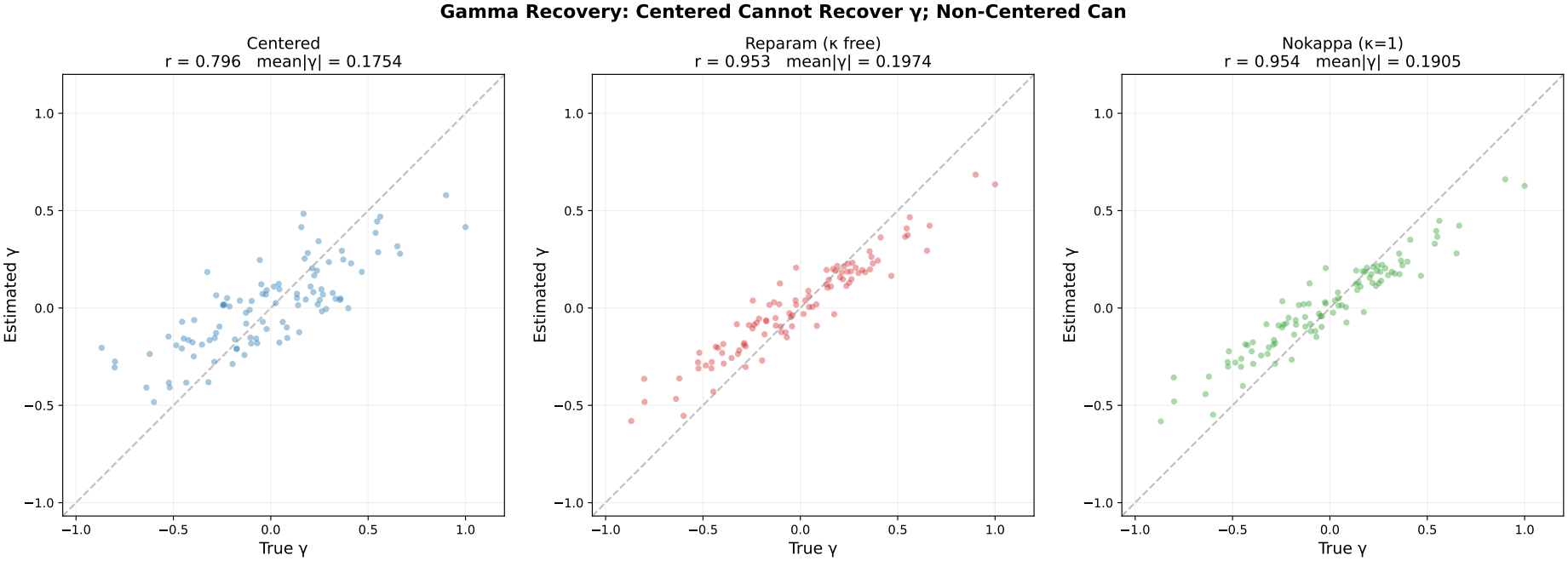
The age-varying genetic effect is identifiable (simulation). Estimated vs. true *γ* across parameterizations: the centered parameterization cannot recover it (*r* = 0.80); the non-centered / *κ*=1 parameterization used here does (*r* = 0.95).

**Figure S11:**
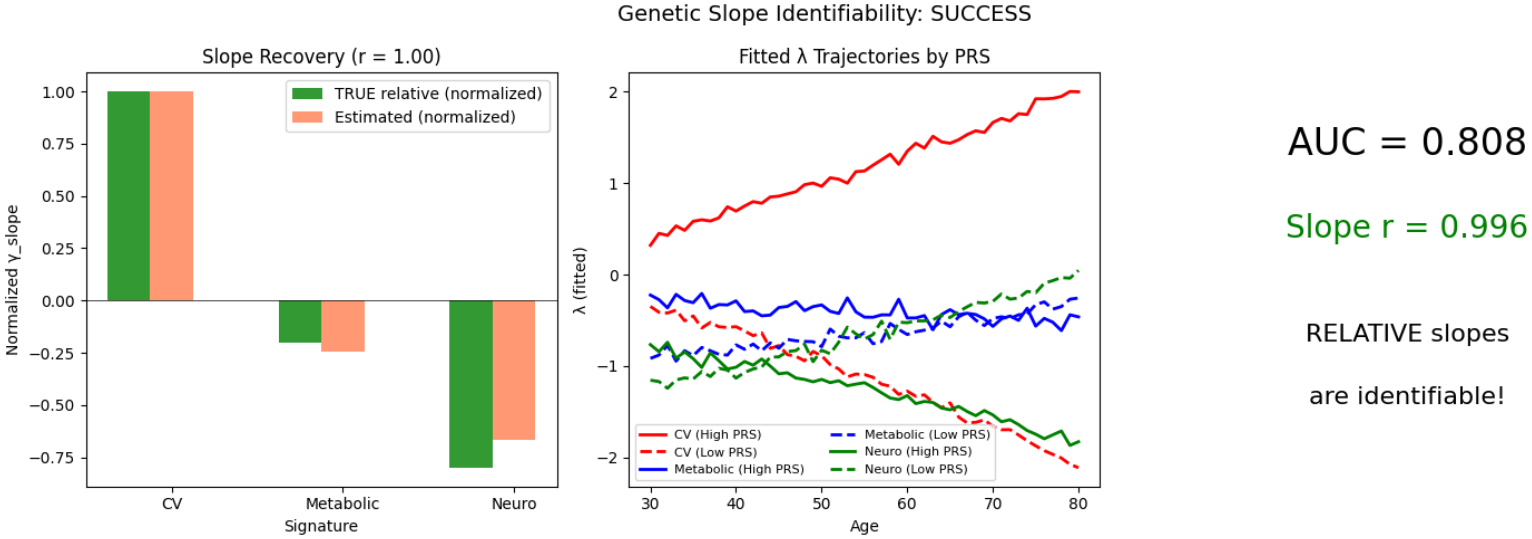
The health anchor *α*_*i*_ makes *absolute* age interactions identifiable (simulation). Without an anchor only relative age interactions are recovered (left, *r* = 0.996, relative scale); with the fixed health baseline *α*_*i*_ = *G*_*i*_*γ*^health^ the *absolute* age interactions are recovered (right, *r* = 0.974).

**Figure S12:**
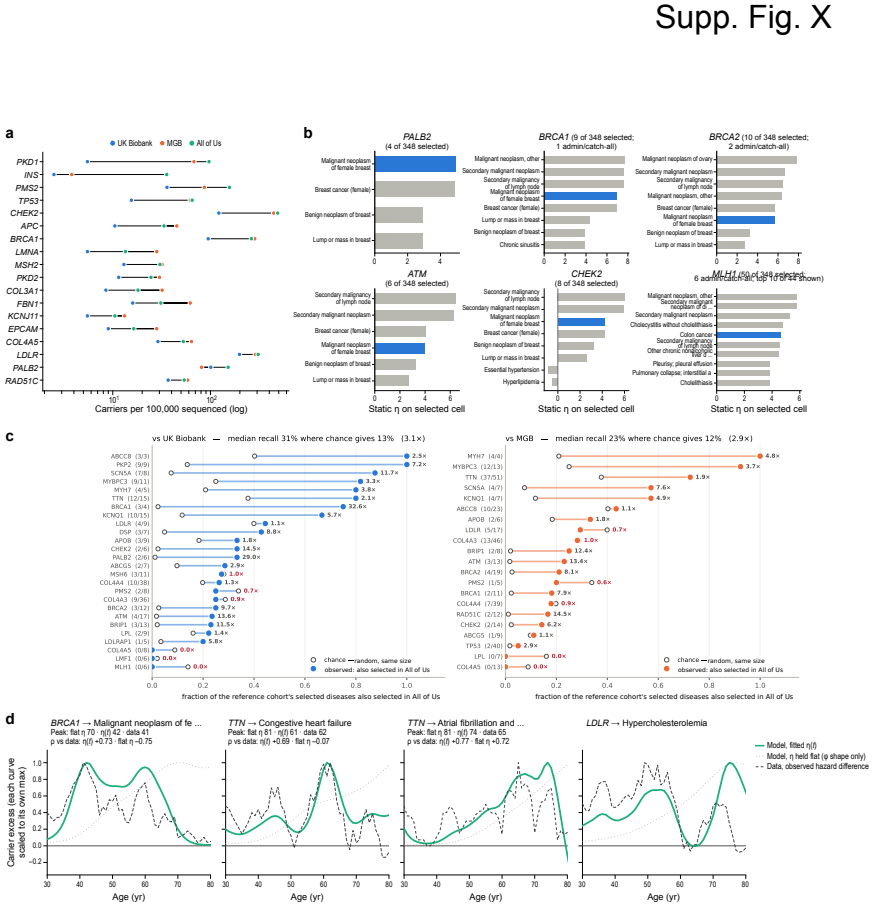
The All of Us replication. *(a) Ascertainment*. Carriers per 10^5^ sequenced per gene, on each cohort’s own sequenced denominator (UK Biobank 200,343, MGB 53,111, All of Us 170,825; Table 1, Supp. Tables S3, S4). *PKD1*, with six pseudogenes, has the largest All of Us to UK Biobank rate ratio, 17.6*×*, followed by *INS* 14.3 *×, PMS2* 4.4 *×, CHEK2* 4.0 *×, BRCA1* 2.8 *×*, with *STK11, APOC3* and *PROS1* at a UK Biobank rate of zero. *F2* is excluded because rs1799963 lies in the 3’UTR, outside exome capture (it is not removed by filtering). *(b) Which diseases were selected*. Blue, the gene’s known (home) disease; gray, the other diseases the model selected. Bars are ordered by |*η*|, so a gray bar longer than the blue one is another selected disease with a larger carrier effect than the home disease. Selection is sparse: *PALB2* 4 diseases (all breast), *BRCA1* 9, *BRCA2* 10. Administrative and catch-all codes are excluded from the bars and counted in each title; over all 34 genes they are 7% of selected gene–disease pairs and do not differ between genes that select more and fewer diseases. *(c) Selection consistency*. Per gene, the fraction of UK Biobank (or MGB) selected diseases that All of Us also selects, against what a random selection of its size would recover (recall, not Jaccard; Methods). Against UK Biobank, median recall 31% where chance gives 13%, a median 3.1*×* enrichment, with 20*/*26 genes above chance (*BRCA2* 25% vs. 3%); against MGB, 23% vs. 12%, 2.9*×* (15*/*21). Restricted to each gene’s three strongest UK Biobank selected diseases (largest |Δpen|), median recall is 67% (20*/*26 above chance), and 60% for the five strongest (19*/*24). *(d) What η*(*t*) *adds, against observed hazard*. Three curves per gene–disease pair, each scaled to its own maximum so only shape is compared: *η* held at its time-average (dotted: the age-shape a *time-invariant* effect gives, non-flat only because *σ* is nonlinear), fitted *η*(*t*) (solid), and the observed first-event hazard difference against the same matched non-carriers (dashed, no fitted quantity). The solid–dotted separation is large for *BRCA1* (*r*_*s*_ = +0.73 fitted vs. − 0.75 flat), *TTN* → CHF (+0.69 vs. − 0.07) and *LDLR* → hypercholesterolemia (+0.67 vs. − 0.55), negligible for *TTN* → AF (+0.77 vs. +0.72). Against a 90% bootstrap interval on the observed peak: *BRCA1* 42 vs. 41 [39, 60], *TTN* CHF 61 vs. 62 [58, 63], *TTN* → AF 74 vs. 65 [61, 75], all inside; *LDLR* → hypercholesterolemia gives 75 vs. 49 [47, 56] because model and data differ in which mode is tallest, not in where the modes lie (Supplementary Note 1). Panels (b)–(d) are in-sample: *η* was fitted on these carriers against these controls. Read-out conventions in Methods.

**Figure S13:**
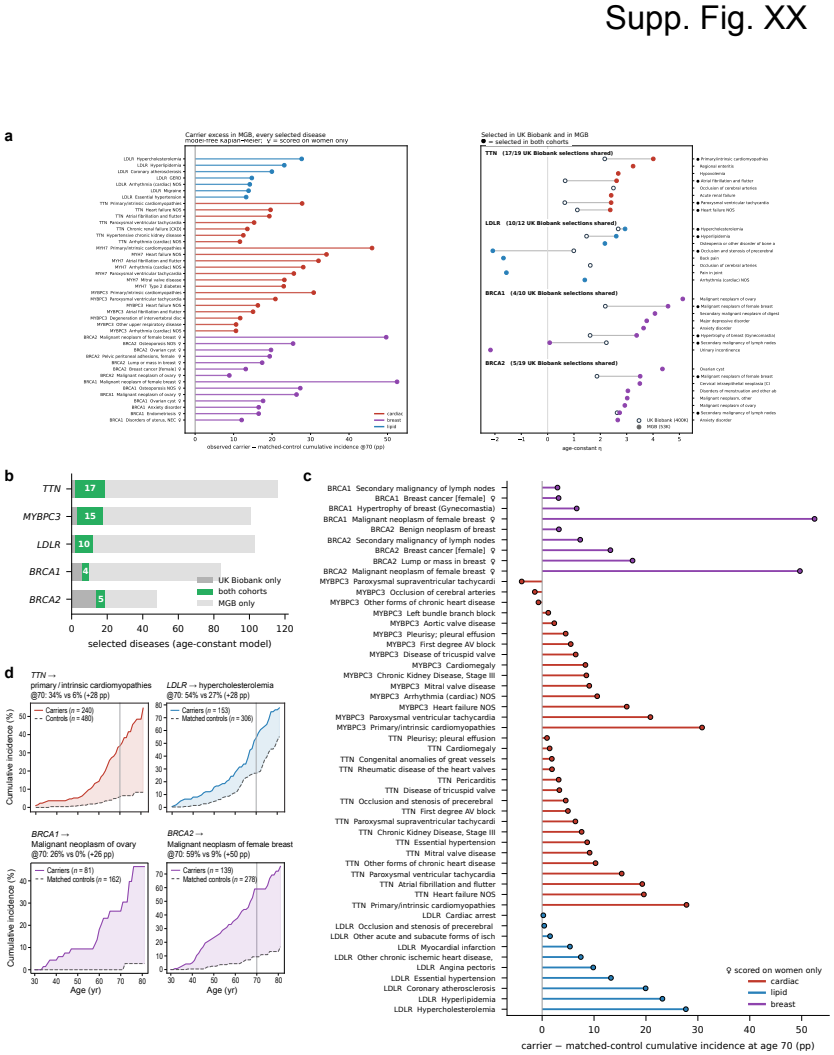
The MGB replication: the age-constant model. Throughout, carrier–control separation is read from the observed data without the model; the fitted carrier effect enters only in Supp. Fig. S14. *(a) The other selected diseases show excess in MGB’s observed data, and UK Biobank selected the same diseases*. For every selected gene–disease pair, the observed Kaplan–Meier separation between carriers and matched controls at age 70 (*left* ); and, for the genes with other selected diseases in UK Biobank, the two cohorts’ selected-disease lists side by side against the age-constant *η* for that pair (*right* ), annotated with the share of UK Biobank’s selected pairs MGB also selects: *TTN* 17*/*19, *LDLR* 10*/*12, *BRCA1* 4*/*10, *BRCA2* 5*/*19. Women-only pairs are marked. *(b) Selection overlap*. Gene–disease pairs selected by the age-constant model in each gene, split into UK Biobank only, both cohorts, and MGB only; the shared count is labeled (*TTN* 17, *MYBPC3* 15, *LDLR* 10, *BRCA1* 4, *BRCA2* 5; 51 shared pairs in all). *(c) Kaplan– Meier for shared selections*. Pairs selected in *both* UK Biobank and MGB, with the observed difference in cumulative incidence at age 70 between carriers and matched controls for each of the 51 pairs. *(d) Four pairs in full*. Observed cumulative incidence in MGB carriers and matched controls, one pair per gene family: *TTN* → primary/intrinsic cardiomyopathies (34% vs. 6% at age 70, +28 pp), *LDLR* → hypercholesterolemia (54% vs. 27%, +28 pp), *BRCA1* → ovarian cancer (26% vs. 0%, +26 pp) and *BRCA2* → female breast cancer (59% vs. 9%, +50 pp). *Conventions*. All four panels use the *age-constant* model, under which the other selected diseases appear in both cohorts; UK Biobank’s age-varying model selects four diseases for *BRCA1* against 10 for the age-constant model on the same assigned signature. Carriers and matched controls are followed under identical entry and censoring, so these panels are within-cohort contrasts, insensitive to the cohorts’ differing follow-up. Incidence is read on the Kaplan–Meier scale because MGB’s censoring time is the end of a fragmentary EHR record rather than a uniform administrative date. Absolute penetrance is estimated separately, against registry curves (Fig. 3d). Entry and censoring conventions in Methods.

**Figure S14:**
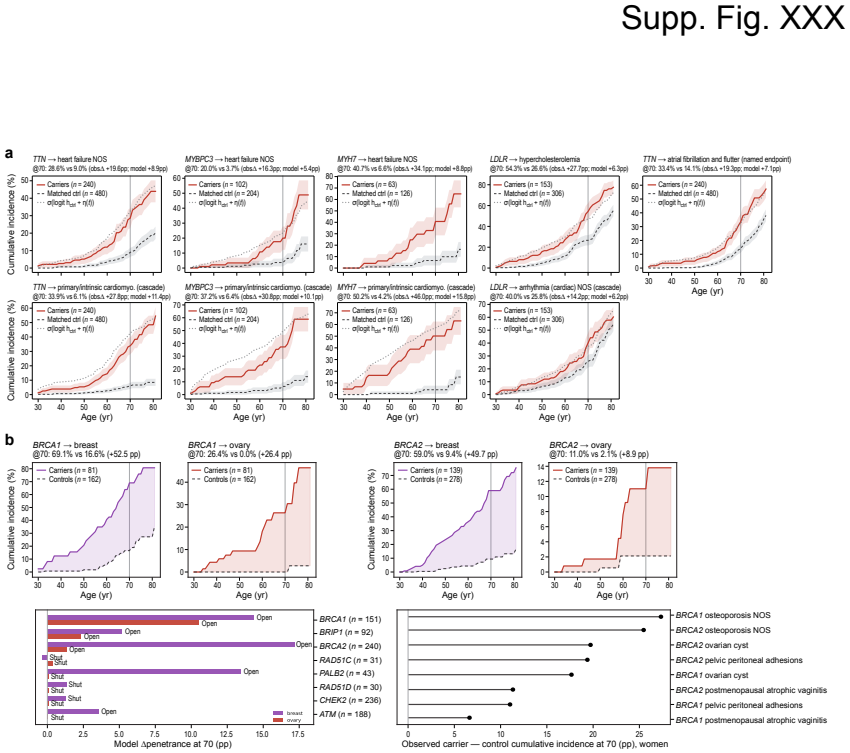
The MGB replication: what the fitted *η*(*t*) implies for observed incidence. Companion to Supp. Fig. S13, which shows the model-free age-constant results. *(a) Observed Kaplan–Meier against the penetrance difference η*(*t*) *implies*. Observed cumulative incidence in the *same* carriers and the *same* matched controls the atlas was fitted on, for each gene’s named target endpoint (*top row*, five) plus the top other selected disease (*bottom row*, four). The dotted curve takes the observed *control* hazard as the empirical estimate of *σ*(*ϕ*), adds the fitted *η*(*t*) in log-odds, and returns it through the same product-limit form, *σ*(logit *h*_ctrl_(*t*) + *η*(*t*)). *(b) Gene-specific organ involvement under a signature assignment fixed in advance*. The eight breast-panel genes under a single pre-specified assignment: the *union* of the expression home (S16) and the largest female signature (S10), applied identically to all eight with no per-gene tuning. The spike-and-slab prior selects ovarian cancer in *BRCA1, BRCA2* and *BRIP1* and does not select it in *ATM, CHEK2* and *PALB2*, separating genes with established ovarian risk from breast-predominant ones, and the observed data agree (female *BRCA1* carriers reach 26.4% ovarian cumulative incidence by age 70 against 0% in matched controls). *RAD51C* and *RAD51D* carry known ovarian risk but were not selected with only 31 and 30 carriers. Per-gene carrier effects in Supp. Table S5, from the same fit. *Conventions*. Carriers and matched controls are followed under identical entry and censoring, so these panels are within-cohort contrasts, insensitive to the cohorts’ differing follow-up. Incidence is read on the Kaplan–Meier scale because MGB’s censoring time is the end of a fragmentary EHR record rather than a uniform administrative date. Absolute penetrance is estimated separately, against registry curves (Fig. 3d). Entry and censoring conventions in Methods.

**Table S1:**
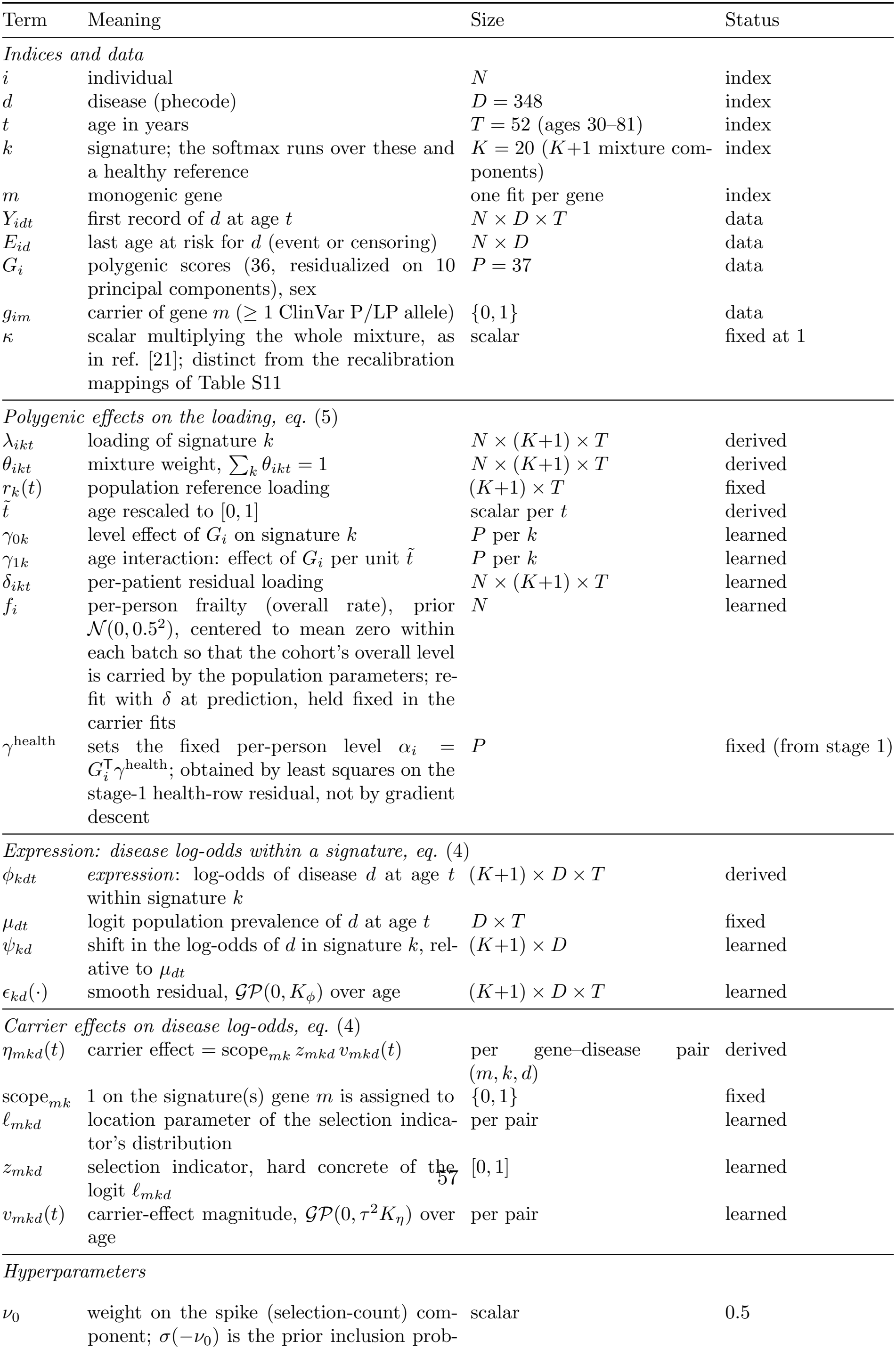
**Terms of equations** (4) **and** (5). “Fixed” means set before the fit and not updated; “learned” means updated by gradient descent. *σ* is the logistic function and logit = *σ*^−1^. Status refers to the population fit (stages 1–2 of Table S2); every learned term except *δ* is held fixed in stage 3. The healthy reference contributes a row to every *k*-indexed object. A *gene–disease pair* is a triple (*m, k, d*) with scope_*mk*_ = 1. The *spike* and the *slab* are the two components of the prior on the carrier effect, defined below: a point mass at zero, and a Gaussian on the magnitude.

| Term | Meaning | Size | Status |
|---|---|---|---|
| *Indices and data* |  |  |  |
| *i* | individual | *N* | index |
| *d* | disease (phecode) | *D* = 348 | index |
| *t* | age in years | *T* = 52 (ages 30–81) | index |
| *k* | signature; the softmax runs over these and a healthy reference | *K* = 20 (*K*+1 mixture components) | index |
| *m* | monogenic gene | one fit per gene | index |
| *Y*<sub>*idt*</sub> | first record of *d* at age *t* | *N* × *D* × *T* | data |
| *E*<sub>*id*</sub> | last age at risk for *d* (event or censoring) | *N* × *D* | data |
| *G*<sub>*i*</sub> | polygenic scores (36, residualized on 10 principal components), sex | *P* = 37 | data |
| *g*<sub>*im*</sub> | carrier of gene *m* (≥ 1 ClinVar P/LP allele) | {0, 1} | data |
| *κ* | scalar multiplying the whole mixture, as in ref. [21]; distinct from the recalibration mappings of Table S11 | scalar | fixed at 1 |
| *Polygenic effects on the loading, eq.* (5) |  |  |  |
| *λ*<sub>*ikt*</sub> | loading of signature *k* | *N* × (*K*+1) × *T* | derived |
| *θ*<sub>*ikt*</sub> | mixture weight, ∑<sub>*k*</sub> *θ*<sub>*ikt*</sub> = 1 | *N* × (*K*+1) × *T* | derived |
| *r*<sub>*k*</sub>(*t*) | population reference loading | (*K*+1) × *T* | fixed |
| *t̃* | age rescaled to [0, 1] | scalar per *t* | derived |
| *γ*<sub>0*k*</sub> | level effect of *G*<sub>*i*</sub> on signature *k* | *P* per *k* | learned |
| *γ*<sub>1*k*</sub> | age interaction: effect of *G*<sub>*i*</sub> per unit *t̃* | *P* per *k* | learned |
| *δ*<sub>*ikt*</sub> | per-patient residual loading | *N* × (*K*+1) × *T* | learned |
| *f*<sub>*i*</sub> | per-person frailty (overall rate), prior 𝒩(0, 0.5<sup>2</sup>), centered to mean zero within each batch so that the cohort's overall level is carried by the population parameters; re-fit with *δ* at prediction, held fixed in the carrier fits | *N* | learned |
| *γ*<sup>health</sup> | sets the fixed per-person level *α*<sub>*i*</sub> = *G*<sub>*i*</sub><sup>T</sup>*γ*<sup>health</sup>; obtained by least squares on the stage-1 health-row residual, not by gradient descent | *P* | fixed (from stage 1) |
| *Expression: disease log-odds within a signature, eq.* (4) |  |  |  |
| *ϕ*<sub>*kdt*</sub> | *expression*: log-odds of disease *d* at age *t* within signature *k* | (*K*+1) × *D* × *T* | derived |
| *μ*<sub>*dt*</sub> | logit population prevalence of *d* at age *t* | *D* × *T* | fixed |
| *ψ*<sub>*kd*</sub> | shift in the log-odds of *d* in signature *k*, rel-ative to *μ*<sub>*dt*</sub> | (*K*+1) × *D* | learned |
| *ϵ*<sub>*kd*</sub>(·) | smooth residual, 𝒢𝒫(0, *K*<sub>*ϕ*</sub>) over age | (*K*+1) × *D* × *T* | learned |
| *Carrier effects on disease log-odds, eq.* (4) |  |  |  |
| *η*<sub>*mkd*</sub>(*t*) | carrier effect = scope<sub>*mk*</sub> *z*<sub>*mkd*</sub> *v*<sub>*mkd*</sub>(*t*) | per gene–disease pair (*m*, *k*, *d*) | derived |
| scope<sub>*mk*</sub> | 1 on the signature(s) gene *m* is assigned to | {0, 1} | fixed |
| *ℓ*<sub>*mkd*</sub> | location parameter of the selection indica-tor's distribution | per pair | learned |
| *z*<sub>*mkd*</sub> | selection indicator, hard concrete of the logit *ℓ*<sub>*mkd*</sub> | [0, 1] | learned |
| *v*<sub>*mkd*</sub>(*t*) | carrier-effect magnitude, 𝒢𝒫(0, *τ*<sup>2</sup>*K*<sub>*η*</sub>) over age | per pair | learned |
| *Hyperparameters* |  |  |  |
| *ν*<sub>0</sub> | weight on the spike (selection-count) com-ponent; *σ*(−*ν*<sub>0</sub>) is the prior inclusion prob-[illegible] | scalar | 0.5 |

**Table S2:** The three estimation stages. Stage 1 is run once per batch of 10,000. Its *γ*_0_ and *ψ* are averaged across all 40 batches before stage 2 begins; *ϵ* is carried over from each batch’s own stage-1 fit, and *δ* is discarded and re-initialized at zero.

| Stage | Data | Estimated | Held |
|---|---|---|---|
| 1 | full cohort, 40 leave-one-batch-out folds | ψ, ϵ, γ<sub>0</sub>, γ<sup>health</sup>, δ, *f* | γ<sub>1</sub> ≡ 0, κ = 1 |
| 2 | all 40 batches of 10,000, fit separately | γ<sub>1</sub>, δ, *f*; γ<sub>0</sub> carried over and up-dated at the same rate as γ<sub>1</sub>, ψ and ϵ at one tenth of it | κ = 1 |
| 3 | carriers and matched non-carriers (sex, 5-year age bin) | ℓ<sub>*mkd*</sub>, *v*<sub>*mkd*</sub>(·), δ | all population parameters, at their stage-2 values |

**Table S3:** Variant curation and ascertainment by cohort. Panel positions queried (identical list in all three cohorts), carriers observed, and rate per 10^5^ sequenced. Denominators are each cohort’s sequenced set: UK Biobank 200,343 (200K exome release, not the 407,878 model cohort), MGB 53,111, All of Us 170,825 (analysis cohort). Assays differ: UK Biobank exome **rLCR SEGDUP-filtered**, MGB exome unfiltered, All of Us WGS. MGB being exome *and* unfiltered is what identifies the filter rather than the assay as the cause of the UK Biobank deficit. *F2* is excluded from that interpretation: rs1799963 lies in the 3’ UTR, outside exome capture entirely. ClinVar review status was not retained when the carrier lists were built and is not reported here.

| Gene | Panel | UKB/10<sup>5</sup> | MGB/10<sup>5</sup> | AoU/10<sup>5</sup> | AoU:UKB | Variants 0 in UKB |
|---|---|---|---|---|---|---|
| STK11 | 400 | 0.0 | 0.0 | nan | ∞ | – |
| APOC3 | 2 | 0.0 | 7.5 | nan | ∞ | – |
| PROS1 | 142 | 0.0 | 11.3 | 31.6 | ∞ | – |
| PKD1 | 2245 | 5.5 | 67.8 | 96.6 | 17.59 | – |
| INS | 40 | 2.5 | 3.8 | 35.7 | 14.31 | – |
| PMS2 | 934 | 35.9 | 86.6 | 156.3 | 4.35 | 12 |
| TP53 | 1142 | 15.5 | 62.1 | 65.0 | 4.20 | – |
| CHEK2 | 1046 | 121.8 | 444.4 | 487.6 | 4.00 | 17 |
| APC | 2590 | 10.5 | 45.2 | 33.4 | 3.18 | – |
| BRCA1 | 4106 | 94.8 | 284.3 | 262.8 | 2.77 | 46 |
| LMNA | 543 | 5.5 | 28.2 | 13.5 | 2.45 | – |
| MSH2 | 2088 | 13.0 | 32.0 | 30.4 | 2.35 | – |
| PKD2 | 439 | 11.5 | 30.1 | 24.6 | 2.14 | – |
| COL3A1 | 945 | 8.5 | 32.0 | 18.1 | 2.14 | – |

**Table S4:**
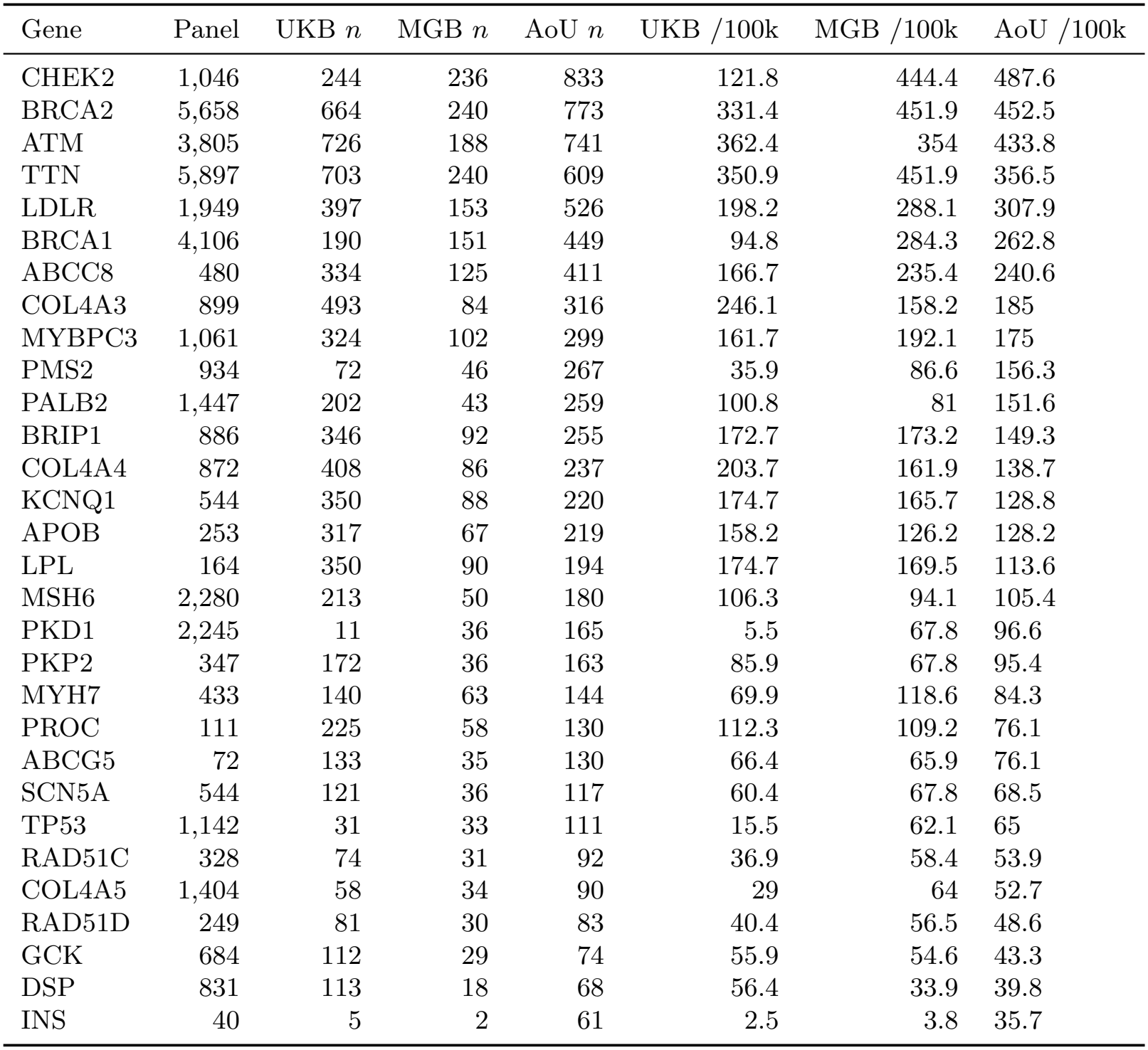
Carrier ascertainment across the three cohorts. Per-gene ClinVar P/LP carrier counts and rates per 100,000 sequenced participants. Denominators differ and are each cohort’s *sequenced* set: UK Biobank 200,343 (the 200K exome release), MGB 53,111, All of Us 170,825 (the analysis set: participants in the model data with whole-genome sequence, EHR and recorded sex). The ascertainment contrast is model-free. UK Biobank’s exome callset is filtered for low-complexity and segmental-duplication regions, and that filter removes recurrent pathogenic alleles: *PMS2* is unestimable there (72 carriers), while All of Us (whole-genome, so unaffected) finds 267, a 4.35*×* rate. The same holds for *CHEK2* (4.00*×* ) and *BRCA1* (2.77*×* ). MGB, also exome but without that filter, sits between the two, which points to the filter, not the assay, as the cause. *F2* is the extreme case (234 *×*): its causal allele *rs1799963* is in the 3’ UTR and is therefore outside the exome capture entirely, not merely filtered. Genes are labelled by stratum: *commonallele* genes are driven by one high-frequency variant and are not interpretable as monogenic carriers.

| Gene | Panel | UKB *n* | MGB *n* | AoU *n* | UKB /100k | MGB /100k | AoU /100k |
|---|---|---|---|---|---|---|---|
| CHEK2 | 1,046 | 244 | 236 | 833 | 121.8 | 444.4 | 487.6 |
| BRCA2 | 5,658 | 664 | 240 | 773 | 331.4 | 451.9 | 452.5 |
| ATM | 3,805 | 726 | 188 | 741 | 362.4 | 354 | 433.8 |
| TTN | 5,897 | 703 | 240 | 609 | 350.9 | 451.9 | 356.5 |
| LDLR | 1,949 | 397 | 153 | 526 | 198.2 | 288.1 | 307.9 |
| BRCA1 | 4,106 | 190 | 151 | 449 | 94.8 | 284.3 | 262.8 |
| ABCC8 | 480 | 334 | 125 | 411 | 166.7 | 235.4 | 240.6 |
| COL4A3 | 899 | 493 | 84 | 316 | 246.1 | 158.2 | 185 |
| MYBPC3 | 1,061 | 324 | 102 | 299 | 161.7 | 192.1 | 175 |
| PMS2 | 934 | 72 | 46 | 267 | 35.9 | 86.6 | 156.3 |
| PALB2 | 1,447 | 202 | 43 | 259 | 100.8 | 81 | 151.6 |
| BRIP1 | 886 | 346 | 92 | 255 | 172.7 | 173.2 | 149.3 |
| COL4A4 | 872 | 408 | 86 | 237 | 203.7 | 161.9 | 138.7 |
| KCNQ1 | 544 | 350 | 88 | 220 | 174.7 | 165.7 | 128.8 |
| APOB | 253 | 317 | 67 | 219 | 158.2 | 126.2 | 128.2 |
| LPL | 164 | 350 | 90 | 194 | 174.7 | 169.5 | 113.6 |
| MSH6 | 2,280 | 213 | 50 | 180 | 106.3 | 94.1 | 105.4 |
| PKD1 | 2,245 | 11 | 36 | 165 | 5.5 | 67.8 | 96.6 |
| PKP2 | 347 | 172 | 36 | 163 | 85.9 | 67.8 | 95.4 |
| MYH7 | 433 | 140 | 63 | 144 | 69.9 | 118.6 | 84.3 |
| PROC | 111 | 225 | 58 | 130 | 112.3 | 109.2 | 76.1 |
| ABCG5 | 72 | 133 | 35 | 130 | 66.4 | 65.9 | 76.1 |
| SCN5A | 544 | 121 | 36 | 117 | 60.4 | 67.8 | 68.5 |
| TP53 | 1,142 | 31 | 33 | 111 | 15.5 | 62.1 | 65 |
| RAD51C | 328 | 74 | 31 | 92 | 36.9 | 58.4 | 53.9 |
| COL4A5 | 1,404 | 58 | 34 | 90 | 29 | 64 | 52.7 |
| RAD51D | 249 | 81 | 30 | 83 | 40.4 | 56.5 | 48.6 |
| GCK | 684 | 112 | 29 | 74 | 55.9 | 54.6 | 43.3 |
| DSP | 831 | 113 | 18 | 68 | 56.4 | 33.9 | 39.8 |
| INS | 40 | 5 | 2 | 61 | 2.5 | 3.8 | 35.7 |

**Table S5:** A pre-specified scope recovers gene-specific cancer spectra in MGB. The scope — expression home union largest female signature — was fixed in advance and applied identically to all eight breast-panel genes, with no per-gene tuning. The spike–slab gate opens ovarian cancer in *BRCA1, BRCA2* and *BRIP1* and leaves it shut in *ATM, CHEK2* and *PALB2*, separating genes with established ovarian risk from breast-predominant ones. The offsets carry the same information: *BRCA1* ‘s ovarian *η exceeds* its breast *η* while *BRCA2* ‘s does not, matching the larger ovarian relative risk of *BRCA1* ; and *BRIP1*, a moderate-penetrance ovarian gene with weak breast evidence, is ovarian-predominant. Nothing in the fit encodes gene-specific cancer spectra. *RAD51C* and *RAD51D* carry genuine ovarian risk but stayed shut; at 31 and 30 carriers they are the two smallest samples in the panel, so this reflects power rather than contradiction. *η* is a logit-scale offset; Δpen is a model-scale carrier − control difference at age 70, given for localisation and relative ordering and *not* as a penetrance estimate.

|  |  | breast |  | ovary |  |
|---|---|---|---|---|---|
| gene | carriers | *η* | Δpen (pp) | gate | *η* |
| *BRCA2* | 240 | 3.51 | +17.18 | open | 2.93 |
| *BRCA1* | 151 | 4.57 | +14.36 | open | **5.13** |
| *PALB2* | 43 | 2.56 | +13.44 | shut | 0.00 |
| *BRIP1* | 92 | 1.52 | +5.17 | open | **3.53** |
| *ATM* | 188 | 0.65 | +3.60 | shut | 0.00 |
| *RAD51D* | 30 | 0.00 | +1.37 | shut | 0.00 |
| *CHEK2* | 236 | 0.00 | +1.30 | shut | 0.00 |
| *RAD51C* | 31 | 0.00 | −0.40 | shut | 0.00 |

**Table S6:** Disease-specific carrier effect *η* for the Figure 3 genes. Gene, system, assigned signature, home disease (the gene’s known disease), model penetrance excess Δpen (carrier control, pp), carrier cumulative-incidence penetrance at age 70, and the population-registry value where available (model / registry KM). Cancers are registry-calibrated, age-varying *η*(*t*); breast is assigned to S19 (Methods). Because the cancer effects here are fitted to registry diagnoses, their Δpen is larger than in the atlases fitted to recorded diagnoses alone (Tables S7 and S8); for *BRCA2* breast cancer, +23.6 here against +14.1 (age-constant) and +12.0 (age-varying) there.

| Gene | System | Sig | Home disease | Δpen (pp) | Carrier @70 | Registry @70 (model / KM) |
|---|---|---|---|---|---|---|
| *BRCA2* | breast (F) | S19 | Breast cancer | +23.6 | 34.1% | 34 / 31% |
| *BRCA1* | breast (F) | S19 | Breast cancer | +24.5 | 36.4% | 36 / 38% |
| *MLH1/MSH2/MSH6/PMS2/EPCAM* | colorectal | S17 | Colorectal cancer | +9.8 | 11.1% | 11 / 11% |
| *BRCA2* | prostate (M) | S13 | Prostate cancer | +7.8 | 13.4% | 13 / 16% |
| *LDLR* | lipid | S5 | Hypercholesterolemia | +15.3 | 30.4% | – |
| *TTN* | cardiac | S0 | Heart failure | +5.9 | 7.9% | – |
| *TTN* | cardiac | S0 | Atrial fibrillation | +8.1 | 12.5% | – |

**Table S7:**
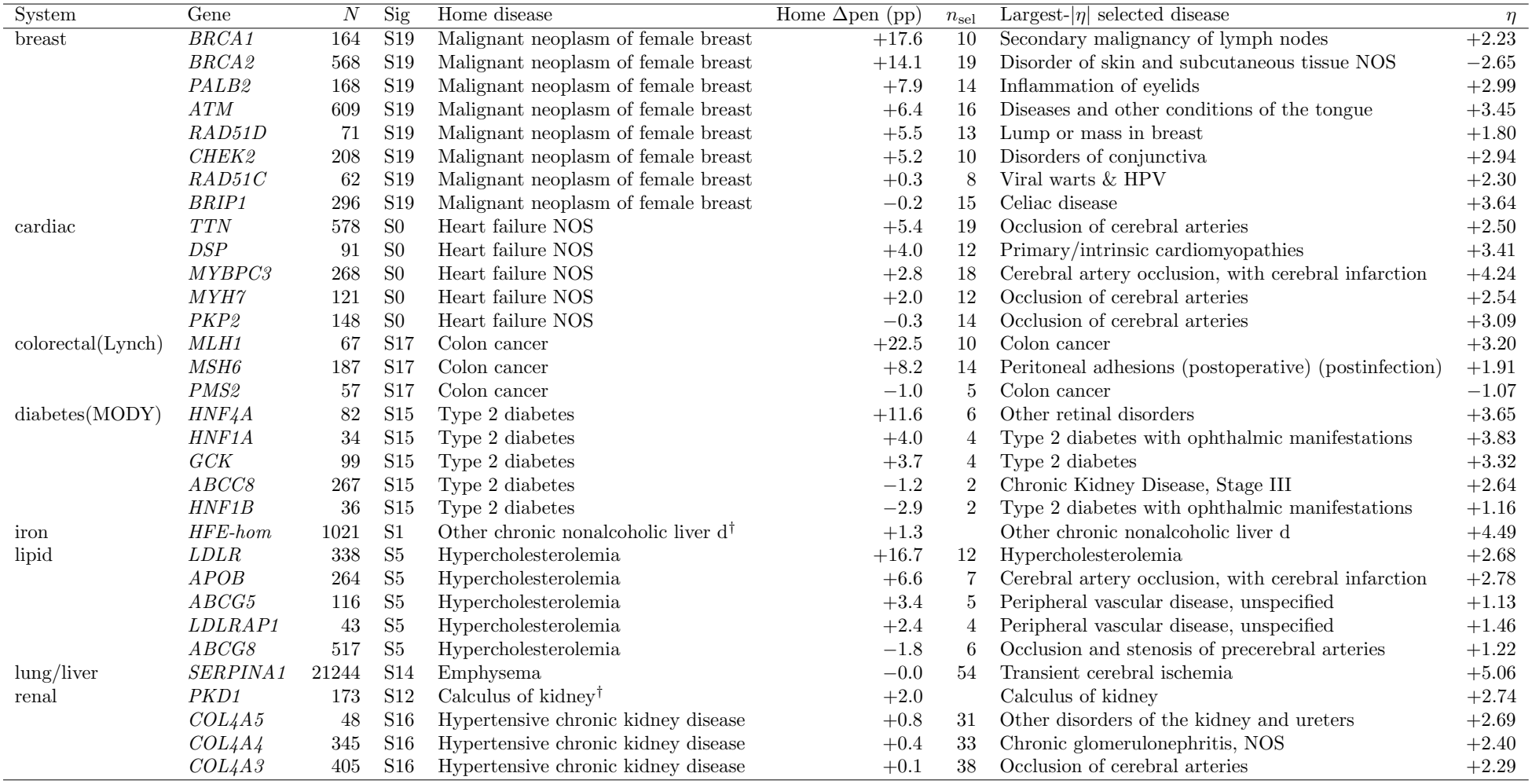
The 32-gene atlas of age-constant carrier effects (eight organ systems). Each gene is assigned to the signature that most strongly expresses its known disease, and the model selects which diseases change. *Home disease* and *Home* Δ*pen*: the gene’s known disease and its carrier − control difference in cumulative incidence at age 70 (percentage points), on the probability scale and so comparable across genes. *n*_sel_: number of selected diseases. *Largest-* |*η*| *selected disease* and *η*: the selected disease with the largest carrier effect on the log-odds scale; for a rare disease a large log-odds effect can mean a small change in risk, so this is often not the home disease. ^†^*PKD1* and *HFE* homozygotes were fit separately (Methods); their row shows the first selected disease in place of a home disease. These values are fitted to recorded diagnoses only, not to the cancer registry; penetrance calibrated to the cancer registry is in Fig. 3d and Table S6.

| System | Gene | *N* | Sig | Home disease | Home Δpen (pp) | *n*<sub>sel</sub> | Largest-\|*η*\| selected disease | *η* |
|---|---|---|---|---|---|---|---|---|
| breast | *BRCA1* | 164 | S19 | Malignant neoplasm of female breast | +17.6 | 10 | Secondary malignancy of lymph nodes | +2.23 |
|  | *BRCA2* | 568 | S19 | Malignant neoplasm of female breast | +14.1 | 19 | Disorder of skin and subcutaneous tissue NOS | −2.65 |
|  | *PALB2* | 168 | S19 | Malignant neoplasm of female breast | +7.9 | 14 | Inflammation of eyelids | +2.99 |
|  | *ATM* | 609 | S19 | Malignant neoplasm of female breast | +6.4 | 16 | Diseases and other conditions of the tongue | +3.45 |
|  | *RAD51D* | 71 | S19 | Malignant neoplasm of female breast | +5.5 | 13 | Lump or mass in breast | +1.80 |
|  | *CHEK2* | 208 | S19 | Malignant neoplasm of female breast | +5.2 | 10 | Disorders of conjunctiva | +2.94 |
|  | *RAD51C* | 62 | S19 | Malignant neoplasm of female breast | +0.3 | 8 | Viral warts & HPV | +2.30 |
|  | *BRIP1* | 296 | S19 | Malignant neoplasm of female breast | −0.2 | 15 | Celiac disease | +3.64 |
| cardiac | *TTN* | 578 | S0 | Heart failure NOS | +5.4 | 19 | Occlusion of cerebral arteries | +2.50 |
|  | *DSP* | 91 | S0 | Heart failure NOS | +4.0 | 12 | Primary/intrinsic cardiomyopathies | +3.41 |
|  | *MYBPC3* | 268 | S0 | Heart failure NOS | +2.8 | 18 | Cerebral artery occlusion, with cerebral infarction | +4.24 |
|  | *MYH7* | 121 | S0 | Heart failure NOS | +2.0 | 12 | Occlusion of cerebral arteries | +2.54 |
|  | *PKP2* | 148 | S0 | Heart failure NOS | −0.3 | 14 | Occlusion of cerebral arteries | +3.09 |
| colorectal(Lynch) | *MLH1* | 67 | S17 | Colon cancer | +22.5 | 10 | Colon cancer | +3.20 |
|  | *MSH6* | 187 | S17 | Colon cancer | +8.2 | 14 | Peritoneal adhesions (postoperative) (postinfection) | +1.91 |
|  | *PMS2* | 57 | S17 | Colon cancer | −1.0 | 5 | Colon cancer | −1.07 |
| diabetes(MODY) | *HNF4A* | 82 | S15 | Type 2 diabetes | +11.6 | 6 | Other retinal disorders | +3.65 |
|  | *HNF1A* | 34 | S15 | Type 2 diabetes | +4.0 | 4 | Type 2 diabetes with ophthalmic manifestations | +3.83 |
|  | *GCK* | 99 | S15 | Type 2 diabetes | +3.7 | 4 | Type 2 diabetes | +3.32 |
|  | *ABCC8* | 267 | S15 | Type 2 diabetes | −1.2 | 2 | Chronic Kidney Disease, Stage III | +2.64 |
|  | *HNF1B* | 36 | S15 | Type 2 diabetes | −2.9 | 2 | Type 2 diabetes with ophthalmic manifestations | +1.16 |
| iron | *HFE-hom* | 1021 | S1 | Other chronic nonalcoholic liver d<sup>†</sup> | +1.3 |  | Other chronic nonalcoholic liver d | +4.49 |
| lipid | *LDLR* | 338 | S5 | Hypercholesterolemia | +16.7 | 12 | Hypercholesterolemia | +2.68 |
|  | *APOB* | 264 | S5 | Hypercholesterolemia | +6.6 | 7 | Cerebral artery occlusion, with cerebral infarction | +2.78 |
|  | *ABCG5* | 116 | S5 | Hypercholesterolemia | +3.4 | 5 | Peripheral vascular disease, unspecified | +1.13 |
|  | *LDLRAP1* | 43 | S5 | Hypercholesterolemia | +2.4 | 4 | Peripheral vascular disease, unspecified | +1.46 |
|  | *ABCG8* | 517 | S5 | Hypercholesterolemia | −1.8 | 6 | Occlusion and stenosis of precerebral arteries | +1.22 |
| lung/liver | *SERPINA1* | 21244 | S14 | Emphysema | −0.0 | 54 | Transient cerebral ischemia | +5.06 |
| renal | *PKD1* | 173 | S12 | Calculus of kidney<sup>†</sup> | +2.0 |  | Calculus of kidney | +2.74 |
|  | *COL4A5* | 48 | S16 | Hypertensive chronic kidney disease | +0.8 | 31 | Other disorders of the kidney and ureters | +2.69 |
|  | *COL4A4* | 345 | S16 | Hypertensive chronic kidney disease | +0.4 | 33 | Chronic glomerulonephritis, NOS | +2.40 |
|  | *COL4A3* | 405 | S16 | Hypertensive chronic kidney disease | +0.1 | 38 | Occlusion of cerebral arteries | +2.29 |

**Table S8:** The age-varying (*η*(*t*)) carrier atlas. Companion to the age-constant Table S7: here *η* is age-varying and the model is fit to carriers and matched controls with the population parameters fixed (not fitted to the registry), so Δpen_70_ differs from the registry-calibrated values in Table S6. Δpen_70_ = carrier − control penetrance difference (pp) at the home disease at age 70; *peak* = age of maximal |*η*(*t*) | ; *n*_sel_ = diseases selected by the spike-and-slab prior; the top age-varying diseases are the other diseases selected on the assigned signature.

| System | Gene | *N* | Sig | Home disease | Δpen<sub>70</sub> | peak | *n*<sub>sel</sub> | Top time-varying reshaped diseases |
|---|---|---|---|---|---|---|---|---|
| lipid | *LDLR* | 338 | S5 | Hypercholesterolemia | +13.7 | 61 | 9 | Hypercholesterolemia; Hyperlipidemia; Coronary atheroscleros |
|  | *APOB* | 264 | S5 | Hypercholesterolemia | +3.5 | 42 | 9 | Hypercholesterolemia; Other chronic ischemic; Coronary atheroscleros |
|  | *ABCG5* | 116 | S5 | Hypercholesterolemia | +2.2 | 76 | 7 | Other chronic ischemic; Hypercholesterolemia; Coronary atheroscleros |
|  | *LDLRAP1* | 43 | S5 | Hypercholesterolemia | +2.0 | 81 | 5 | Angina pectoris; Myocardial infarction; Other chronic ischemic |
|  | *LPL* | 283 | S5 | Hyperlipidemia | +1.0 | 54 | 9 | Hyperlipidemia; Hypercholesterolemia; Other chronic ischemic |
|  | *LMF1* | 63 | S5 | Hyperlipidemia | +0.4 | 72 | 6 | Other chronic ischemic; Coronary atheroscleros; Essential hypertension |
|  | *ABCG8* | 517 | S5 | Hypercholesterolemia | −1.0 | 41 | 8 | Essential hypertension; Myocardial infarction; Other chronic ischemic |
| breast (cancer) | *BRCA2* | 568 | S19 | Malignant neoplasm of fe | +12.0 | 66 | 12 | Secondary malignancy o; Malignant neoplasm of ; Disorder of skin and s |
|  | *BRCA1* | 164 | S19 | Malignant neoplasm of fe | +10.3 | 48 | 4 | Malignant neoplasm of ; Secondary malignancy o; Benign neoplasm of ski |
|  | *ATM* | 609 | S19 | Malignant neoplasm of fe | +5.2 | 44 | 17 | Secondary malignancy o; Malignant neoplasm of ; Seborrheic keratosis |
|  | *PALB2* | 168 | S19 | Malignant neoplasm of fe | +4.6 | 73 | 6 | Malignant neoplasm of ; Breast cancer [female]; Other non-epithelial c |
|  | *CHEK2* | 208 | S19 | Malignant neoplasm of fe | +2.8 | 60 | 6 | Malignant neoplasm of ; Benign neoplasm of ski; Breast cancer [female] |
|  | *RAD51D* | 71 | S19 | Malignant neoplasm of fe | +2.4 | 81 | 2 | Malignant neoplasm of ; Other non-epithelial c; - |
|  | *RAD51C* | 62 | S19 | Malignant neoplasm of fe | +1.0 | 39 | 1 | Other non-epithelial c; -; - |
|  | *BRIP1* | 296 | S19 | Malignant neoplasm of fe | −0.1 | 33 | 13 | Secondary hypothyroidi; Thyrotoxicosis with or; Disorders of lacrimal |
| colorectal (Lynch) | *MSH6* | 187 | S17 | Colon cancer | +4.4 | 70 | 11 | Colon cancer; Benign neoplasm of col; Anal and rectal polyp |
|  | *MLH1* | 67 | S17 | Colon cancer | +0.5 | 81 | 6 | Benign neoplasm of col; Noninfectious gastroen; Hemorrhage of gastroin |
|  | *PMS2* | 57 | S17 | Colon cancer | −0.1 | 73 | 8 | Noninfectious gastroen; Benign neoplasm of col; Hemorrhage of rectum a |
| cardiac | *TTN* | 578 | S0 | Congestive heart failure | +2.6 | 58 | 15 | Primary/intrinsic card; Congestive heart failu; Heart failure NOS |
|  | *DSP* | 91 | S0 | Congestive heart failure | +0.9 | 51 | 7 | Atrial fibrillation an; Rheumatic disease of t; Congestive heart failu |
|  | *MYH7* | 121 | S0 | Congestive heart failure | +0.5 | 39 | 5 | Rheumatic disease of t; Disease of tricuspid v; Atrial fibrillation an |
|  | *PKP2* | 148 | S0 | Congestive heart failure | +0.5 | 71 | 9 | Other forms of chronic; Rheumatic disease of t; Left bundle branch blo |
|  | *MYBPC3* | 268 | S0 | Congestive heart failure | +0.5 | 43 | 11 | Cardiomegaly; Pleurisy; pleural effu; Primary/intrinsic card |
|  | *KCNQ1* | 283 | S0 | Atrial fibrillation and | +0.1 | 53 | 15 | Cardiomegaly; Rheumatic disease of t; Congenital anomalies o |
|  | *SCN5A* | 104 | S0 | Atrial fibrillation and | −0.0 | 65 | 8 | Heart failure NOS; Cardiomegaly; Other forms of chronic |
| diabetes (MODY) | *HNF4A* | 82 | S15 | Type 2 diabetes | +13.9 | 50 | 1 | Type 2 diabetes; -; - |
|  | *GCK* | 99 | S15 | Type 2 diabetes | +4.9 | 53 | 1 | Type 2 diabetes; -; - |
|  | *HNF1A* | 34 | S15 | Type 2 diabetes | +4.5 | 30 | 0 | -; -; - |
|  | *ABCC8* | 267 | S15 | Type 2 diabetes | −1.1 | 81 | 3 | Type 2 diabetes with o; Diabetic retinopathy; Type 1 diabetes |
|  | *HNF1B* | 36 | S15 | Type 2 diabetes | −1.8 | 63 | 0 | -; -; - |
| renal | *COL4A3* | 405 | S16 | Hypertensive chronic kid | +0.1 | 81 | 36 | Pulmonary heart diseas; Pleurisy; pleural effu; Hemiplegia |
|  | *COL4A4* | 345 | S16 | Hypertensive chronic kid | −0.0 | 62 | 38 | Hypovolemia; Pulmonary collapse; in; Other diseases of resp |
|  | *COL4A5* | 48 | S16 | Hypertensive chronic kid | −0.1 | 53 | 8 | Constipation; Hypotension NOS; Other anemias |
| pulmonary | *SERPINA1* | 3000 | S14 | Emphysema | −0.1 | 47 | 11 | Obstructive chronic br; Pneumococcal pneumonia; Chronic airway obstruc |

**Table S9:** The population model without the carrier term under-predicts monogenic carriers. O/E = share of carriers ever diagnosed with the gene’s focal disease */* mean predicted cumulative incidence of that disease to age 70 (*>* 1 under-predicts), from the held-out population predictions with carrier status not used; breast cancer in women only. The median over the 34 atlas genes is 2.2. Adding the carrier effect *η* brings carriers to the observed level (Fig. 3d).

| gene | *N* | O/E (no carrier term) |
|---|---|---|
| *LDLR* | 338 | 2.24 |
| *APOB* | 264 | 1.21 |
| *BRCA1* | 80 | 3.81 |
| *BRCA2* | 313 | 3.23 |
| *MLH1* | 67 | 10.92 |
| *MSH6* | 187 | 6.14 |
| *TTN* | 578 | 4.73 |
| *CHEK2* | 112 | 2.45 |
| **median (34 genes)** |  | **2.23** |

### Cross-cohort carrier ascertainment

To check that the carrier definition transfers, the same ClinVar P/LP panel and the same carrier rule (exact *chrom:pos:ref:alt* match) were applied to an independent exome cohort, the Mass General Brigham Biobank (53K WES). Only the underlying callset differs. Carrier counts are concordant (median rate ratio 1.19; 39 of 61 genes within two-fold of unity), and the exceptions are mechanical rather than biological (Supp. Table S10): the UK Biobank exome release is filtered for low-complexity and segmental-duplication regions, so genes with paralogues (*PMS2, CHEK2* ) are conservatively ascertained here. This bounds the ascertainment loss for those genes; it does not affect any other result, since carrier counts are independent of the diagnosis data.

**Table S10:**
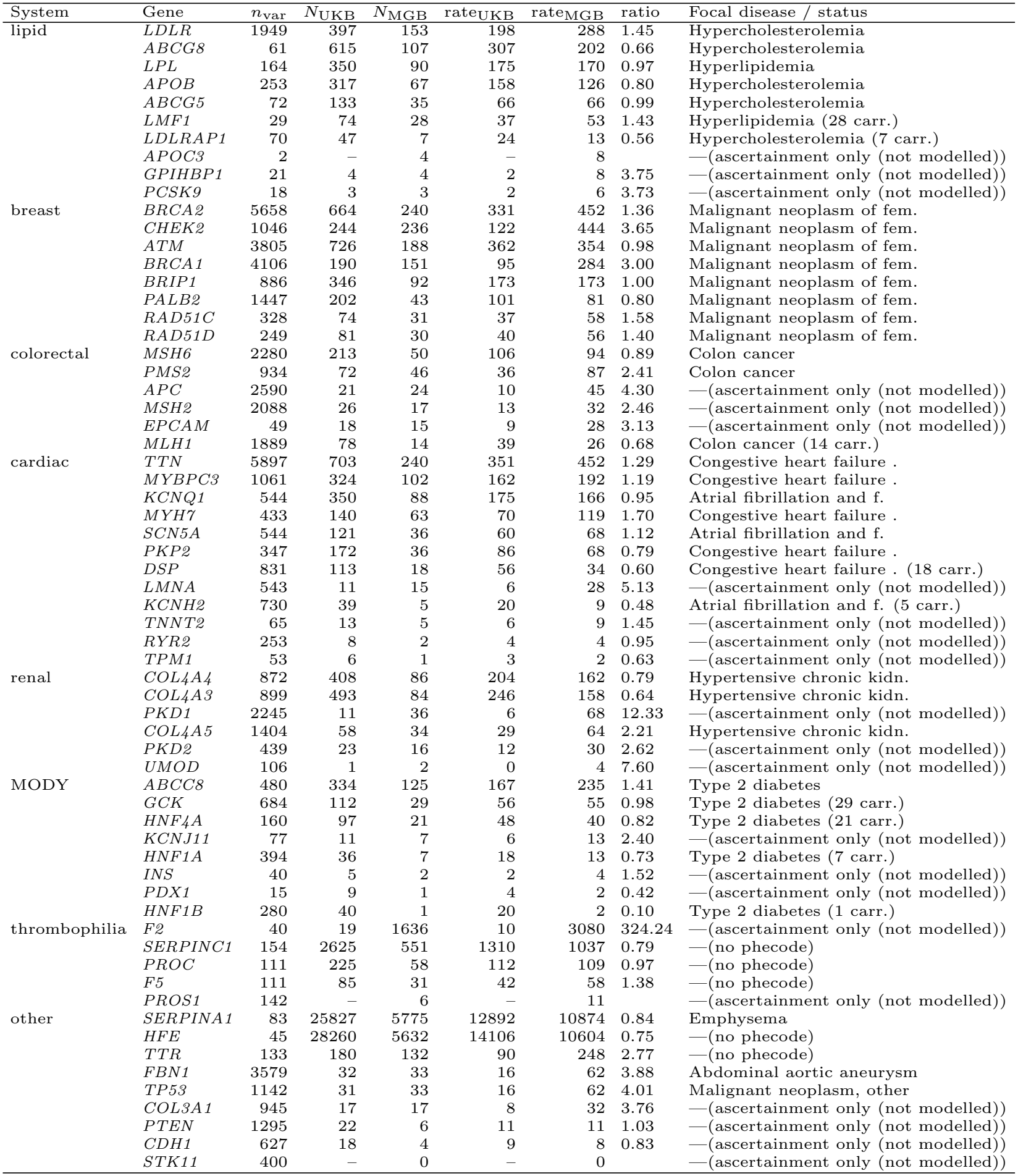
Carrier ascertainment transfers to an independent cohort; the exceptions have specific causes. ClinVar Pathogenic/Likely-pathogenic carriers per gene in UK Biobank 200K WES and MGB 53K WES, using the *same* panel and the *same* carrier rule (exact *chrom:pos:ref:alt*); only the callset differs. Median rate ratio 1.19; 39 of 61 genes fall within two-fold of unity. The discordances have four causes. *Callset filtering:* the UK Biobank exome release is filtered to remove low-complexity repeats and segmental duplications, whereas the MGB release has no such region mask; genes with paralogues are therefore conservatively ascertained in UK Biobank (*PMS2*, whose pseudogene *PMS2CL* is ∼ 98% identical, loses 93% of its panel positions against an ∼ 80% baseline, and *CHEK2* loses *1100delC* entirely). *Assay design: F2 G20210A* is 3’UTR and outside exome capture, so no filtering choice recovers it. *Ancestry:* ratios below unity concentrate in genes whose common pathogenic allele is Northern-European enriched (*HFE* C282Y, *SERPINA1* PI*Z), consistent with UK Biobank being ∼ 94% European; *TTR* V122I runs the other way, being African-ancestry enriched. *Small n:* extreme ratios on single-digit counts are counting noise. Carrier counts are independent of the diagnosis data and of the model.

| System | Gene | *n*<sub>var</sub> | *N*<sub>UKB</sub> | *N*<sub>MGB</sub> | rate<sub>UKB</sub> | rate<sub>MGB</sub> | ratio | Focal disease / status |
|---|---|---|---|---|---|---|---|---|
| lipid | *LDLR* | 1949 | 397 | 153 | 198 | 288 | 1.45 | Hypercholesterolemia |
|  | *ABCG8* | 61 | 615 | 107 | 307 | 202 | 0.66 | Hypercholesterolemia |
|  | *LPL* | 164 | 350 | 90 | 175 | 170 | 0.97 | Hyperlipidemia |
|  | *APOB* | 253 | 317 | 67 | 158 | 126 | 0.80 | Hypercholesterolemia |
|  | *ABCG5* | 72 | 133 | 35 | 66 | 66 | 0.99 | Hypercholesterolemia |
|  | *LMF1* | 29 | 74 | 28 | 37 | 53 | 1.43 | Hyperlipidemia (28 carr.) |
|  | *LDLRAP1* | 70 | 47 | 7 | 24 | 13 | 0.56 | Hypercholesterolemia (7 carr.) |
|  | *APOC3* | 2 | – | 4 | – | 8 |  | —(ascertainment only (not modelled)) |
|  | *GPIHBP1* | 21 | 4 | 4 | 2 | 8 | 3.75 | —(ascertainment only (not modelled)) |
|  | *PCSK9* | 18 | 3 | 3 | 2 | 6 | 3.73 | —(ascertainment only (not modelled)) |
| breast | *BRCA2* | 5658 | 664 | 240 | 331 | 452 | 1.36 | Malignant neoplasm of fem. |
|  | *CHEK2* | 1046 | 244 | 236 | 122 | 444 | 3.65 | Malignant neoplasm of fem. |
|  | *ATM* | 3805 | 726 | 188 | 362 | 354 | 0.98 | Malignant neoplasm of fem. |
|  | *BRCA1* | 4106 | 190 | 151 | 95 | 284 | 3.00 | Malignant neoplasm of fem. |
|  | *BRIP1* | 886 | 346 | 92 | 173 | 173 | 1.00 | Malignant neoplasm of fem. |
|  | *PALB2* | 1447 | 202 | 43 | 101 | 81 | 0.80 | Malignant neoplasm of fem. |
|  | *RAD51C* | 328 | 74 | 31 | 37 | 58 | 1.58 | Malignant neoplasm of fem. |
|  | *RAD51D* | 249 | 81 | 30 | 40 | 56 | 1.40 | Malignant neoplasm of fem. |
| colorectal | *MSH6* | 2280 | 213 | 50 | 106 | 94 | 0.89 | Colon cancer |
|  | *PMS2* | 934 | 72 | 46 | 36 | 87 | 2.41 | Colon cancer |
|  | *APC* | 2590 | 21 | 24 | 10 | 45 | 4.30 | —(ascertainment only (not modelled)) |
|  | *MSH2* | 2088 | 26 | 17 | 13 | 32 | 2.46 | —(ascertainment only (not modelled)) |
|  | *EPCAM* | 49 | 18 | 15 | 9 | 28 | 3.13 | —(ascertainment only (not modelled)) |
|  | *MLH1* | 1889 | 78 | 14 | 39 | 26 | 0.68 | Colon cancer (14 carr.) |
| cardiac | *TTN* | 5897 | 703 | 240 | 351 | 452 | 1.29 | Congestive heart failure . |
|  | *MYBPC3* | 1061 | 324 | 102 | 162 | 192 | 1.19 | Congestive heart failure . |
|  | *KCNQ1* | 544 | 350 | 88 | 175 | 166 | 0.95 | Atrial fibrillation and f. |
|  | *MYH7* | 433 | 140 | 63 | 70 | 119 | 1.70 | Congestive heart failure . |
|  | *SCN5A* | 544 | 121 | 36 | 60 | 68 | 1.12 | Atrial fibrillation and f. |
|  | *PKP2* | 347 | 172 | 36 | 86 | 68 | 0.79 | Congestive heart failure . |
|  | *DSP* | 831 | 113 | 18 | 56 | 34 | 0.60 | Congestive heart failure . (18 carr.) |
|  | *LMNA* | 543 | 11 | 15 | 6 | 28 | 5.13 | —(ascertainment only (not modelled)) |
|  | *KCNH2* | 730 | 39 | 5 | 20 | 9 | 0.48 | Atrial fibrillation and f. (5 carr.) |
|  | *TNNT2* | 65 | 13 | 5 | 6 | 9 | 1.45 | —(ascertainment only (not modelled)) |
|  | *RYR2* | 253 | 8 | 2 | 4 | 4 | 0.95 | —(ascertainment only (not modelled)) |
|  | *TPM1* | 53 | 6 | 1 | 3 | 2 | 0.63 | —(ascertainment only (not modelled)) |
| renal | *COL4A4* | 872 | 408 | 86 | 204 | 162 | 0.79 | Hypertensive chronic kidn. |
|  | *COL4A3* | 899 | 493 | 84 | 246 | 158 | 0.64 | Hypertensive chronic kidn. |
|  | *PKD1* | 2245 | 11 | 36 | 6 | 68 | 12.33 | —(ascertainment only (not modelled)) |
|  | *COL4A5* | 1404 | 58 | 34 | 29 | 64 | 2.21 | Hypertensive chronic kidn. |
|  | *PKD2* | 439 | 23 | 16 | 12 | 30 | 2.62 | —(ascertainment only (not modelled)) |
|  | *UMOD* | 106 | 1 | 2 | 0 | 4 | 7.60 | —(ascertainment only (not modelled)) |
| MODY | *ABCC8* | 480 | 334 | 125 | 167 | 235 | 1.41 | Type 2 diabetes |
|  | *GCK* | 684 | 112 | 29 | 56 | 55 | 0.98 | Type 2 diabetes (29 carr.) |
|  | *HNF4A* | 160 | 97 | 21 | 48 | 40 | 0.82 | Type 2 diabetes (21 carr.) |
|  | *KCNJ11* | 77 | 11 | 7 | 6 | 13 | 2.40 | —(ascertainment only (not modelled)) |
|  | *HNF1A* | 394 | 36 | 7 | 18 | 13 | 0.73 | Type 2 diabetes (7 carr.) |
|  | *INS* | 40 | 5 | 2 | 2 | 4 | 1.52 | —(ascertainment only (not modelled)) |
|  | *PDX1* | 15 | 9 | 1 | 4 | 2 | 0.42 | —(ascertainment only (not modelled)) |
|  | *HNF1B* | 280 | 40 | 1 | 20 | 2 | 0.10 | Type 2 diabetes (1 carr.) |
| thrombophilia | *F2* | 40 | 19 | 1636 | 10 | 3080 | 324.24 | —(ascertainment only (not modelled)) |
|  | *SERPINC1* | 154 | 2625 | 551 | 1310 | 1037 | 0.79 | —(no phecode) |
|  | *PROC* | 111 | 225 | 58 | 112 | 109 | 0.97 | —(no phecode) |
|  | *F5* | 111 | 85 | 31 | 42 | 58 | 1.38 | —(no phecode) |
|  | *PROS1* | 142 | – | 6 | – | 11 |  | —(ascertainment only (not modelled)) |
| other | *SERPINA1* | 83 | 25827 | 5775 | 12892 | 10874 | 0.84 | Emphysema |
|  | *HFE* | 45 | 28260 | 5632 | 14106 | 10604 | 0.75 | —(no phecode) |
|  | *TTR* | 133 | 180 | 132 | 90 | 248 | 2.77 | —(no phecode) |
|  | *FBN1* | 3579 | 32 | 33 | 16 | 62 | 3.88 | Abdominal aortic aneurysm |
|  | *TP53* | 1142 | 31 | 33 | 16 | 62 | 4.01 | Malignant neoplasm, other |
|  | *COL3A1* | 945 | 17 | 17 | 8 | 32 | 3.76 | —(ascertainment only (not modelled)) |
|  | *PTEN* | 1295 | 22 | 6 | 11 | 11 | 1.03 | —(ascertainment only (not modelled)) |
|  | *CDH1* | 627 | 18 | 4 | 9 | 8 | 0.83 | —(ascertainment only (not modelled)) |
|  | *STK11* | 400 | – | 0 | – | 0 |  | —(ascertainment only (not modelled)) |

**Table S11:**
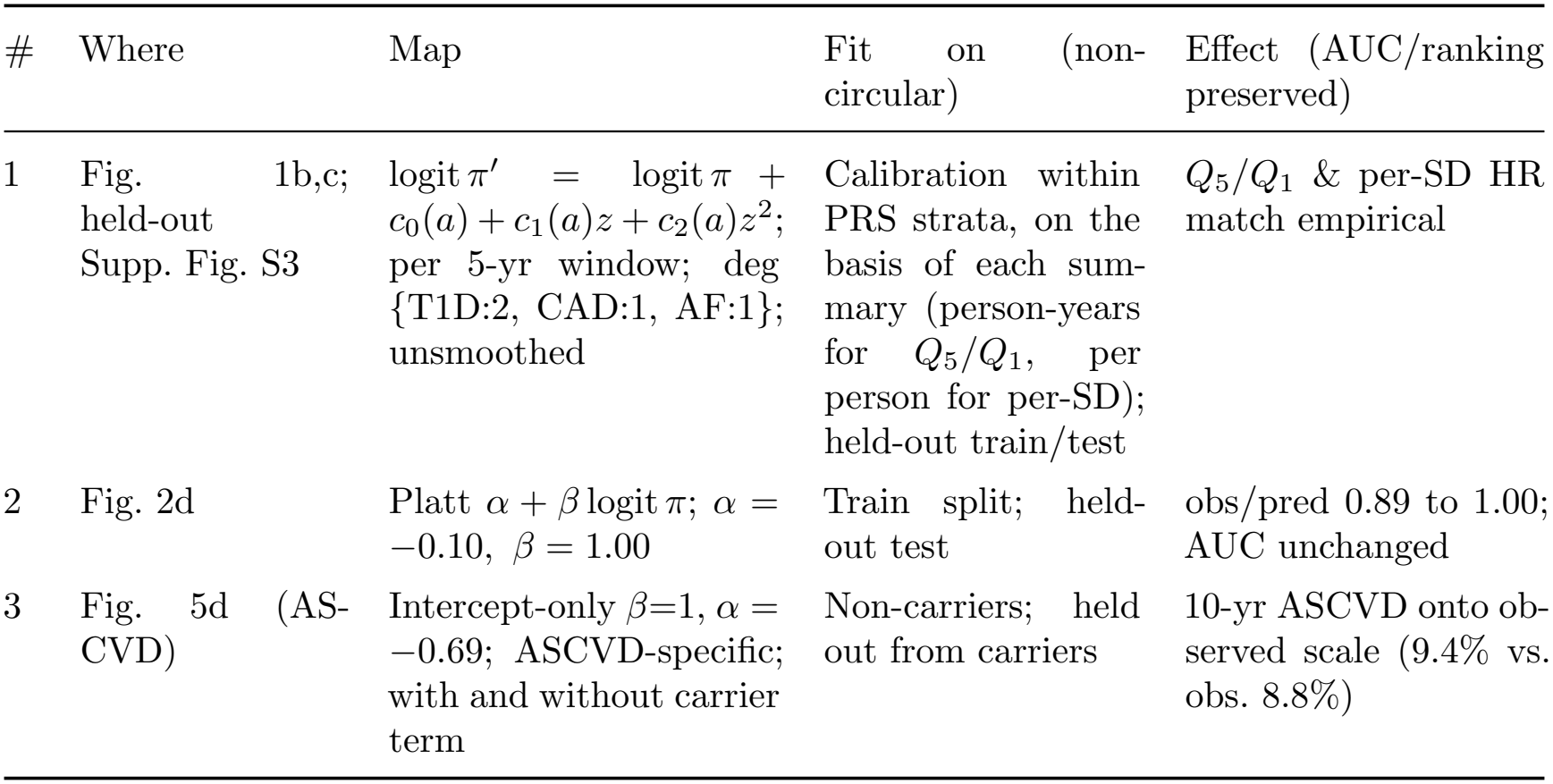
Every recalibration used in the paper. All three maps are *monotone in the predicted probability π*, so ranking (AUC) is preserved; each is fit on a held-out split or on a group other than the one being validated (controls, or non-carriers). The age-interaction model is reported natively; Fig. 5d applies map 3.

| # | Where | Map | Fit on (non-circular) | Effect (AUC/ranking preserved) |
|---|---|---|---|---|
| 1 | Fig. 1b,c; held-out Supp. Fig. S3 | logit π′ = logit π + c<sub>0</sub>(a) + c<sub>1</sub>(a)z + c<sub>2</sub>(a)z<sup>2</sup>; per 5-yr window; deg {T1D:2, CAD:1, AF:1}; unsmoothed | Calibration within PRS strata, on the basis of each sum-mary (person-years for Q<sub>5</sub>/Q<sub>1</sub>, per person for per-SD); held-out train/test | Q<sub>5</sub>/Q<sub>1</sub> & per-SD HR match empirical |
| 2 | Fig. 2d | Platt α + β logit π; α = −0.10, β = 1.00 | Train split; held-out test | obs/pred 0.89 to 1.00; AUC unchanged |
| 3 | Fig. 5d (AS-CVD) | Intercept-only β=1, α = −0.69; ASCVD-specific; with and without carrier term | Non-carriers; held out from carriers | 10-yr ASCVD onto ob-served scale (9.4% vs. obs. 8.8%) |

## References

[1] Lorelei A Mucci, Jacob B Hjelmborg, Jennifer R Harris, Kamila Czene, David J Havelick, Thomas Scheike, Rebecca E Graff, Klaus Holst, Sören Müller, Robert H Unger, Christina McIntosh, Elizabeth Nuttall, Ingunn Brandt, Kathryn L Penney, Mikael Hartman, Peter Kraft, Giovanni Parmigiani, Kaare Christensen, Markku Koskenvuo, Niels V Holm, Kauko Heikkil, Eero Pukkala, Axel Skytthe, Hans-Olov Adami, Jaakko Kaprio, and Nordic Twin Study of Cancer (NorTwinCan) Collaboration. Familial risk and heritability of cancer among twins in nordic countries. JAMA, 315(1):68–76, Jan 2016. doi: 10.1001/jama.2015.17703. URL http://dx.doi.org/10.1001/jama.2015.17703.

[2] Tinca J. C. Polderman, Beben Benyamin, Christiaan A. de Leeuw, Patrick F. Sullivan, Arjen van Bochoven, Peter M. Visscher, and Danielle Posthuma. Meta-analysis of the heritability of human traits based on fifty years of twin studies. Nature Genetics, 47(7):702–709, 2015. doi: 10.1038/ng.3285.

[3] Nina Mars, Jukka T. Koskela, Pietari Ripatti, Tuomo T. J. Kiiskinen, Aki S. Havulinna, Joni V. Lindbohm, Ari Ahola-Olli, Mitja Kurki, Juha Karjalainen, Priit Palta FinnGen, Benjamin M. Neale, Mark Daly, Veikko Salomaa, Aarno Palotie, Elisabeth Widén, and Samuli Ripatti. Polygenic and clinical risk scores and their impact on age at onset and prediction of cardiometabolic diseases and common cancers. Nature Medicine, 26(4):549–557, 2020. doi: 10.1038/s41591-020-0800-0.

[4] Xilin Jiang, Chris Holmes, and Gil McVean. The impact of age on genetic risk for common diseases. PLOS Genetics, 17(8):e1009723, 2021. doi: 10.1371/journal.pgen.1009723.

[5] Xilin Jiang, Alkes L. Price, John Danesh, Gil McVean, Michael Inouye, Sarah Urbut, and Arun Durvasula. Exposure accumulation drives age-dependent disease architectures and polygenic risk scores. medRxiv, 2025. doi: 10.1101/2025.08.29.25334745. Preprint, 2025.08.29.25334745; PMID 40950493; PMCID PMC12424925.

[6] Karoline B. Kuchenbaecker, John L. Hopper, Daniel R. Barnes, Kelly-Anne Phillips, Thea M. Mooij, Marie-José Roos-Blom, et al. Risks of breast, ovarian, and contralateral breast cancer for BRCA1 and BRCA2 mutation carriers. JAMA, 317(23):2402–2416, 2017. doi: 10.1001/jama.2017.7112.

[7] Mev Dominguez-Valentin, Julian R. Sampson, Toni T. Seppälä, Sanne W. ten Broeke, John-Paul Plazzer, Sigve Nakken, Christoph Engel, Stefan Aretz, Mark A. Jenkins, Lone Sunde, Inge Bernstein, Gabriel Capella, Francesc Balaguer, Huw Thomas, D. Gareth Evans, John Burn, Marc Greenblatt, Eivind Hovig, Wouter H. de Vos tot Nederveen Cappel, Rolf H. Sijmons, Lucio Bertario, Maria Grazia Tibiletti, Giulia Martina Cavestro, Annika Lindblom, Adriana Della Valle, Francisco Lopez-Köstner, Nathan Gluck, Lior H. Katz, Karl Heinimann, Carlos A. Vaccaro, Reinhard Büttner, Heike Görgens, Elke Holinski-Feder, Monika Morak, Stefanie Holzapfel, Robert Hüneburg, Magnus von Knebel Doeberitz, Markus Loeffler, Nils Rahner, Hans K. Schäckert, Verena Steinke-Lange, Wolff Schmiegel, Deepak Vangala, Kirsi Pylvänäinen, Laura Renkonen-Sinisalo, John L. Hopper, Aung Ko Win, Robert W. Haile, Noralane M. Lindor, Steven Gallinger, Loic Le Marchand, Polly A. Newcomb, Jane C. Figueiredo, Stephen N. Thibodeau, Karin Wadt, Christina Therkildsen, Henrik Okkels, Zohreh Ketabi, Leticia Moreira, Ariadna Sánchez, Miquel Serra-Burriel, Marta Pineda, Matilde Navarro, Ignacio Blanco, Kate Green, Fiona Lalloo, Emma J. Crosbie, James Hill, Oliver G. Denton, and Pål Møller. Cancer risks by gene, age, and gender in 6350 carriers of pathogenic mismatch repair variants: findings from the prospective lynch syndrome database. Genetics in Medicine, 22(1):15–25, 2020. doi: 10.1038/s41436-019-0596-9.

[8] Sean J. Jurgens, Xin Wang, Seung Hoan Choi, et al. Rare coding variant analysis for human diseases across biobanks and ancestries. Nature Genetics, 56:1811–1820, 2024. doi: 10.1038/s41588-024-01894-5.

[9] Petko P. Fiziev, Jeremy McRae, Jacob C. Ulirsch, et al. Rare penetrant mutations confer severe risk of common diseases. Science, 380:eabo1131, 2023. PMID 37262146.

[10] Marissa Kellogg, Andrea Hildebrand, Tia Dinatale, Manaz Rezayee, Jessica Minnier, Andrea L. C. Schneider, Lia Ernst, Michelle Cameron, Elizabeth Gerard, Alica Goldman, Remi Stevelink, Amy Brooks-Kayal, Kathryn M. Pridgen, VA Million Veteran Program, Julie A. Lynch, and Craig C. Teerlink. Pathogenic epilepsy gene variant prevalence and penetrance among U.S. military veterans in the Million Veteran Program cohort. medRxiv, 2026. doi: 10.64898/2026.08.18.26360604. Preprint, 2026.08.18.26360604.

[11] Mitja I. Kurki, Juha Karjalainen, Priit Palta, et al. FinnGen provides genetic insights from a well-phenotyped isolated population. Nature, 613:508–518, 2023. doi: 10.1038/s41586-022-05473-8.

[12] Iain S. Forrest, Kumardeep Chaudhary, Ha My T. Vy, Ben O. Petrazzini, Shantanu Bafna, Daniel M. Jordan, Ghislain Rocheleau, Ruth J. F. Loos, Girish N. Nadkarni, Judy H. Cho, and Ron Do. Population-based penetrance of deleterious clinical variants. JAMA, 327(4):350–359, 2022. doi: 10.1001/jama.2021.23686.

[13] Caroline F. Wright, Luke N. Sharp, Leigh Jackson, Anna Murray, James S. Ware, Daniel G. MacArthur, Heidi L. Rehm, Kashyap A. Patel, and Michael N. Weedon. Guidance for estimating penetrance of monogenic disease-causing variants in population cohorts. Nature Genetics, 56(9):1772–1779, 2024. doi: 10.1038/s41588-024-01842-3.

[14] Rebecca Kingdom and Caroline F. Wright. Incomplete penetrance and variable expressivity: from clinical studies to population cohorts. Frontiers in Genetics, 13:920390, 2022. doi: 10.3389/fgene.2022.920390.

[15] Rafik Tadros, Catherine Francis, Xiao Xu, et al. Shared genetic pathways contribute to risk of hypertrophic and dilated cardiomyopathies with opposite directions of effect. Nature Genetics, 53:128–134, 2021. PMID 33495596.

[16] W. D. Dotson, M. P. Douglas, K. Kolor, A. C. Stewart, M. S. Bowen, M. Gwinn, A. Wulf, H. M. Anders, C. Q. Chang, M. Clyne, T. K. Lam, S. D. Schully, M. Marrone, W. G. Feero, and M. J. Khoury. Prioritizing Genomic Applications for Action by Level of Evidence: A Horizon-Scanning Method. Clinical Pharmacology & Therapeutics, 95(4):394–402, 2014. doi: 10.1038/clpt.2013.226.

[17] Akl C. Fahed, Minxian Wang, Julian R. Homburger, Aniruddh P. Patel, Alexander G. Bick, Cynthia L. Neben, Carmen Lai, Deanna Brockman, Anthony Philippakis, Patrick T. Ellinor, Christopher A. Cassa, Matthew Lebo, Kenney Ng, Eric S. Lander, Alicia Y. Zhou, Sekar Kathiresan, and Amit V. Khera. Polygenic background modifies penetrance of monogenic variants for tier 1 genomic conditions. Nature Communications, 11(1):3635, 2020. doi: 10.1038/s41467-020-17374-3.

[18] Amit V. Khera, Mark D. Chaffin, Krishna G. Aragam, Mary E. Haas, Jeanette Erdmann, Cristen J. Willer, Daniel I. Chasman, Andre Franke, Marc S. Sabatine, Diego Ardissino, Danish Salesman, Heribert Schunkert, Nilesh J. Samani, and Sekar Kathiresan. Genome-wide polygenic scores for common diseases identify individuals with risk equivalent to monogenic mutations. Nature Genetics, 50(9):1219–1224, 2018. doi: 10.1038/s41588-018-0183-z. URL https://doi.org.

[19] Andrew Lee, Nasim Mavaddat, Amber N. Wilcox, et al. BOADICEA: a comprehensive breast cancer risk prediction model incorporating genetic and nongenetic risk factors. Genetics in Medicine, 21(8):1708–1718, 2019. doi: 10.1038/s41436-018-0406-9.

[20] Tommy Nyberg, Mark N. Brook, Lorenzo Ficorella, et al. CanRisk-Prostate: a comprehensive, externally validated risk model for the prediction of future prostate cancer. Journal of Clinical Oncology, 41(5):1092–1104, 2023. doi: 10.1200/JCO.22.01453.

[21] Sarah M Urbut, Yi Ding, Tetsushi Nakao, Satoshi Koyama, Anika Misra, Xilin Jiang, Achyutha Harish, Leslie Gaffney, Whitney E Hornsby, Jordan W Smoller, Alexander Gusev, Pradeep Natarajan, and Giovanni Parmigiani. A Bayesian framework for longitudinal EHR and genetic discovery. Nature, 656(8129):924–935, 2026. doi: 10.1038/s41586-026-10780-5.

[22] Xilin Jiang, Martin Jinye Zhang, Yidong Zhang, Arun Durvasula, Michael Inouye, Chris Holmes, Alkes L. Price, and Gil McVean. Age-dependent topic modeling of comorbidities in UK Biobank identifies disease subtypes with differential genetic risk. Nature Genetics, 55: 1854–1865, 2023. doi: 10.1038/s41588-023-01522-8.

[23] Artem Shmatko, Alexander Wolfgang Jung, Kumar Gaurav, others, and Moritz Gerstung. Learning the natural history of human disease with generative transformers. Nature, 647: 248–256, 2025. doi: 10.1038/s41586-025-09529-3.

[24] Chris German, Suyash Shringarpure, Payam Dibaeinia, James Ashenhurst, Bertram L. Koelsch, Adam Auton, and Aly A. Khan. Integrating Polygenic Risk Improves Generative Forecasting of Disease Trajectories. In Pacific Symposium on Biocomputing 2026, volume 31, pages 600–611. World Scientific, 2026. doi: 10.1142/97898198247550044.

[25] Amit V. Khera, Mark Chaffin, Kaitlin H. Wade, et al. Polygenic prediction of weight and obesity trajectories from birth to adulthood. Cell, 177(3):587–596.e9, 2019. doi: 10.1016/j.cell.2019.03.028.

[26] Michael Inouye, Gad Abraham, Christopher P. Nelson, et al. Genomic risk prediction of coronary artery disease in 480,000 adults: implications for primary prevention. Journal of the American College of Cardiology, 72(16):1883–1893, 2018. doi: 10.1016/j.jacc.2018.07.079.

[27] Sakari Jukarainen, Tuomo Kiiskinen, Sara Kuitunen, Aki S. Havulinna, Juha Karjalainen, Mattia Cordioli, Joel T. Rämö, Nina Mars FinnGen, Kaitlin E. Samocha, Hanna M. Ollila, Matti Pirinen, and Andrea Ganna. Genetic risk factors have a substantial impact on healthy life years. Nature Medicine, 28(9):1893–1901, 2022. doi: 10.1038/s41591-022-01957-2.

[28] Clare Bycroft, Colin Freeman, Desislava Petkova, Gavin Band, Lloyd T. Elliott, Kevin Sharp, Allan Motyer, Damjan Vukcevic, Olivier Delaneau, Jared O’Connell, Adrian Cortes, Samantha Welsh, Alan Young, Mark Effingham, Gil McVean, Stephen Leslie, Naomi Allen, Peter Donnelly, and Jonathan Marchini. The UK Biobank resource with deep phenotyping and genomic data. Nature, 562(7726):203–209, 2018. doi: 10.1038/s41586-018-0579-z.

[29] Deborah J. Thompson, Daniel Wells, Saskia Selzam, Iliana Peneva, Rachel Moore, Kevin Sharp, William A. Tarran, Edward J. Beard, Fernando Riveros-Mckay, Carla Giner-Delgado, Duncan Palmer, Priyanka Seth, James Harrison, Marta Futema, Gil McVean, Vincent Plagnol, Peter Donnelly, and Michael E. Weale. A systematic evaluation of the performance and properties of the UK Biobank polygenic risk score (PRS) release. PLOS ONE, 19(9):e0307270, 2024. doi: 10.1371/journal.pone.0307270. Genomics England Research Consortium.

[30] Melissa J. Landrum, Jennifer M. Lee, Mark Benson, Garth R. Brown, Chen Chao, Shanmuga Chitipiralla, Baoshan Gu, Jennifer Hart, Douglas Hoffman, Wonhee Jang, Karen Karapetyan, Kenneth Katz, Chunlei Liu, Zenith Maddipatla, Adriana Malheiro, Kurt McDaniel, Michael Ovetsky, George Riley, George Zhou, J. Bradley Holmes, Brandi L. Kattman, and Donna R. Maglott. ClinVar: improving access to variant interpretations and supporting evidence. Nucleic Acids Research, 46(D1):D1062–D1067, 2018. doi: 10.1093/nar/gkx1153.

[31] Joseph D. Szustakowski, Suganthi Balasubramanian, Erika Kvikstad, Shareef Khalid, Paola G. Bronson, Ariella Sasson, Emily Wong, Daren Liu, J. Wade Davis, Carolina Haefliger, A. Katrina Loomis, Rajesh Mikkilineni, Hyun Ji Noh, Samir Wadhawan, Xiaodong Bai, Alicia Hawes, Olga Krasheninina, Ricardo Ulloa, Alexander E. Lopez, Erika N. Smith, Jeffrey F. Waring, Christopher D. Whelan, Ellen A. Tsai, John D. Overton, William J. Salerno, Howard Jacob, Sandor Szalma, Heiko Runz, Gregory Hinkle, Paul Nioi, Slavé Petrovski Melissa R. Miller, Aris Baras, Lyndon J. Mitnaul, and Jeffrey G. Reid. Advancing human genetics research and drug discovery through exome sequencing of the UK Biobank. Nature Genetics, 53(7):942–948, 2021. doi: 10.1038/s41588-021-00885-0.

[32] Sining Chen and Giovanni Parmigiani. Meta-analysis of BRCA1 and BRCA2 penetrance. Journal of Clinical Oncology, 25(11):1329–1333, 2007. doi: 10.1200/JCO.2006.09.1066.

[33] The All of Us Research Program Investigators. The “All of Us” research program. New England Journal of Medicine, 381(7):668–676, 2019. doi: 10.1056/NEJMsr1809937.

[34] All of Us Research Program Genomics Investigators. Genomic data in the All of Us research program. Nature, 627(8003):340–346, 2024. doi: 10.1038/s41586-023-06957-x. PMID 38374255.

[35] Anders Boeck Jensen, Pope L. Moseley, Tudor I. Oprea, Sabrina Gade Ellesøe, Robert Eriksson, Henriette Schmock, Peter Bjødstrup Jensen, Lars Juhl Jensen, and Søren Brunak. Temporal disease trajectories condensed from population-wide registry data covering 6.2 million patients. Nature Communications, 5:4022, 2014. doi: 10.1038/ncomms5022.

[36] Lisa Bastarache, Jacob J. Hughey, Scott Hebbring, Joy Marlo, Wanke Zhao, Wanting T. Ho, Sara L. Van Driest, Tracy L. McGregor, Jonathan D. Mosley, Quinn S. Wells, et al. Phenotype risk scores identify patients with unrecognized Mendelian disease patterns. Science, 359(6381): 1233–1239, 2018. doi: 10.1126/science.aal4043.

[37] Karoline B. Kuchenbaecker, Lesley McGuffog, Daniel Barrowdale, Andrew Lee, Penny Soucy, Joe Dennis, Susan M. Domchek, Mark Robson, Amanda B. Spurdle, Susan J. Ramus, et al. Evaluation of polygenic risk scores for breast and ovarian cancer risk prediction in BRCA1 and BRCA2 mutation carriers. Journal of the National Cancer Institute, 109(7):djw302, 2017. doi: 10.1093/jnci/djw302.

[38] Joshua C. Denny, Marylyn D. Ritchie, Melissa A. Basford, Jill M. Pulley, Lisa Bastarache, Kristin Brown-Gentry, Deede Wang, Dan R. Masys, Dan M. Roden, and Dana C. Crawford. PheWAS: demonstrating the feasibility of a phenome-wide scan to discover gene–disease associations. Bioinformatics, 26(9):1205–1210, 2010. doi: 10.1093/bioinformatics/btq126.

[39] Patrick Wu, Aliya Gifford, Xiangrui Meng, Xue Li, Harry Campbell, Tim Varley, Juan Zhao, Robert Carroll, Lisa Bastarache, Joshua C. Denny, Evropi Theodoratou, and Wei-Qi Wei. Mapping ICD-10 and ICD-10-CM codes to phecodes: Workflow development and initial evaluation. JMIR Medical Informatics, 7(4):e14325, 2019. doi: 10.2196/14325.

[40] Joshua D. Backman, Alexander H. Li, Anthony Marcketta, Dylan Sun, Joelle Mbatchou, Michael D. Kessler, Christian Benner, Daren Liu, Adam E. Locke, Suganthi Balasubramanian, Ashish Yadav, Nilanjana Banerjee, Christopher E. Gillies, Amy Damask, Simon Liu, Xiaodong Bai, Alicia Hawes, Evan Maxwell, Lauren Gurski, Kyoko Watanabe, Jack A. Kosmicki, Veera Rajagopal, Jason Mighty, Marcus Jones, Lyndon Mitnaul, Eli Stahl, Giovanni Coppola, Eric Jorgenson, Lukas Habegger, William J. Salerno, Alan R. Shuldiner, Luca A. Lotta, John D. Overton, Michael N. Cantor, Jeffrey G. Reid, George Yancopoulos, Hyun Min Kang, Jonathan Marchini, Aris Baras, Gonçalo R. Abecasis, and Manuel A. R. Ferreira. Exome sequencing and analysis of 454,787 UK Biobank participants. Nature, 599(7886):628–634, 2021. doi: 10.1038/s41586-021-04103-z.

[41] M. J. Bayarri, J. O. Berger, and F. Liu. Modularization in Bayesian analysis, with emphasis on analysis of computer models. Bayesian Analysis, 4(1):119–150, 2009. doi: 10.1214/09-BA404.

[42] David T. Frazier and David J. Nott. Posterior risk of modular and semi-modular Bayesian inference. Journal of the American Statistical Association, 121(554):1588–1600, 2026. doi: 10.1080/01621459.2025.2580690.

[43] Christos Louizos, Max Welling, and Diederik P. Kingma. Learning sparse neural networks through L0 regularization. In International Conference on Learning Representations (ICLR), 2018. URL https://openreview.net/forum?id=H1Y8hhg0b.

